# Longitudinal Surface-Based Morphometry Changes in the Hippocampus in Dementia

**DOI:** 10.1101/2025.04.07.25325336

**Authors:** Salah Aziz, Romeo Penheiro, Cassandra Morrison, Peter Zhukovsky, John AE Anderson, the Alzheimer’s Disease Neuroimaging Initiative (ADNI)

## Abstract

The hippocampus is central to Alzheimer’s disease (AD), characterized by atrophy and cognitive decline. While volume loss is well-documented, surface-based morphometric (SBM) features—curvature, gyrification, and thickness—remain less explored. Using T1-weighted MRI data from the Alzheimer’s Disease Neuroimaging Initiative (4,617 timepoints; CN: 475, MCI: 673, AD: 269), hippocampal subfields were analyzed with HippUnfold. Linear mixed effects models examined volume and SBM changes, tracking cognitive trajectories in stable (n = 1017) and progressing (n = 301) individuals.

Focusing on CA1, the stable AD group showed significant volume reductions (β = −1.01, p < .001) compared to CN (β = 6.19, p < .001). SBM metrics significantly increased over time across subregions (e.g., curvature: CN: β = −0.72, p < .001; AD: β = 0.006, p < .001), though gyrification did not reach significance in bilateral CA1, CA3, CA4, and subiculum. Time-dependent interactions indicated progressive volume loss and SBM increases across groups (all p’s < .001). Notably, SBM metrics predicted cognitive improvements in AD.

While volume loss remains a key AD marker, it may not capture early morphometric changes. SBM features provide additional insights, underscoring the need for integrated volumetric and surface-based analyses to refine disease detection and therapeutic strategies.

## 1 Introduction

Progressive hippocampal atrophy is a hallmark of Alzheimer’s disease (AD), crucial for identifying neurodegeneration in AD and its prodromal stage, mild cognitive impairment (MCI). Hippocampal atrophy is linked to memory decline, making it a key focus in AD studies. Most studies of brain structure in AD have concentrated on volumetric changes within the medial temporal lobe. However, surface-based metrics like curvature, gyrification, and thickness remain largely unexplored due to challenges in hippocampal segmentation on routine MRI scans (Insausti & Amaral, 2004; Chang et al., 2018; Wisse et al., 2017; Yushkevich et al., 2015). Although post-mortem studies reveal variability in hippocampal subfields constrained by the folded structure (Duvernoy, 1998), segmentation protocols remain challenging, despite harmonization efforts (La Joie et al., 2020; Olsen et al., 2019; Wisse et al., 2017).

Recent advances in automated segmentation, particularly deep learning with convolutional neural networks, offer faster, more accurate methods than manual segmentation (Goubran et al., 2020; Henschel et al., 2020; Thyreau et al., 2018). Tools like HippUnfold (Dekraker et al., 2021, 2022) enable the extraction of SBM metrics, providing insights beyond volume into surface features sensitive to early neurodegeneration. HippUnfold reconstructs a model of hippocampal tissue, enabling precise subfield parcellation without distortions and capturing metrics like curvature, thickness, and gyrification—each associated with AD-relevant structural changes (Dale et al., 1999; Fischl & Dale, 2000; Schaer et al., 2008).

Additional metrics beyond volume include methods like Scoring by Nonlocal Image Patch Estimator (SNIPE) grading. SNIPE, a non-local patch-based measure of anatomical similarity combined with hippocampal segmentation, has demonstrated enhanced sensitivity over traditional volume-based measures in detecting group differences across the spectrum of cognitive decline (Coupé et al., 2012)

Recent work using SNIPE grading revealed significantly higher classification accuracies for distinguishing between cohorts, outperforming volume-based approaches. (Morrison et al., 2023a; 2023b) These findings suggest that SNIPE grading is not only more sensitive to group differences but also provides stronger associations with cognitive trajectories and disease states than hippocampal volume alone. Building on this, our study takes a complementary approach by incorporating hippocampal morphometry features such as curvature, gyrification, and thickness in addition to volume. By doing so, we aim to further elucidate the structural contributions of the hippocampus to cognition and its sensitivity to group differences across the cognitive decline spectrum.

Our study focuses on hippocampal subfields implicated in AD: CA1-CA4, dentate gyrus (DG), subiculum (Sub), and subiculum-reticular layer of the medial temporal lobe (SRLM). Typically, CA1, DG, and Sub show atrophy in AD, whereas CA2 and CA3 are often spared (Christopher-Hayes et al., 2023). These subfields correlate with cognitive functions impaired in AD, such as episodic memory (Tulving, 1972; Eichenbaum, 2017), with CA1, CA4, and DG volume reductions associated with memory decline in MCI and mild AD (Broadhouse et al., 2019; Kerchner et al., 2012). Other cognitive domains, such as visuospatial processing, executive functioning, and language, are also impacted (Rizzo et al., 2000; Jacobs et al., 2015; Manning et al., 2023).

Using the ADNI dataset, we aim to investigate hippocampal subfield changes in individuals classified as CN, MCI, and AD, as this has not been extensively studied using this method. While previous research has often focused on cross-sectional analyses of hippocampal subfields, our study takes a novel longitudinal approach, examining changes across four ADNI phases (ADNI 1, GO, 2, 3) and covering a range of cognitive stages. We hypothesized that individuals with AD would exhibit significant hippocampal atrophy compared to healthy controls, with pronounced atrophy in the CA1 and subiculum subregions. We further anticipated that these changes would provide unique insights into the progression of cognitive decline over time. Due to the novelty of the metric device, we had limited a priori hypotheses regarding the direction of surface metrics, aside from expecting decreases in cortical thickness. While HippUnfold has mainly been applied cross-sectionally, our study extends its use longitudinally within the ADNI dataset, enabling the tracking of hippocampal metric changes over time and deepening our understanding of AD progression.

## 2 Methods

### 2.1 ADNI

Data used in the preparation of this article were obtained from the Alzheimer’s Disease Neuroimaging Initiative (ADNI) database (adni.loni.usc.edu). The ADNI was launched in 2003 as a public-private partnership, led by Principal Investigator Michael W. Weiner, MD. The primary goal of ADNI has been to test whether serial magnetic resonance imaging (MRI), positron emission tomography (PET), other biological markers, and clinical and neuropsychological assessment can be combined to measure the progression of mild cognitive impairment (MCI) and early Alzheimer’s disease (AD) (for more information, visit http://www.adni-info.org). The study received ethical approval from the review boards of all participating institutions. Written informed consent was obtained from participants or their study partner.

### 2.2 ADNI Study Data

MRI metadata, neuropsychological data, and demographic data were obtained on ADNI’s public data website, and merged with the HippUnfold metric data. We used rolling joins to eliminate scans from the study in which the relevant neuropsychological batteries were greater than 180 days from the MRI scan, as the next sequence of scans were scheduled 6 months apart from each other (Morrison et al. 2023a). We obtained demographic data and cognitive scores from the ADNI study data portal. On the recommendation of the Center for Psychometric Analysis of Aging and Neurodegeneration (CPAAN) at University of Washington for ADNI, we used the data harmonized from the Alzheimer’s Disease Sequencing Project Harmonization Consortium (ADSP-PHC), an attempt to harmonize cognitive domain scores across multiple studies of aging (Mukherjee et al., 2023). Cognitive composite scores are divided into four domains: memory (PHC_MEM), executive functioning (PHC_EXF), visuospatial processing (PHC_VSP), and language processing (PHC_LAN). The composite scores were standardized using Z-standardization. The analysis utilized data from the ADNI study, which included some missing neuropsychological measures across participants. This missing data is noted in the analysis.

### 2.3 Participants

In this study, we emphasize the significance of participant retention throughout the entire duration of the longitudinal study. By maintaining a consistent cohort of participants in our data selection, we would be able to gain a more comprehensive understanding of the progression of cognitive decline, enabling us to capture the genuine, dynamic nature of its changes, as well as to ensure power and internal validity in our study (Fewtrell et al., 2008; Siddiqi et al., 2008; Gupta et al., 2015).

In general, eligibility criteria for ADNI include ages 55-90, having a baseline diagnosis of either CN, MCI or early-stage AD, a minimum of six years of education, a sufficient command of English to ensure they can adequately complete the study assessments, and being able to undergo MRI scanning procedures, exclusions include scans within one year of enrollment showing infection, infarction, or focal lesions; bodily augmentations or foreign objects interfering with MRI; and significant systemic illnesses or unstable medical conditions that hinder protocol compliance. Other inclusion and exclusion criteria can be found at https://adni.loni.usc.edu/help-faqs/adni-documentation/.

To ensure the consistency and reliability of our findings, we excluded participants whose diagnostic status regressed at any point in their time during the ADNI study, as it would bring uncertainty as to the interpretation of cognitive trajectories. Attrition rates in the ADNI cohorts are particularly high (Insel et al., 2019), and for this reason, we chose to select participants that had at least two consecutive scans throughout their involvement.

Although here we reference the AD diagnosis as such, our identification of diagnosis of participants with Alzheimer’s disease is based on a *clinical* diagnosis in ADNI, and not one founded on neurobiological identification of tau and amyloid deposits.

A total of 1,429 participants were initially included in this study. After excluding 27 participants due to segmentation failures, pre-processing issues, or visible artifacts in scans, and an additional 87 participants due to fluctuations in diagnosis, the final number of participants analysed was N = 1,315, with an average of 3.51 annual scans (4,617 timepoints). Populations of diagnostic groups include cognitively-normal (CN): 475; mild-cognitive impairment (MCI): 673; Alzheimer’s disease (AD): 269.

To better identify the trajectory of cognitive impairment of the participants, we categorized the participants by the clinical diagnosis measured at their final timepoint in contrast to their baseline diagnostic group. We made the following categorical scheme of trajectories: *stable*, or *non-progressors*, (n = 1017): (CN, n = 411; MCI, n = 366; AD, n = 240) and *progressor*s (n =301): (CN to MCI, n = 51; CN to AD, n =23; MCI to AD, n = 227). We chose to group together the CN progressors due to a small sample size.

**Table 1.**
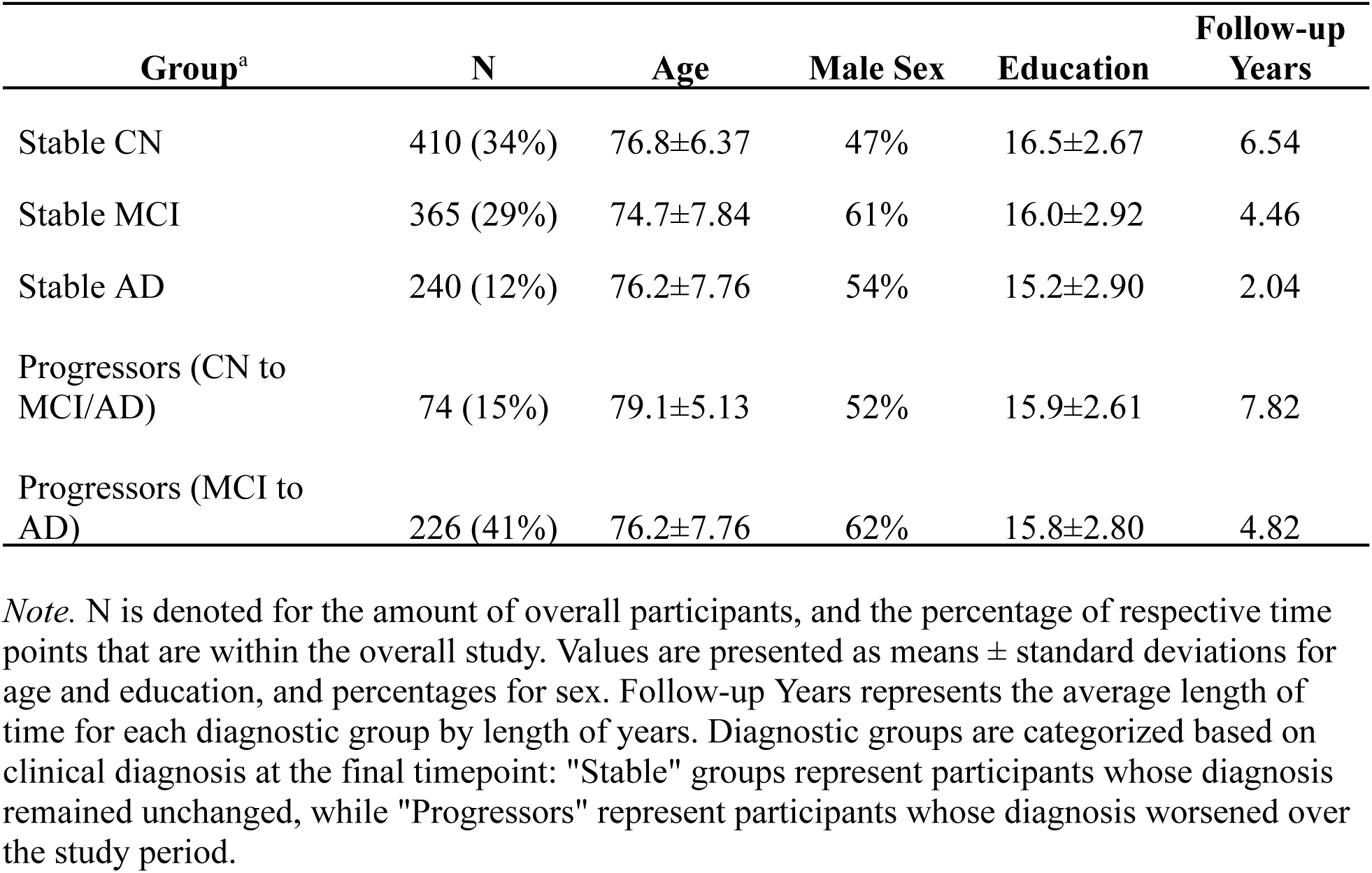
Demographic Information by Group.

| Group<sup>a</sup> | N | Age | Male Sex | Education | Follow-up Years |
| --- | --- | --- | --- | --- | --- |
| Stable CN | 410 (34%) | 76.8±6.37 | 47% | 16.5±2.67 | 6.54 |
| Stable MCI | 365 (29%) | 74.7±7.84 | 61% | 16.0±2.92 | 4.46 |
| Stable AD | 240 (12%) | 76.2±7.76 | 54% | 15.2±2.90 | 2.04 |
| Progressors (CN to MCI/AD) | 74 (15%) | 79.1±5.13 | 52% | 15.9±2.61 | 7.82 |
| Progressors (MCI to AD) | 226 (41%) | 76.2±7.76 | 62% | 15.8±2.80 | 4.82 |

*Note.* N is denoted for the amount of overall participants, and the percentage of respective time points that are within the overall study. Values are presented as means ± standard deviations for age and education, and percentages for sex. Follow-up Years represents the average length of time for each diagnostic group by length of years. Diagnostic groups are categorized based on clinical diagnosis at the final timepoint: "Stable" groups represent participants whose diagnosis remained unchanged, while "Progressors" represent participants whose diagnosis worsened over the study period.

### 2.4 Structural MRI Acquisition and HippUnfold Processing

Participants underwent scanning using 1.5T and 3.0T T1-weighted protocols. Full criteria and details available at http://www.adni-info.org. Original raw T1-weighted scans were retrieved from the ADNI database and processed using HippUnfold v.1.1 on CBRAIN (Sherif et al., 2014). The hippocampal data were first intensity-corrected with the N4 algorithm from the Advanced Normalization Toolkit (ANTs) (Avants et al., 2008). Next, images were aligned to the CITI168 atlas (Pauli et al., 2018) through affine registration performed with NiftyReg (Modat et al., 2010). This alignment was refined by combining it with a precomputed transform from the CITI168 atlas to an oblique orientation along the hippocampus’s long axis, using Convert3D (Yushkevich et al., 2006).

After alignment, the images were resampled to a voxel size of 0.3 mm³ and cropped to dimensions of 128 × 256 × 128 voxels, centering on the left and right hippocampi of the CITI168 atlas. To ensure consistency, left hippocampi were sagittally flipped to match the orientation of right hippocampi. HippUnfold metrics were Z-standardized to facilitate comparison. Volumetric measurements were available for SRLM and Cyst, while thickness metrics were not generated for DG.

### 2.5 Residuals

We controlled for estimated-total intracranial volume (eTIV), age, sex, education level, and scanner-site. eTIV was calculated by utilizing Fastsurfer v.2.12., a segmentation and volume pipeline (Faber et al., 2022; Henschel et al., 2020; Henschel et al., 2022). Parcellation of eTIV via total white and grey matter was obtained by using the Desikan-Killianny-Tourville (DKT) (Klein & Tourville, 2012) protocol generated by Fastsurfer; the volume metric of the parcellation was then calculated using FSLstats in FSL (Woolrich et al., 2009).

### 2.6 Statistical Analyses

We first conducted statistical analyses to explore the differences between groups. To determine if the observed differences in age, education, cognitive scores, and study followup-years among the diagnostic groups were statistically significant, we performed a series of one-way ANOVAs. We examined biological sex separately as a Chi-Square test as a categorical variable.

Statistical analysis was conducted in “R” v.4.3.2 (R Core Team, 2021). We used a linear mixed effects model with the package lme4 to investigate the hierarchical mixed effects of hippocampal morphometry in participants over time. Due to convergence and singularity issues--likely stemming from the complexity of the nested response structure and the volume of data--we chose to focus on specific combinations of hemisphere, hippocampal subfield, and anatomical metric, rather than attempting a single comprehensive model. For each combination, a separate model was implemented using the data subset accordingly:

*Residualized Hippocampus Measure ∼ Progression Group * Followup Years + Covariates (1 + Followup Years | Participant ID)*

By using this subsetting strategy, we ensured reliable parameter estimates, allowing us to draw meaningful conclusions regarding the effects of hippocampal morphometry on cognitive scores. This approach resulted in 50 distinct models being implemented, corrected for multiple comparisons.

A trajectory plot (Figure 1) was generated to visualize changes over time for each hemisphere, hippocampal label, and metric (volume, curvature, gyrification, thickness). Data was grouped by diagnosis and linear models with the interaction term diagnosis and follow-up years were fitted to estimate slopes of over time. Post-hoc pairwise comparisons of slopes were conducted using the *emmeans* package, with corrections of multiple comparisons with the False Discovery Rate (FDR) approach using the p.adjust function in R, corresponding to the Benjamini-Hochberg procedure. (Benjamini & Hochberg, 1995) Significant results were noted in the analysis.

**Figure 1:**
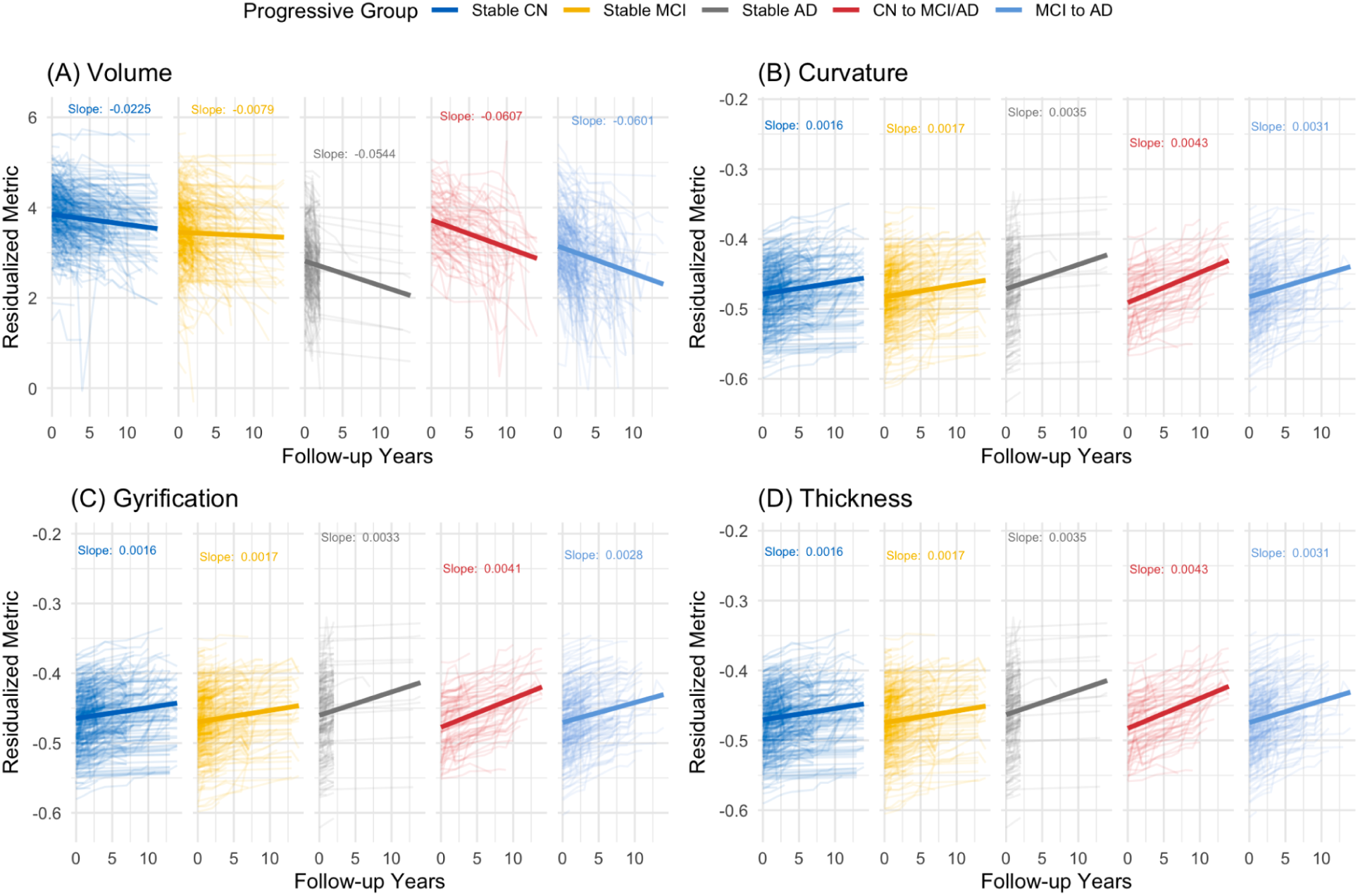
Longitudinal Change in Left CA1 Morphometry for Different Diagnostic Groups. *Note*: Residualized surface-based morphometry (SBM) change over time across four metrics: (A) Volume, (B) Curvature, (C) Gyrification, and (D) Thickness. Trajectories for five progressive groups—Stable CN, Stable MCI, Stable AD, Progressive CN to MCI/AD, and Progressive MCI to AD—are shown. Solid lines represent best-fit slopes, while shaded paths depict individual trajectories. Slope values, reported to four decimal places for clarity, may appear identical despite underlying differences. Pairwise comparisons reveal significant differences in volume across all groups (p < 0.001), and between CN and AD, MCI and AD, and transitions from CN to MCI/AD and MCI/AD for other metrics (p < 0.001).

**Table 2:** Breakdown of Hippocampal Subfield Models by Anatomical Measure and Hemisphere.

| Subfield | Measures | Hemisphere | Total Models |
| --- | --- | --- | --- |
| CA1-CA4, Sub | Volume, Curvature, Gyrification, Thickness | L, R | 40 |
| DG | Volume, Curvature, Gyrification | L, R | 6 |
| Cyst, SRLM | Volume only | L, R | 4 |

*Note.* The model structure includes 8 hippocampal subfields (CA1, CA2, CA3, CA4, DG, Subiculum, Cyst, SRLM) analyzed across 4 anatomical measures (volume, curvature, gyrification, thickness) where applicable. Each subfield-measure combination was analyzed separately for the left (L) and right (R) hemispheres. Notably, Cyst and SRLM subfields only have volume measurements, and DG excludes thickness. This breakdown results in a total of 50 unique models.

The diagnostic group Stable CN was set as the reference level for the intercept and group level effects. This model also provided us with participant level variance of slopes and interaction with timepoints. The model was estimated using Restricted Maximum Likelihood (REML), a method that provides accurate estimates of variance components by accounting for the fixed effects, along with the Bound Optimization By Quadratic Approximation (BOBYQA) optimizer, which ensures the model efficiently converges even in complex arrangements of data where traditional methods may struggle. This approach was used to predict standardized and residualized modality values with diagnostic groups and follow-up years. Then we looked at how random effects from the models accounted for individual variability.

For the time-dependent interactions, we transition to visual representations of among diagnosis groups, focusing on how the average slopes representing changes over time are spatially distributed across various hippocampal regions using Connectome Workbench (Van Essen & The Human Connectome Project, 2012). We assess the interaction effects between Follow-up Years and the diagnosis groups. These interactions reveal how the slopes for different diagnosis groups diverge, highlighting the temporal dynamics associated with the progression of diagnosis.

Afterwards, we ran linear models to assess how the metrics of morphometry can predict changes in cognitive composite scores. Afterwards we ran sensitivity analyses due to differences of MRI tesla strengths and high variability in our PCA analysis.

To control for multiple comparisons, we applied the FDR adjustment^6^. To predict cognitive scores, we fitted linear regression models for each particular hemisphere, metric correspondant to a hippocampal subfield with the slope and progressive change group as predictors for each cognitive domain. The general formula for each regression model was:

*Cognitive Score ∼ Morphometry * Diagnostic Group.*

## 3 Results

### 3.1 Descriptive Statistics

In addition to the one-way ANOVAs conducted for continuous variables, a chi-square test was performed to examine the association between diagnostic group and sex. The results showed a significant association between sex and diagnostic group, χ²(4, N =1318) = [2158.3], [p < .001].^7^

### 3.2 Main Findings

The analysis of hippocampal subregions revealed several broad trends across different metrics. Figure 1 illustrates a general pattern where decreases in volume are often accompanied by increases in SBM metrics. This relationship is particularly evident in individuals diagnosed with Stable AD and progressive groups when compared to the Stable CN controls. This pattern of decreased volume but increased curvature, gyrification and thickness are evident in bilateral CA1, Sub, and the DG.

Interestingly, a distinct pattern emerged in the volume and SBM metrics across hippocampal subregions. While volumetric measures in bilateral CA2 and CA3 regions remained relatively stable across all cohorts, SBM metrics such as curvature, gyrification, and thickness showed consistent *increases*, suggesting shifts in surface topology despite stable volume. A similar trend was observed in the CA4 subregion, where slight volumetric decreases were accompanied by upward trends in other SBM metrics. These findings highlight the complementary insights provided by SBM metrics, particularly in capturing morphological changes that volume alone may not detect.

Moreover, divergent trends were noted in the left Sub, where a decrease in volume was observed alongside a relatively flat curvature slope. These patterns underscore the heterogeneous nature of hippocampal subfield changes and the importance of integrating volumetric and SBM metrics to comprehensively assess structural changes associated with cognitive decline.

While the visual patterns suggest overlapping trajectories, the results from our linear mixed effect models provide additional insight into these changes. Specifically, the models reveal significant differences in the effect sizes between the diagnostic groups. In particular, the comparison between Stable CN and Stable AD groups shows clear distinctions in the rates of structural change.

### 3.3 Fixed Effects

Table 3 presents the fixed effects of the diagnostic group on hippocampal metrics across subregions, reporting findings that remained significant after false discovery rate (FDR) correction. While all comparisons were subjected to FDR adjustment, the effects described here reflect the most robust surviving results. Individuals with stable AD exhibited significant volume reductions and increases in SBM metrics compared to CN individuals. The largest volume reduction was observed in L CA1 (β = −1.01, p < .001), followed by SRLM and Sub, indicating substantial atrophy in these regions. Curvature showed the largest effect sizes for SBM metrics, ranging up to .006 for CA1 curvature for Stable AD (p < 0.01). In contrast, gyrification had the smallest estimates among the SBM metrics, and insignificant in both Stable MCI and AD across CA1, CA3, CA4, and the DG.

**Table 3:** Summary of One-Way ANOVA Results Across Demographic and Cognitive Variables by Diagnostic Group.

| Variable | ANOVA Results<sup>a</sup> | Overall Effect Size ($\eta^2$) | Post-Hoc Comparisons<sup>b</sup> |
| --- | --- | --- | --- |
| Age | $F(4, 4612)=28.02, p < .001$ | 0.02 | Progressive CN to MCI/AD vs Stable MCI (diff = 3.88) |
| Education | $F(4, 4612)=24.55, p < .001$ | 0.02 | Stable CN versus Stable AD (diff = 1.33) |
| Followup Years | $F(4, 4612)=298.5, p < .001$ | 0.21 | Progressive CN to MCI/AD vs Stable AD (diff = 7.18) |
| PHC_MEM | $F(4, 4612)=1570, p < .001$ | 0.58 | Stable AD versus Stable CN (diff = -2.10) |

<sup>6</sup> Among the analyzed models, 72% of results for the fixed effects of diagnosis, including the intercept, met significance criteria ( $p < 0.05$ ) after applying the FDR correction. The significance ratios are as follows: Intercept (Stable CN) at 100%, Stable MCI at 78%, CN to MCI/AD at 78%, Stable AD 70%, and Progressive MCI to AD at 32%. The interactions of diagnosis and follow-up years yielded in total 67.6% and for the following terms: MCI to AD at 94%, CN to MCI/AD at 84%, Stable AD at 82%, Follow-Up Years at 70%, and Stable MCI at 8%.

|  |  |  |  |
| --- | --- | --- | --- |
| PHC_VSP | $F(4, 2665)=76.64, p < .001$ | 0.10 | Stable AD vs Stable CN (diff = -0.93) |
| PHC_LAN | $F(4, 4606)=658.6, p < .001$ | 0.36 | Stable CN vs Stable AD (diff = 1.74) |
| PHC_EXF | $F(4, 4587)=622.4, p < .001$ | 0.35 | Stable CN vs Stable AD (diff = 1.73) |

*Note.* The table presents the results of one-way ANOVA tests across various demographic and cognitive variables by diagnostic group. The "Post-Hoc Comparisons" column highlights the largest absolute differences between diagnostic groups with Tukey Honest Significant Difference test, showing which groups had the most substantial contrast.
<sup>a</sup> The ANOVA was performed on complete cases, meaning that only participants with available neuropsychological measures were included in the analysis.
<sup>b</sup> The largest absolute difference for each variable is reported. All post-hoc comparisons are significant ( $p < .001$ ), except for the Age comparison between Progressive MCI to AD vs Stable AD ( $p = .110$ ), and Education comparison between Stable MCI and Progressive MCI to AD ( $p = .997$ ).

**Table 4:**
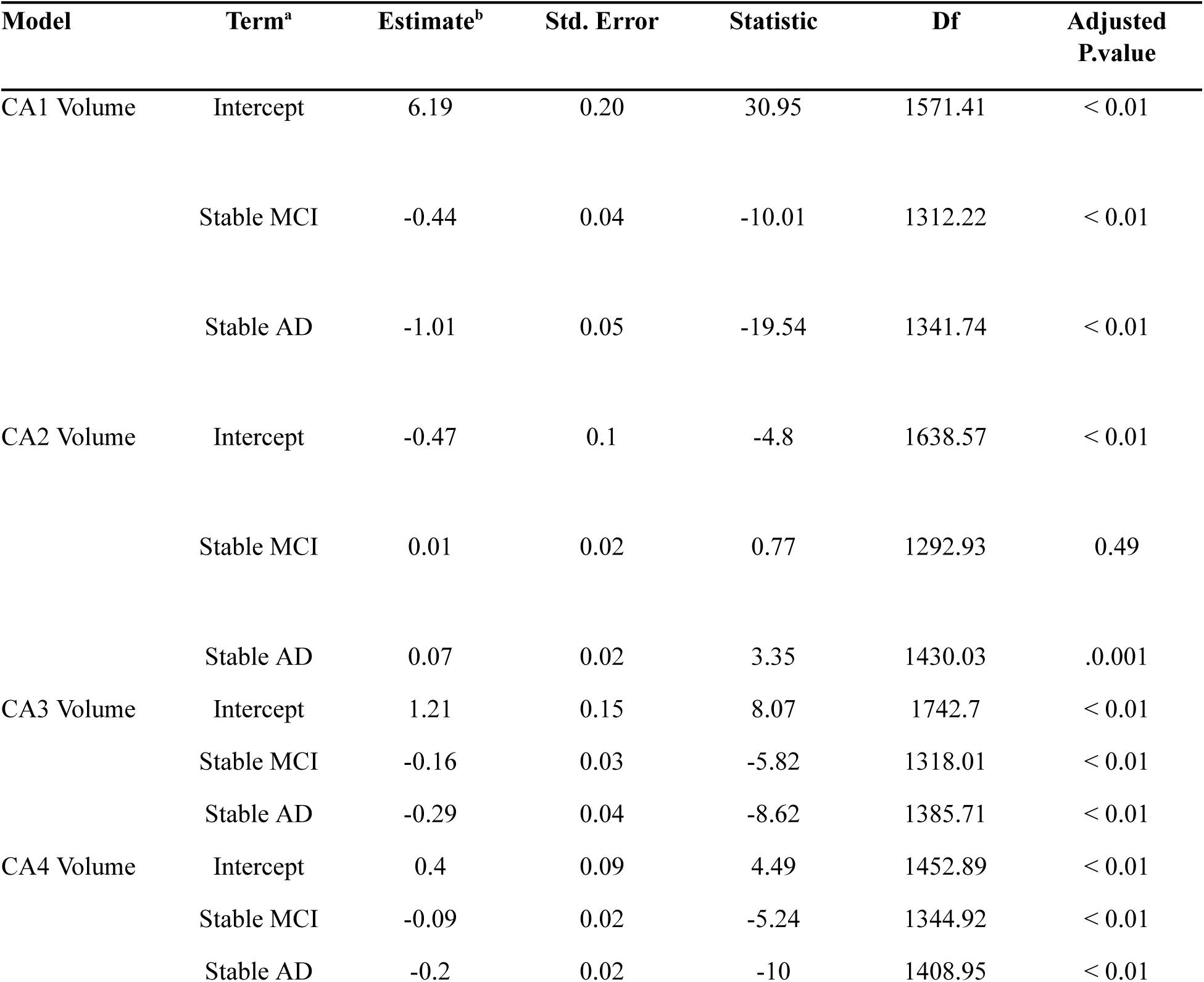
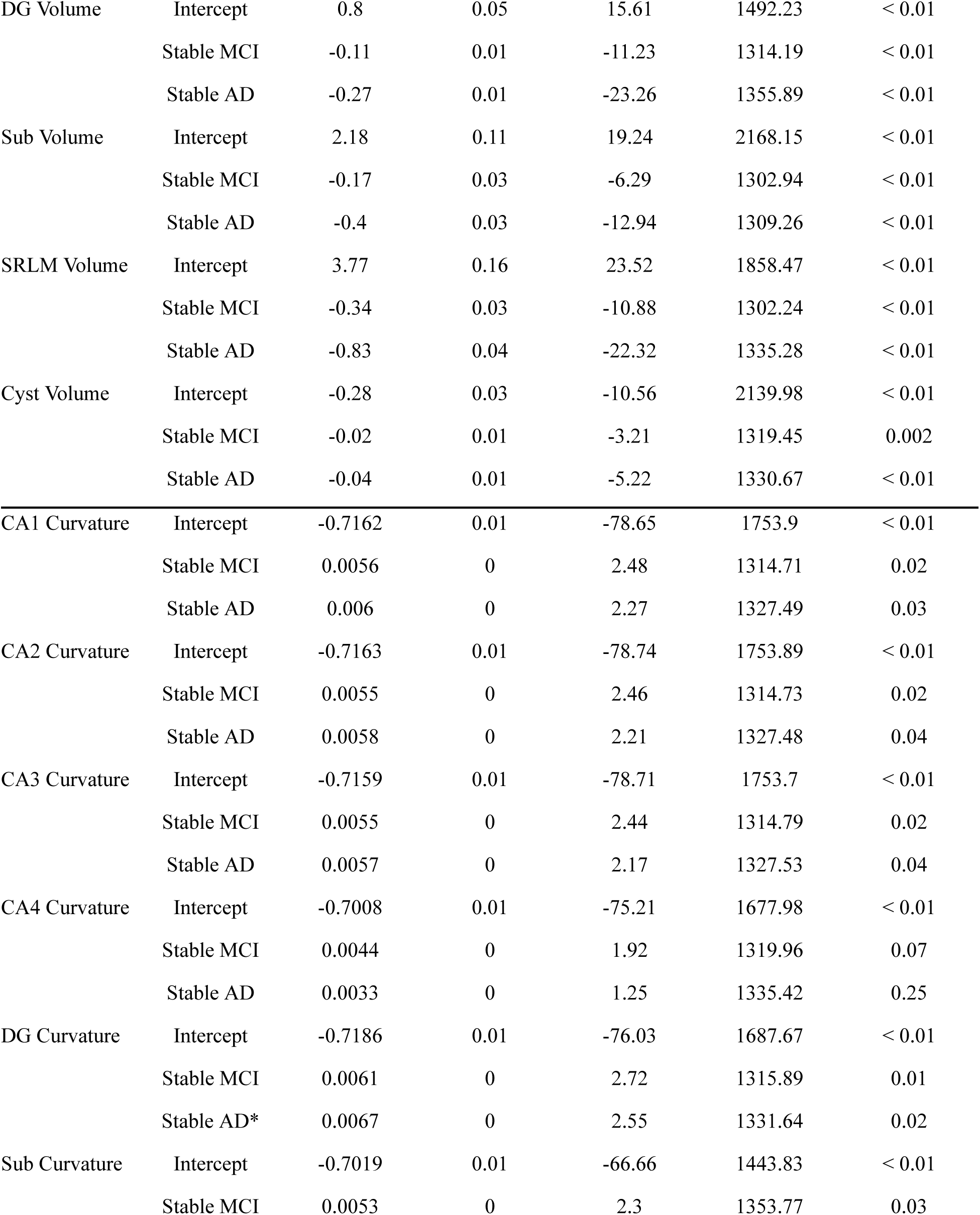
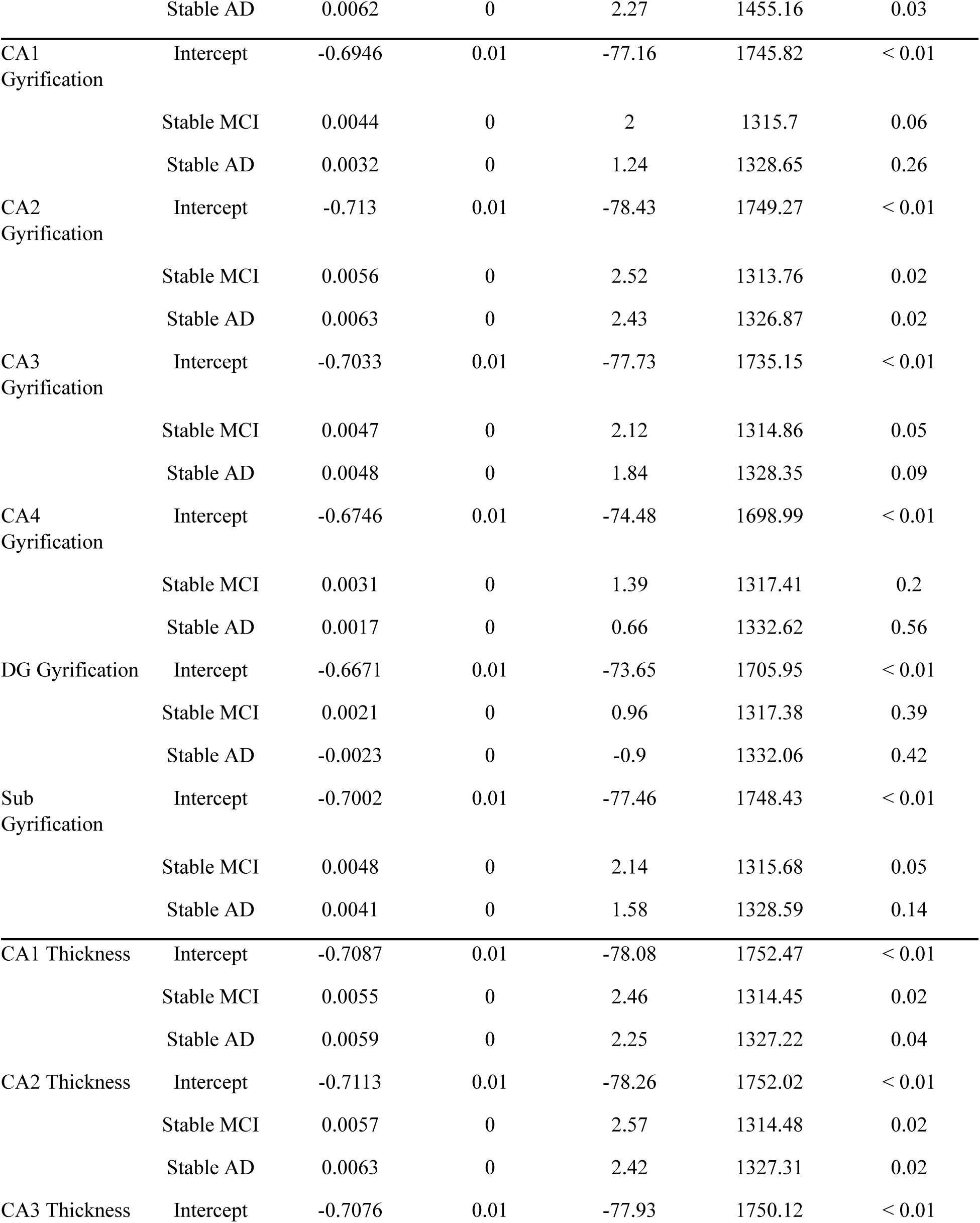
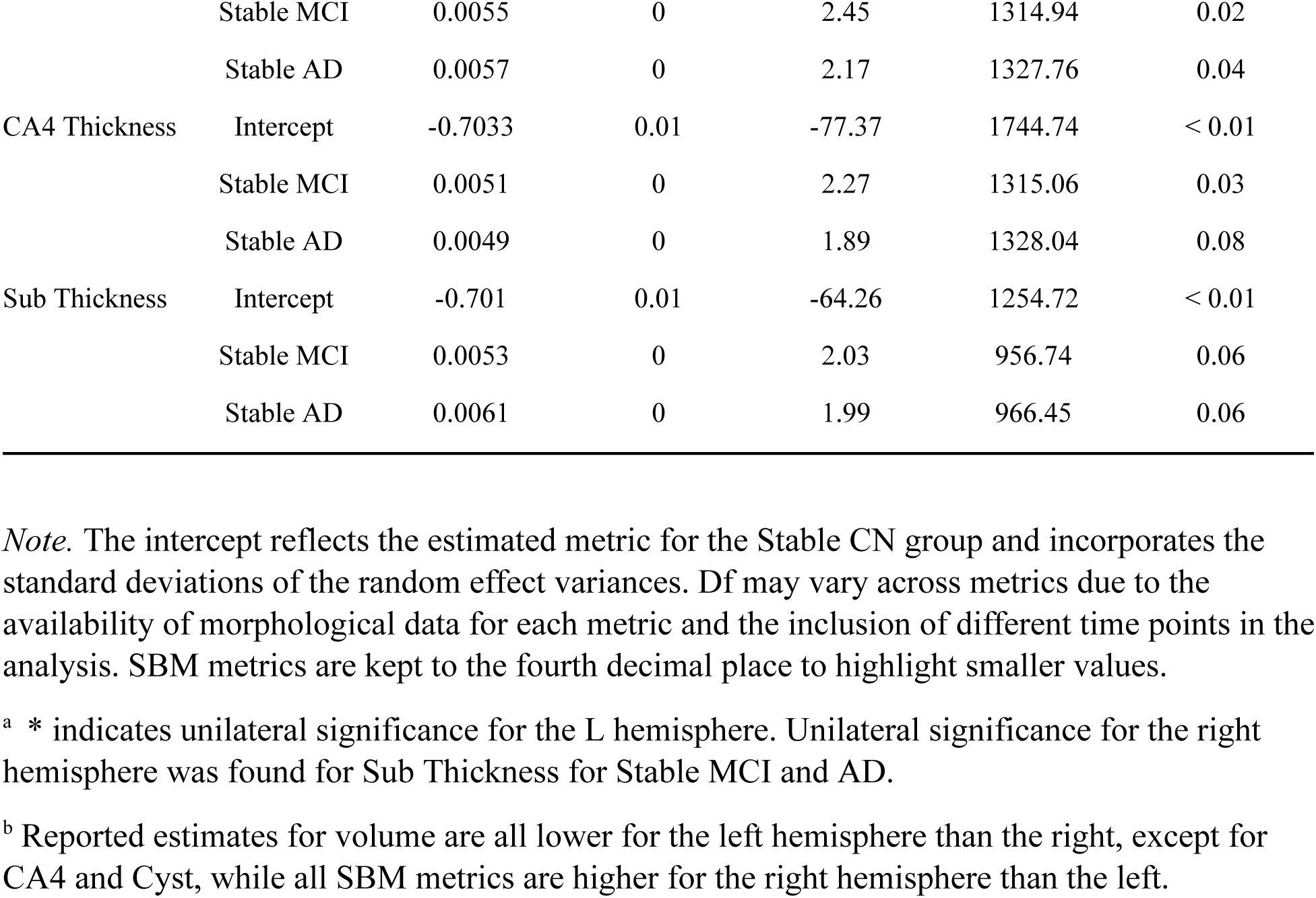
Fixed Effects of Diagnostic Status on Hippocampal Metrics Across Subregions.

The covariates included in the models were significant for most metrics, with exceptions primarily in volume and L Sub thickness. Age was a significant predictor except for volume in L CA3 and L CA4. Education showed significance across all models except for volume in bilateral Cysts. Scanner site had no significant effects on any volume metric or L Sub thickness. Sex was not significant for volume in L CA2, bilateral CA4, and L DG.

The random intercept standard deviation ranged from 0.60 to 0.03, with volume measures showing the highest variances and gyrification exhibiting the lowest. Random slope standard deviation variances ranged from 0.04 to 0.004, reflecting a similar trend, with volume metrics exhibiting higher variability. The correlation between the intercept and slope ranged from 0.04 to 0.64, with volume metrics showing the strongest correlation and curvature and gyrification exhibiting the weakest. Residual standard deviations ranged from −0.4 to −5.70E-5, with volume metrics displaying the highest residual variance and curvature metrics showing the lowest.

### 3.4 Slopes of Main Effects and Interaction Effects

Table 5 indicates the significance of interactions retained by multiple comparisons, with the CA1 region serving as an exemplar, as the pattern of effects of the interaction terms are consistent with other significant interactions; see supplementary materials for full data. The trend of effects of volume decreasing and SBM metrics rising are consistent with the fixed effects in the models. For the volume metrics, reductions in volume had greatest negative effects for the MCI to AD group. For example, CA1 was significant in both the Stable AD group (β = −0.05) and in the transition from CN to MCI/AD (β = −0.03), yet MCI to AD showed (β = −0.06). However, significant increases were observed in CA2 for Stable AD (β = 0.04), and in CA3 for MCI to AD (β = .03). Reductions in the Sub, DG, SRLM, and Cyst regions were found in the comparisons between CN and MCI/AD, as well as in the MCI to AD group. SBM metrics showcased higher increases for the MCI to AD group in comparison to the Stable AD group. Whereas gyrification was not significant in the main effects for Stable AD in a number of subfields, the interaction for time shows significance.

**Table 5:** Interactions of Follow-up Years and Diagnostic Group Surviving Multiple Comparisons.

| Model | Term | Estimate | Std. Error | Statistic | Df | Adj. P. value<sup>a</sup> |
| --- | --- | --- | --- | --- | --- | --- |
| Volume | Follow-Up Years | -0.01 | 0.00 | -2.56 | 419.78 | 0.02 |
|  | Follow-Up<br>Years:Stable AD | -0.05 | 0.01 | -4.08 | 655.96 | < 0.01 |
|  | Follow-Up<br>Years:CN to<br>MCI/AD | -0.03 | 0.00 | -3.49 | 295.12 | < 0.01 |
|  | Follow-Up<br>Years:MCI to<br>AD | -0.06 | 0.00 | -9.54 | 475.33 | < 0.01 |
| Curvature | Follow-Up Years | 0.0003 | 0.00 | 2.59 | 705.97 | 0.02 |
|  | Followup-Years:<br>Stable AD | 0.0010 | 0.00 | 2.74 | 895.80 | 0.01 |
|  | Follow-Up<br>Years:CN to<br>MCI/AD | 0.0010 | 0.00 | 4.20 | 503.15 | < 0.01 |
|  | Follow-Up<br>Years:MCI to<br>AD | 0.0012 | 0.00 | 6.00 | 768.69 | < 0.01 |
| Gyrification | Follow-Up Years | 0.0003 | 0.00 | 2.52 | 709.06 | 0.02 |
|  | Follow-Up<br>Years:Stable AD | 0.0009 | 0.00 | 2.39 | 899.30 | 0.02 |
|  | Follow-Up<br>Years:CN to<br>MCI/AD | 0.0009 | 0.00 | 3.78 | 507.62 | 0.00 |
|  | Follow-Up<br>Years:MCI to<br>AD | 0.0010 | 0.00 | 4.75 | 773.71 | < 0.01 |
| Thickness | Follow-Up Years | 0.0003 | 0.00 | 2.46 | 703.37 | 0.02 |
|  | Follow-Up<br>Years:Stable AD | 0.0011 | 0.00 | 2.94 | 888.05 | 0.01 |
|  | Follow-Up<br>Years:CN to<br>MCI/AD | 0.0010 | 0.00 | 4.17 | 501.11 | < 0.01 |
|  | Follow-Up<br>Years:MCI to<br>AD | 0.0013 | 0.00 | 6.27 | 766.97 | < 0.01 |

*Note.* This table presents significant interaction effects between diagnostic groups for volume, and SBM metrics across hippocampal subregions, with p-values adjusted for multiple comparisons using the False Discovery Rate (FDR) method. SBM metrics are kept to the fourth decimal place to highlight smaller values.

Figure 2 illustrates the average slopes within each metric across diagnostic groups. For volume, CA1 exhibits the lowest slopes, while CA2 shows the highest. Curvature varies by hemisphere, with DG having the highest slopes in the left hemisphere but among the lowest in the right, while CA4 shows the opposite pattern. For gyrification, DG has the lowest slopes, whereas CA2 has the highest. In thickness, Sub displays some of the highest slopes, while CA4 has the lowest.

**Figure 2:**
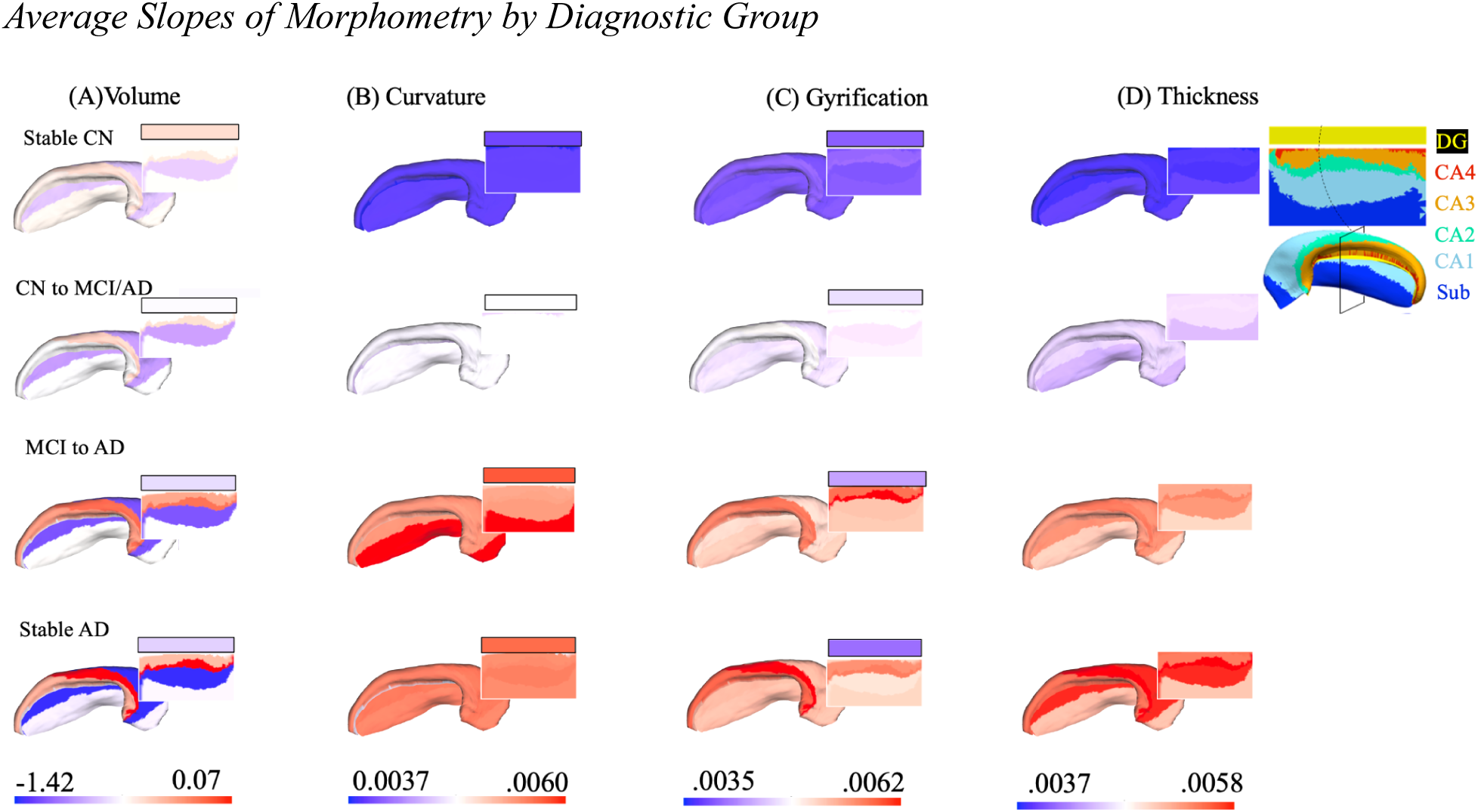
Average Slopes of Morphometry by Diagnostic Group. *Note*. (A)–(D) display left dorsal and unfolded hippocampal maps from Connectome Workbench, using label files with RGB values based on averaged slopes by metric, hemisphere, and diagnostic group, ordered by slope severity. Colors represent *relative* slope differences, with blue, white, and red indicating the lowest, median, and highest averages. See supplementary section for bilateral slope comparison.

### 3.5 Effects on Cognition

We applied FDR adjustment for multiple comparisons to control for Type I errors. Overall, only 22.8% of results for interactions survived comparisons for significance. Our analysis revealed that among the models that reached statistical significance, memory is the predominant cognitive domain for the metrics that served as predictors, with visuospatial processing being the lowest.

Figure 3 illustrates the range of effect sizes from these interactions. Interestingly, while volume was observed to have minimal effect sizes on cognition, the positive SBM metrics predicted positive effect sizes on cognition for the Stable AD group.

**Figure 3:**
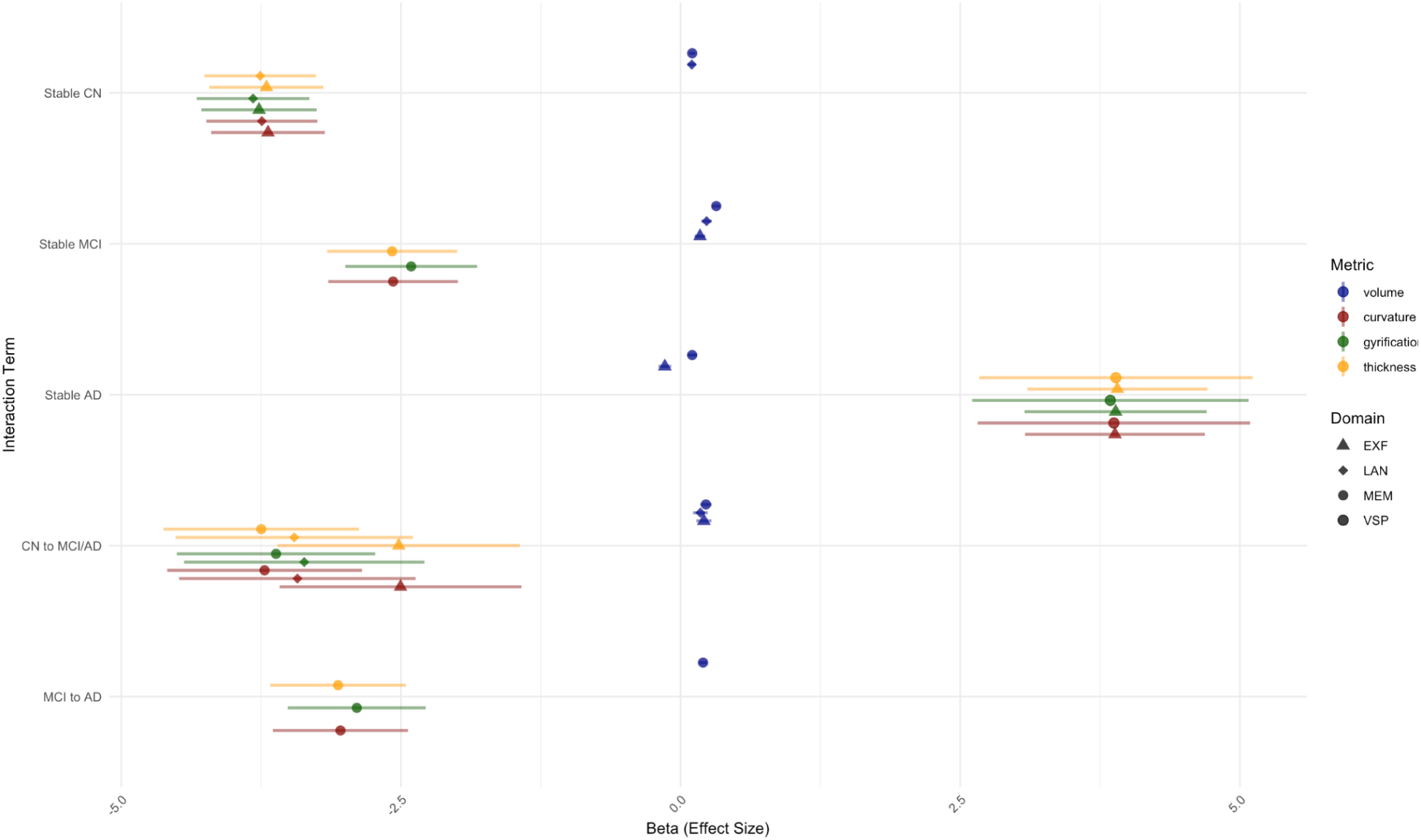
Effect Sizes for Interactions of Morphometry with Diagnosis. *Note.* This point-range plot of the left CA1 presents the interaction effects of diagnosis and slope on cognitive domains, executive functioning (EXF), language processing (LAN), and memory (MEM). Each panel corresponds to a specific subfield for the left hemisphere, with the point-ranges indicating the magnitude of the effects and standard error.

### 3.6 Sensitivity Analyses

We found more significant findings in the 3.0T than the 1.5T data. In the 3.0T data, more significant findings were identified for curvature, gyrification, and thickness, whereas the 1.5T data showed more significant effects on volumetric measures. A subsequent analyses of 1.5 and 3.0T data collectively showed that there is a general trend that a majority of the findings share significant findings that are reported in our main analysis.

However, there are some divergences. Specifically, the 1.5T data alone picked up decreases in effect sizes of CA1 volumes for time interactions from CN to MCI/AD, and Stable MCI; whereas for volume for the CA2 and CA3 regions there were increases of effect sizes than the 3.0 scans.

## 4 Discussion

Our analyses have uncovered several noteworthy findings regarding hippocampal and cortical metrics in AD. As hypothesized, we observed greater negative effect sizes for volume reductions in AD, particularly in the CA1 and subiculum subregions. This is consistent with prior research and reinforces the role of these regions as biomarkers in AD progression. These findings align with Mak et al. (2016), who reported significant atrophy in this area, and corroborates earlier studies by Apostolova et al. (2006), La Joie et al. (2013), and Zhao et al. (2019), which highlighted the CA1 and subiculum as critical sites of neurodegeneration in AD. These findings reinforce the importance of these regions as biomarkers of AD progression.

However, contrary to our expectation, the decrease of cortical thickness was not confirmed. Instead of the anticipated cortical thinning in AD, we observed an increase in cortical thickness. This result parallels findings from Phan et al. (2024), who noted an anticorrelation between cortical thickness and brain atrophy, particularly in the anterior cingulate cortex (ACC). Possible explanations for this phenomenon include neuroplasticity and neuroinflammation. The increased cortical thickness could reflect adaptive neuroplastic changes or inflammatory responses altering cortical architecture. Another study utilizing HippUnfold examining age-related decline in thickness and surface area in the hippocampus reveals that there are patterns of decline in cortical thinning that are mostly linear, but are highly heterogeneous (Yu et al, 2024). Cortical thickening has also been reported in early preclinical AD (Montal et al., 2018), as well as in normal tau protein accumulation in AD cohorts (Hojjati et al., 2023).

We did not hypothesize an increase in gyrification that we observed in AD. This finding is reminiscent of recent work by Núñez et al. (2020), which found increased gyrification in the entorhinal cortex in AD. This unexpected finding may be indicative of a compensatory response to white matter (WM) atrophy or neurogenesis in specific cortical areas. However, this hypothesis remains controversial, and further research is required to better understand the underlying mechanisms.

Similarly, we also did not hypothesize an increase in cortical curvature, yet the increase in cortical curvature we documented may be a necessary correlate of the increased gyrification. This suggests a complex relationship between cortical folding and overall brain morphology in AD. The interplay between gyrification and curvature is an area that warrants further investigation to understand the adaptive or maladaptive changes occurring in AD. One possible explanation is the phenomenon of white matter pulling away from grey matter. As WM atrophy occurs in AD, the reduced volume of white matter may lead to a “stretching” effect on the overlying cortical gray matter. This could increase gyrification as the cortical surface is stretched in response to the shrinking underlying white matter.

In summary, while the volume reductions in the HC align with expected patterns of neurodegeneration in AD, the observed increases in gyrification, cortical thickness, and curvature present novel insights into the disease’s impact on brain morphology, or to be more precise, morphology in the HC. These findings suggest both adaptive and pathological changes and highlight the need for further research to elucidate their mechanisms and implications for AD understanding and diagnosis.

Our sensitivity analysis replicates previous findings that 3.0T is more sensitive to effect sizes and significant differences in neurodegeneration (Chow et al., 2015; Leandrou et al., 2020). Although general trends are found within both field strengths, mixing data of these strengths have been shown to not drastically affect bias or variation between participants (Ho et al., 2010).

## 5 Limitations

While our findings are substantive, there are several limitations. First, the ADNI dataset lacks full harmonization to account for neuropsychiatric differences across its phases. Additionally, HippUnfold’s subfield definitions, based on 3DBigBrain (DeKraker et al., 2020), differ slightly from traditional segmentation boundaries. For example, recent work by Hickling et al. (2024) shows CA4 associated with DG, and the Sub region divided into two substrates. Another limitation is the time gap between MRI scans and neuropsychological assessments; despite allowing a 180-day cutoff, closer tracking would provide a more accurate link between brain states and neuropsychiatric measures.

Though HippUnfold represents an advanced deep-learning tool for hippocampal subfield segmentation, it has limitations, particularly regarding validation against manual segmentation. Similar to concerns raised with FreeSurfer, HippUnfold’s subfield classification may be less precise without high-resolution T2 data, and it has shown a tendency to overestimate subfield volumes (Sghirripa et al., 2024). To mitigate volume measurement inaccuracies, we recommend including additional surface parameters in future analyses.

## 6 Conclusion

In summary, our findings illuminate the complex interplay between structural changes in the HC and cognitive decline in AD. We discovered that both Stable CN and AD groups exhibited significant reductions in hippocampal volume, yet paradoxically, they demonstrated increased SBM effects, suggesting a nuanced adaptation in the brain’s architecture as it grapples with the onset of dementia.

The significance of time interactions revealed that individuals transitioning from CN to MCI/AD and MCI to AD displayed particularly pronounced effects, reinforcing the notion that early interventions could be crucial in mitigating the trajectory of cognitive decline. Notably, language and executive functioning emerged as a powerful predictor in our linear regression analyses for SBM metrics, eclipsing the influence of volume, although the association between volume loss and memory decline in AD remained strong and undeniable.

These results not only deepen our understanding of the morphological transformations occurring in the hippocampus as AD progresses but also underscore the necessity of examining surface-based parameters alongside traditional volumetric measures. As we look to the future, our work paves the way for broader investigations into the intricate relationships between hippocampal morphology, cognitive functions, and emotional health, offering hope for more comprehensive approaches to diagnosing and treating cognitive decline in AD.

## Supporting information

All Supplementary Material

## Data Availability

All data produced in the present study are available upon reasonable request to the authors.

http://www.loni.ucla.edu/ADNI

https://github.com/salahaziz94/ADNI_HippUnfold

## Acknowledgements

Data collection and sharing for this project was funded by the Alzheimer’s Disease Neuroimaging Initiative (ADNI) (National Institutes of Health Grant U01 AG024904) and DOD ADNI (Department of Defense award number W81XWH-12-2-0012). ADNI is funded by the National Institute on Aging, the National Institute of Biomedical Imaging and Bioengineering, and through generous contributions from the following: AbbVie, Alzheimer’s Association; Alzheimer’s Drug Discovery Foundation; Araclon Biotech; BioClinica, Inc.; Biogen; Bristol-Myers Squibb Company; CereSpir, Inc.; Cogstate; Eisai Inc.; Elan Pharmaceuticals, Inc.; Eli Lilly andCompany; EuroImmun; F. Hoffmann-La Roche Ltd and its affiliated company Genentech, Inc.; Fujirebio; GE Healthcare; IXICO Ltd.; Janssen Alzheimer Immunotherapy Research & Development, LLC.; Johnson & Johnson Pharmaceutical Research & Development LLC.; Lumosity; Lundbeck; Merck & Co., Inc.; Meso Scale Diagnostics, LLC.; NeuroRx Research; Neurotrack Technologies; Novartis Pharmaceuticals Corporation; Pfizer Inc.; Piramal Imaging; Servier; Takeda Pharmaceutical Company; and Transition Therapeutics. The Canadian Institutes of Health Research is providing funds to support ADNI clinical sites in Canada. Private sector contributions are facilitated by the Foundation for the National Institutes of Health (www.fnih.org). The grantee organization is the Northern California Institute for Research and Education, and the study is coordinated by the Alzheimer’s Therapeutic Research Institute at the University of Southern California. ADNI data are disseminated by the Laboratory for Neuro Imaging at the University of Southern California.

## Author Note

We have no known conflict of interest to disclose. This project was supported by an NSERC Discovery Grant (DGECR-2022-00309) and a Canada Research Chair (Tier II, CRC-2020-00174) to JAEA.We have no known conflict of interest to disclose. This project was supported by an NSERC Discovery Grant (DGECR-2022-00309) and a Canada Research Chair (Tier II, CRC-2020-00174) to JAEA.

## CRediT Statement

SA: Conceptualization, Data curation, Formal analysis, Investigation, Methodology, Resources, Software, Validation, Visualization, Writing – original draft, Writing – review and editing

RP: Conceptualization, Investigation, Methodology, Software, Supervision, Validation, Writing – review and editing

CM: Investigation, Methodology, Validation, Visualization, Writing - review and editing

PZ: Investigation, Methodology, Validation, Visualization, Writing – review and editing

JAE: Conceptualization, Data curation, Funding acquisition, Investigation, Methodology, Project administration, Resources, Software, Supervision, Validation, Visualization, Writing – review and editing

## Footnotes

6 Among the analyzed models, 72% of results for the fixed effects of diagnosis, including the intercept, met significance criteria (p < 0.05) after applying the FDR correction. The significance ratios are as follows: Intercept (Stable CN) at 100%, Stable MCI at 78%, CN to MCI/AD at 78%, Stable AD 70%, and Progressive MCI to AD at 32%. The interactions of diagnosis and follow-up years yielded in total 67.6% and for the following terms: MCI to AD at 94%, CN to MCI/AD at 84%, Stable AD at 82%, Follow-Up Years at 70%, and Stable MCI at 8%.

7 Standardized residuals from the Chi-Square test showed significant gender distribution differences. In the Stable CN group, females had a residual of 41.99 (higher than expected), and males had −41.99 (lower than expected). In the Stable MCI group, females had −29.23 and males 29.23. The Stable AD group had minimal deviations (females: 5.26; males: −5.26). For the CN to MCI/AD transition, females had 9.09 and males −9.09. In the MCI to AD group, females had −25.33 and males 25.33.

