## Supplementary material for "Longitudinal Surface-Based Morphometry Changes in the Hippocampus in Dementia": All Supplementary Material

#### Figure S1

*Longitudinal Change in Left CA3 Morphometry for Different Diagnostic Groups*

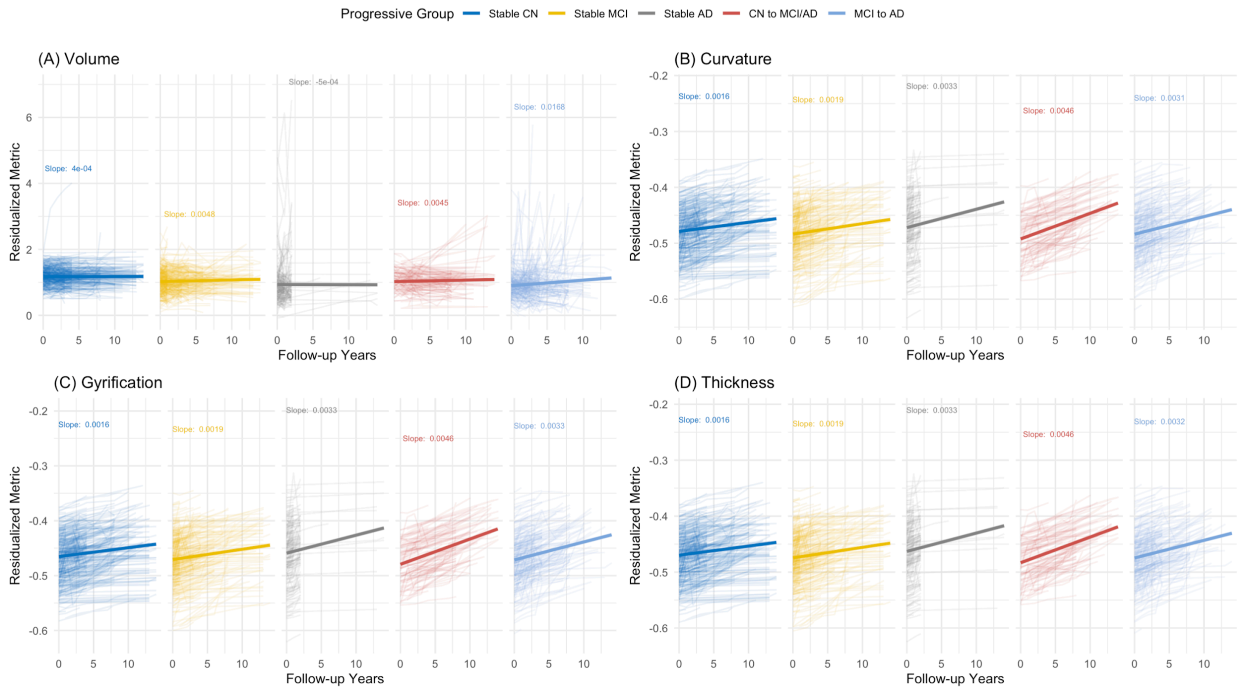


#### Figure S2

*Longitudinal Change in Left Subiculum Morphometry for Different Diagnostic Groups*

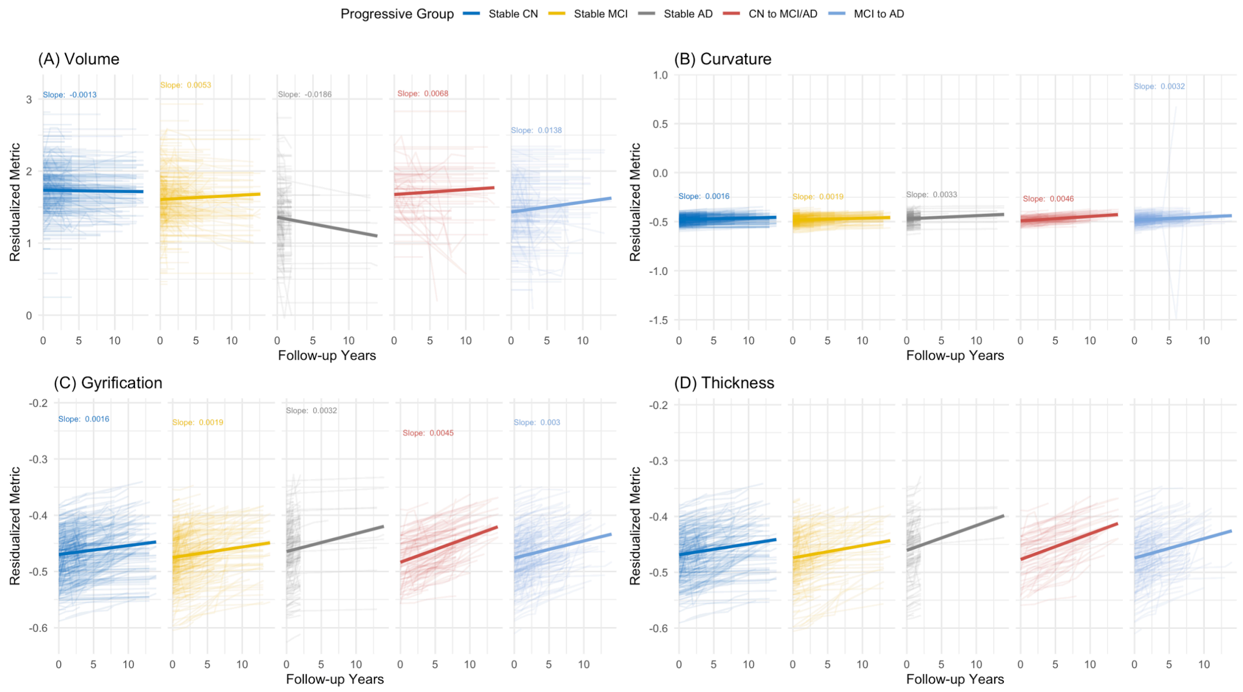


### Figure S3

*Bilateral Slopes of Morphometry on Diagnostic Groups*

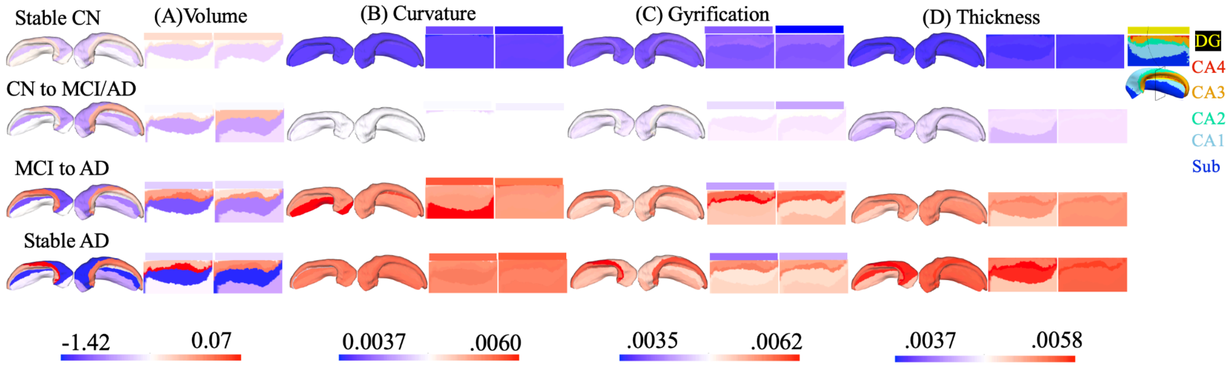


### Figure S4

*Effect Sizes of Interactions of Morphometry with Diagnostic Groups in Left Hemisphere*

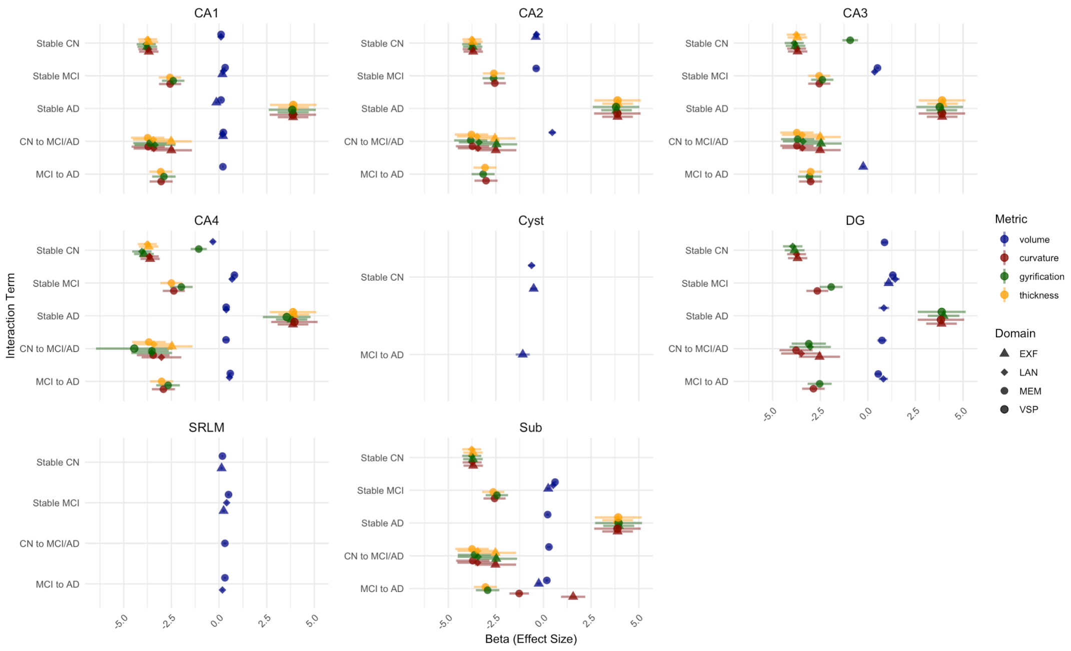


**Table S1**

*All Mixed-Effect Model Results Adjusted for Multiple Comparisons*

| Model | Term | Estimate | Std.Error | Statistic | df | Adj p value |
|---|---|---|---|---|---|---|
| L CA1 volume | (Intercept) | 6.1879 | 0.20 | 30.95 | 1571.41 | < 0.01 |
| L CA1 volume | Stable MCI | -0.4443 | 0.04 | -10.01 | 1312.22 | < 0.01 |
| L CA1 volume | Stable AD | -1.0143 | 0.05 | -19.54 | 1341.74 | < 0.01 |
| L CA1 volume | CN to MCI/AD | -0.1646 | 0.08 | -2.13 | 1242.34 | 0.05 |
| L CA1 volume | MCI to AD | -0.7421 | 0.05 | -14.32 | 1281.16 | < 0.01 |
| L CA1 volume | Followup Years | -0.0102 | 0.00 | -2.56 | 419.78 | 0.02 |
| L CA1 volume | Age | -0.0314 | 0.00 | -13.94 | 1659.59 | < 0.01 |
| L CA1 volume | Education | -0.0024 | 0.01 | -0.39 | 1288.53 | 0.73 |
| L CA1 volume | Scanner Site | 3.43E-05 | 0.00 | 0.24 | 1328.88 | 0.84 |

| | | | | | | |
|---|---|---|---|---|---|---|
| L CA1 volume | SexM | 0.1522 | 0.04 | 4.20 | 1464.25 | < 0.01 |
| L CA1 volume | ETIV | 0.1674 | 0.01 | 13.36 | 4201.14 | < 0.01 |
| L CA1 volume | Stable MCI:Followup Years | -0.0079 | 0.01 | -1.37 | 406.34 | 0.21 |
| L CA1 volume | Stable AD:Followup Years | -0.0477 | 0.01 | -4.08 | 655.96 | < 0.01 |
| L CA1 volume | CN to MCI/AD:Followup Years | -0.0267 | 0.01 | -3.49 | 295.12 | < 0.01 |
| L CA1 volume | MCI to AD:Followup Years | -0.0614 | 0.01 | -9.54 | 475.33 | < 0.01 |
| L CA1 curvature | (Intercept) | -0.7163 | 0.01 | -78.94 | 1764.45 | < 0.01 |
| L CA1 curvature | Stable MCI | 0.0056 | 0.00 | 2.49 | 1322.66 | 0.02 |
| L CA1 curvature | Stable AD | 0.0060 | 0.00 | 2.28 | 1335.55 | 0.03 |
| L CA1 curvature | CN to MCI/AD | -0.0105 | 0.00 | -2.67 | 1290.91 | 0.01 |
| L CA1 curvature | MCI to AD | 0.0010 | 0.00 | 0.40 | 1307.83 | 0.73 |
| L CA1 curvature | Followup Years | 0.0003 | 0.00 | 2.60 | 709.55 | 0.02 |
| L CA1 curvature | Age | 0.0041 | 9.82E-05 | 41.29 | 1835.84 | < 0.01 |
| L CA1 curvature | Education | -0.0029 | 0.00 | -9.70 | 1310.10 | < 0.01 |
| L CA1 curvature | Scanner Site | 0.0000 | 6.95E-06 | -4.60 | 1207.19 | < 0.01 |
| L CA1 curvature | SexM | -0.0368 | 0.00 | -21.90 | 1315.61 | < 0.01 |
| L CA1 curvature | Stable MCI:Followup Years | 0.0003 | 0.00 | 1.76 | 661.59 | 0.10 |
| L CA1 curvature | Stable AD:Followup Years | 0.0010 | 0.00 | 2.74 | 895.39 | 0.01 |
| L CA1 curvature | CN to MCI/AD:Followup Years | 0.0010 | 0.00 | 4.23 | 505.64 | < 0.01 |
| L CA1 curvature | MCI to AD:Followup Years | 0.0012 | 0.00 | 6.02 | 773.82 | < 0.01 |
| L CA1 gyrification | (Intercept) | -0.6946 | 0.01 | -77.44 | 1756.27 | < 0.01 |
| L CA1 gyrification | Stable MCI | 0.0044 | 0.00 | 2.01 | 1323.65 | 0.06 |
| L CA1 gyrification | Stable AD | 0.0032 | 0.00 | 1.25 | 1336.71 | 0.25 |
| L CA1 gyrification | CN to MCI/AD | -0.0107 | 0.00 | -2.75 | 1291.57 | 0.01 |
| L CA1 gyrification | MCI to AD | -0.0007 | 0.00 | -0.27 | 1308.69 | 0.81 |

| | | | | | | |
|---|---|---|---|---|---|---|
| L CA1 gyrification | Followup Years | 0.0003 | 0.00 | 2.53 | 712.69 | 0.02 |
| L CA1 gyrification | Age | 0.0039 | 9.71E-05 | 40.62 | 1824.99 | < 0.01 |
| L CA1 gyrification | Education | -0.0029 | 0.00 | -9.81 | 1310.00 | < 0.01 |
| L CA1 gyrification | Scanner Site | 0.0000 | 6.86E-06 | -4.67 | 1205.12 | < 0.01 |
| L CA1 gyrification | SexM | -0.0362 | 0.00 | -21.77 | 1315.35 | < 0.01 |
| L CA1 gyrification | Stable MCI:Followup Years | 0.0003 | 0.00 | 1.58 | 665.71 | 0.14 |
| L CA1 gyrification | Stable AD:Followup Years | 0.0009 | 0.00 | 2.39 | 898.87 | 0.02 |
| L CA1 gyrification | CN to MCI/AD:Followup Years | 0.0009 | 0.00 | 3.80 | 510.18 | < 0.01 |
| L CA1 gyrification | MCI to AD:Followup Years | 0.0010 | 0.00 | 4.77 | 778.92 | < 0.01 |
| L CA1 thickness | (Intercept) | -0.7088 | 0.01 | -78.36 | 1763.01 | < 0.01 |
| L CA1 thickness | Stable MCI | 0.0055 | 0.00 | 2.47 | 1322.39 | 0.02 |
| L CA1 thickness | Stable AD | 0.0059 | 0.00 | 2.25 | 1335.27 | 0.03 |
| L CA1 thickness | CN to MCI/AD | -0.0105 | 0.00 | -2.69 | 1290.49 | 0.01 |
| L CA1 thickness | MCI to AD | 0.0010 | 0.00 | 0.37 | 1307.49 | 0.75 |
| L CA1 thickness | Followup Years | 0.0003 | 0.00 | 2.47 | 706.94 | 0.02 |
| L CA1 thickness | Age | 0.0041 | 9.78E-05 | 41.50 | 1832.81 | < 0.01 |
| L CA1 thickness | Education | -0.0029 | 0.00 | -9.74 | 1309.67 | < 0.01 |
| L CA1 thickness | Scanner Site | 0.0000 | 6.93E-06 | -4.61 | 1206.44 | < 0.01 |
| L CA1 thickness | SexM | -0.0366 | 0.00 | -21.81 | 1315.17 | < 0.01 |
| L CA1 thickness | Stable MCI:Followup Years | 0.0003 | 0.00 | 1.80 | 659.16 | 0.09 |
| L CA1 thickness | Stable AD:Followup Years | 0.0011 | 0.00 | 2.94 | 887.63 | 0.01 |
| L CA1 thickness | CN to MCI/AD:Followup Years | 0.0010 | 0.00 | 4.20 | 503.58 | < 0.01 |
| L CA1 thickness | MCI to AD:Followup Years | 0.0013 | 0.00 | 6.29 | 772.08 | < 0.01 |
| L CA2 volume | (Intercept) | -0.4206 | 0.08 | -4.98 | 1449.07 | < 0.01 |
| L CA2 volume | Stable MCI | 0.0139 | 0.02 | 0.77 | 1292.93 | 0.49 |

| | | | | | | |
|---|---|---|---|---|---|---|
| L CA2 volume | Stable AD | 0.0715 | 0.02 | 3.35 | 1430.03 | < 0.01 |
| L CA2 volume | CN to MCI/AD | -0.0459 | 0.03 | -1.51 | 1066.54 | 0.16 |
| L CA2 volume | MCI to AD | 0.0197 | 0.02 | 0.95 | 1198.56 | 0.39 |
| L CA2 volume | Followup Years | 0.0007 | 0.00 | 0.26 | 271.78 | 0.82 |
| L CA2 volume | Age | 0.0080 | 0.00 | 8.28 | 1479.80 | < 0.01 |
| L CA2 volume | Education | -0.0016 | 0.00 | -0.65 | 1282.64 | 0.56 |
| L CA2 volume | Scanner Site | 0.0000 | 6.01E-05 | -0.29 | 1448.80 | 0.80 |
| L CA2 volume | SexM | 0.0190 | 0.02 | 1.25 | 1436.40 | 0.25 |
| L CA2 volume | ETIV | 0.0077 | 0.01 | 1.18 | 2235.10 | 0.28 |
| L CA2 volume | Stable MCI:Followup Years | 0.0026 | 0.00 | 0.67 | 281.96 | 0.56 |
| L CA2 volume | Stable AD:Followup Years | 0.0353 | 0.01 | 4.35 | 422.19 | < 0.01 |
| L CA2 volume | CN to MCI/AD:Followup Years | 0.0070 | 0.01 | 1.38 | 195.92 | 0.21 |
| L CA2 volume | MCI to AD:Followup Years | 0.0362 | 0.00 | 8.28 | 341.24 | < 0.01 |
| L CA2 curvature | (Intercept) | -0.7164 | 0.01 | -79.02 | 1764.42 | 0.00 |
| L CA2 curvature | Stable MCI | 0.0055 | 0.00 | 2.47 | 1322.68 | 0.02 |
| L CA2 curvature | Stable AD | 0.0058 | 0.00 | 2.22 | 1335.53 | 0.04 |
| L CA2 curvature | CN to MCI/AD | -0.0105 | 0.00 | -2.67 | 1290.94 | 0.01 |
| L CA2 curvature | MCI to AD | 0.0010 | 0.00 | 0.36 | 1307.85 | 0.75 |
| L CA2 curvature | Followup Years | 0.0003 | 0.00 | 2.64 | 710.30 | 0.01 |
| L CA2 curvature | Age | 0.0040 | 9.81E-05 | 41.30 | 1834.45 | < 0.01 |
| L CA2 curvature | Education | -0.0029 | 0.00 | -9.71 | 1310.13 | < 0.01 |
| L CA2 curvature | Scanner Site | 0.0000 | 6.94E-06 | -4.59 | 1206.49 | < 0.01 |
| L CA2 curvature | SexM | -0.0369 | 0.00 | -21.91 | 1315.62 | < 0.01 |
| L CA2 curvature | Stable MCI:Followup Years | 0.0003 | 0.00 | 1.75 | 662.31 | 0.10 |
| L CA2 curvature | Stable AD:Followup Years | 0.0010 | 0.00 | 2.68 | 892.82 | 0.01 |

| | | | | | | |
|---|---|---|---|---|---|---|
| L CA2 curvature | CN to MCI/AD:Followup Years | 0.0010 | 0.00 | 4.17 | 506.39 | < 0.01 |
| L CA2 curvature | MCI to AD:Followup Years | 0.0012 | 0.00 | 5.89 | 775.37 | < 0.01 |
| L CA2 gyrification | (Intercept) | -0.7130 | 0.01 | -78.71 | 1759.89 | < 0.01 |
| L CA2 gyrification | Stable MCI | 0.0056 | 0.00 | 2.52 | 1321.71 | 0.02 |
| L CA2 gyrification | Stable AD | 0.0063 | 0.00 | 2.44 | 1334.93 | 0.02 |
| L CA2 gyrification | CN to MCI/AD | -0.0108 | 0.00 | -2.76 | 1289.28 | 0.01 |
| L CA2 gyrification | MCI to AD | 0.0011 | 0.00 | 0.41 | 1306.61 | 0.72 |
| L CA2 gyrification | Followup Years | 0.0003 | 0.00 | 2.47 | 701.20 | 0.02 |
| L CA2 gyrification | Age | 0.0041 | 9.82E-05 | 41.74 | 1836.02 | < 0.01 |
| L CA2 gyrification | Education | -0.0029 | 0.00 | -9.69 | 1309.85 | < 0.01 |
| L CA2 gyrification | Scanner Site | 0.0000 | 6.91E-06 | -4.66 | 1210.10 | < 0.01 |
| L CA2 gyrification | SexM | -0.0365 | 0.00 | -21.80 | 1315.42 | < 0.01 |
| L CA2 gyrification | Stable MCI:Followup Years | 0.0003 | 0.00 | 1.83 | 654.19 | 0.09 |
| L CA2 gyrification | Stable AD:Followup Years | 0.0013 | 0.00 | 3.41 | 895.78 | < 0.01 |
| L CA2 gyrification | CN to MCI/AD:Followup Years | 0.0010 | 0.00 | 4.26 | 498.84 | < 0.01 |
| L CA2 gyrification | MCI to AD:Followup Years | 0.0014 | 0.00 | 6.99 | 763.23 | < 0.01 |
| L CA2 thickness | (Intercept) | -0.7114 | 0.01 | -78.55 | 1762.58 | < 0.01 |
| L CA2 thickness | Stable MCI | 0.0057 | 0.00 | 2.57 | 1322.42 | 0.02 |
| L CA2 thickness | Stable AD | 0.0063 | 0.00 | 2.43 | 1335.37 | 0.02 |
| L CA2 thickness | CN to MCI/AD | -0.0105 | 0.00 | -2.67 | 1290.38 | 0.01 |
| L CA2 thickness | MCI to AD | 0.0014 | 0.00 | 0.52 | 1307.47 | 0.66 |
| L CA2 thickness | Followup Years | 0.0003 | 0.00 | 2.52 | 705.66 | 0.02 |
| L CA2 thickness | Age | 0.0041 | 9.80E-05 | 41.60 | 1833.40 | < 0.01 |
| L CA2 thickness | Education | -0.0029 | 0.00 | -9.70 | 1309.89 | < 0.01 |
| L CA2 thickness | Scanner Site | 0.0000 | 6.93E-06 | -4.61 | 1207.12 | < 0.01 |

| | | | | | | |
|---|---|---|---|---|---|---|
| L CA2 thickness | SexM | -0.0366 | 0.00 | -21.82 | 1315.40 | < 0.01 |
| L CA2 thickness | Stable MCI:Followup Years | 0.0003 | 0.00 | 1.79 | 658.11 | 0.10 |
| L CA2 thickness | Stable AD:Followup Years | 0.0010 | 0.00 | 2.88 | 888.02 | 0.01 |
| L CA2 thickness | CN to MCI/AD:Followup Years | 0.0010 | 0.00 | 4.25 | 502.62 | < 0.01 |
| L CA2 thickness | MCI to AD:Followup Years | 0.0013 | 0.00 | 6.35 | 770.45 | < 0.01 |
| L CA3 volume | (Intercept) | 1.0847 | 0.13 | 8.30 | 1454.85 | < 0.01 |
| L CA3 volume | Stable MCI | -0.1660 | 0.03 | -5.82 | 1318.01 | < 0.01 |
| L CA3 volume | Stable AD | -0.2890 | 0.03 | -8.62 | 1385.71 | < 0.01 |
| L CA3 volume | CN to MCI/AD | -0.1371 | 0.05 | -2.79 | 1184.61 | 0.01 |
| L CA3 volume | MCI to AD | -0.3014 | 0.03 | -9.09 | 1261.25 | < 0.01 |
| L CA3 volume | Followup Years | -0.0007 | 0.00 | -0.19 | 341.16 | 0.87 |
| L CA3 volume | Age | -0.0001 | 0.00 | -0.04 | 1489.57 | 0.98 |
| L CA3 volume | Education | 0.0019 | 0.00 | 0.49 | 1284.61 | 0.67 |
| L CA3 volume | Scanner Site | 8.31E-05 | 9.27E-05 | 0.90 | 1351.40 | 0.42 |
| L CA3 volume | SexM | 0.1422 | 0.02 | 6.06 | 1459.48 | < 0.01 |
| L CA3 volume | ETIV | -0.0183 | 0.01 | -1.93 | 3167.41 | 0.07 |
| L CA3 volume | Stable MCI:Followup Years | -0.0001 | 0.01 | -0.02 | 348.67 | 0.99 |
| L CA3 volume | Stable AD:Followup Years | 0.0276 | 0.01 | 2.61 | 577.09 | 0.02 |
| L CA3 volume | CN to MCI/AD:Followup Years | -0.0060 | 0.01 | -0.85 | 256.39 | 0.44 |
| L CA3 volume | MCI to AD:Followup Years | 0.0272 | 0.01 | 4.66 | 408.16 | < 0.01 |
| L CA3 curvature | (Intercept) | -0.7159 | 0.01 | -79.00 | 1764.24 | < 0.01 |
| L CA3 curvature | Stable MCI | 0.0055 | 0.00 | 2.45 | 1322.73 | 0.02 |
| L CA3 curvature | Stable AD | 0.0057 | 0.00 | 2.17 | 1335.58 | 0.04 |
| L CA3 curvature | CN to MCI/AD | -0.0105 | 0.00 | -2.67 | 1290.99 | 0.01 |
| L CA3 curvature | MCI to AD | 0.0009 | 0.00 | 0.34 | 1307.91 | 0.77 |

| | | | | | | |
|---|---|---|---|---|---|---|
| L CA3 curvature | Followup Years | 0.0003 | 0.00 | 2.64 | 710.76 | 0.01 |
| L CA3 curvature | Age | 0.0040 | 9.80E-05 | 41.25 | 1833.93 | < 0.01 |
| L CA3 curvature | Education | -0.0029 | 0.00 | -9.72 | 1310.13 | < 0.01 |
| L CA3 curvature | Scanner Site | 0.0000 | 6.94E-06 | -4.60 | 1205.77 | < 0.01 |
| L CA3 curvature | SexM | -0.0368 | 0.00 | -21.91 | 1315.59 | < 0.01 |
| L CA3 curvature | Stable MCI:Followup Years | 0.0003 | 0.00 | 1.73 | 662.81 | 0.11 |
| L CA3 curvature | Stable AD:Followup Years | 0.0010 | 0.00 | 2.66 | 892.75 | 0.01 |
| L CA3 curvature | CN to MCI/AD:Followup Years | 0.0010 | 0.00 | 4.16 | 506.74 | < 0.01 |
| L CA3 curvature | MCI to AD:Followup Years | 0.0012 | 0.00 | 5.80 | 775.94 | < 0.01 |
| L CA3 gyrification | (Intercept) | -0.7033 | 0.01 | -78.02 | 1745.67 | 0.00 |
| L CA3 gyrification | Stable MCI | 0.0047 | 0.00 | 2.13 | 1322.80 | 0.05 |
| L CA3 gyrification | Stable AD | 0.0048 | 0.00 | 1.85 | 1336.40 | 0.09 |
| L CA3 gyrification | CN to MCI/AD | -0.0112 | 0.00 | -2.87 | 1289.10 | 0.01 |
| L CA3 gyrification | MCI to AD | -0.0004 | 0.00 | -0.16 | 1307.16 | 0.89 |
| L CA3 gyrification | Followup Years | 0.0003 | 0.00 | 2.42 | 695.01 | 0.02 |
| L CA3 gyrification | Age | 0.0041 | 9.78E-05 | 41.42 | 1814.23 | < 0.01 |
| L CA3 gyrification | Education | -0.0029 | 0.00 | -9.82 | 1310.02 | < 0.01 |
| L CA3 gyrification | Scanner Site | 0.0000 | 6.86E-06 | -4.60 | 1203.77 | < 0.01 |
| L CA3 gyrification | SexM | -0.0360 | 0.00 | -21.66 | 1315.23 | < 0.01 |
| L CA3 gyrification | Stable MCI:Followup Years | 0.0003 | 0.00 | 1.71 | 650.69 | 0.11 |
| L CA3 gyrification | Stable AD:Followup Years | 0.0012 | 0.00 | 3.21 | 873.08 | < 0.01 |
| L CA3 gyrification | CN to MCI/AD:Followup Years | 0.0010 | 0.00 | 4.08 | 496.62 | < 0.01 |
| L CA3 gyrification | MCI to AD:Followup Years | 0.0014 | 0.00 | 6.71 | 762.13 | < 0.01 |
| L CA3 thickness | (Intercept) | -0.7076 | 0.01 | -78.21 | 1760.64 | < 0.01 |
| L CA3 thickness | Stable MCI | 0.0055 | 0.00 | 2.45 | 1322.88 | 0.02 |

| | | | | | | |
|---|---|---|---|---|---|---|
| L CA3 thickness | Stable AD | 0.0057 | 0.00 | 2.17 | 1335.82 | 0.04 |
| L CA3 thickness | CN to MCI/AD | -0.0105 | 0.00 | -2.69 | 1290.92 | 0.01 |
| L CA3 thickness | MCI to AD | 0.0008 | 0.00 | 0.32 | 1307.96 | 0.78 |
| L CA3 thickness | Followup Years | 0.0003 | 0.00 | 2.62 | 709.60 | 0.01 |
| L CA3 thickness | Age | 0.0041 | 9.79E-05 | 41.38 | 1829.32 | < 0.01 |
| L CA3 thickness | Education | -0.0029 | 0.00 | -9.71 | 1309.72 | < 0.01 |
| L CA3 thickness | Scanner Site | 0.0000 | 6.92E-06 | -4.59 | 1204.19 | < 0.01 |
| L CA3 thickness | SexM | -0.0366 | 0.00 | -21.80 | 1315.10 | < 0.01 |
| L CA3 thickness | Stable MCI:Followup Years | 0.0003 | 0.00 | 1.74 | 662.18 | 0.10 |
| L CA3 thickness | Stable AD:Followup Years | 0.0010 | 0.00 | 2.67 | 890.26 | 0.01 |
| L CA3 thickness | CN to MCI/AD:Followup Years | 0.0010 | 0.00 | 4.16 | 506.34 | < 0.01 |
| L CA3 thickness | MCI to AD:Followup Years | 0.0012 | 0.00 | 5.86 | 775.42 | < 0.01 |
| L CA4 volume | (Intercept) | 0.4195 | 0.08 | 5.39 | 1448.75 | < 0.01 |
| L CA4 volume | Stable MCI | -0.0909 | 0.02 | -5.24 | 1350.59 | < 0.01 |
| L CA4 volume | Stable AD | -0.2039 | 0.02 | -10.00 | 1414.16 | < 0.01 |
| L CA4 volume | CN to MCI/AD | -0.0398 | 0.03 | -1.33 | 1211.98 | 0.22 |
| L CA4 volume | MCI to AD | -0.1889 | 0.02 | -9.36 | 1291.93 | < 0.01 |
| L CA4 volume | Followup Years | -0.0006 | 0.00 | -0.28 | 384.27 | 0.81 |
| L CA4 volume | Age | -0.0008 | 0.00 | -0.90 | 1472.57 | 0.42 |
| L CA4 volume | Education | -0.0038 | 0.00 | -1.63 | 1280.06 | 0.13 |
| L CA4 volume | Scanner Site | 4.02E-05 | 5.50E-05 | 0.73 | 1296.31 | 0.52 |
| L CA4 volume | SexM | 0.0208 | 0.01 | 1.49 | 1455.02 | 0.17 |
| L CA4 volume | ETIV | 0.0021 | 0.01 | 0.37 | 3116.41 | 0.75 |
| L CA4 volume | Stable MCI:Followup Years | -0.0039 | 0.00 | -1.27 | 393.92 | 0.24 |
| L CA4 volume | Stable AD:Followup Years | -0.0061 | 0.01 | -0.98 | 547.77 | 0.38 |

| | | | | | | |
|---|---|---|---|---|---|---|
| L CA4 volume | CN to MCI/AD:Followup Years | -0.0011 | 0.00 | -0.26 | 299.80 | 0.82 |
| L CA4 volume | MCI to AD:Followup Years | -0.0102 | 0.00 | -2.95 | 475.70 | 0.01 |
| L CA4 curvature | (Intercept) | -0.7009 | 0.01 | -75.49 | 1688.23 | < 0.01 |
| L CA4 curvature | Stable MCI | 0.0044 | 0.00 | 1.93 | 1327.91 | 0.07 |
| L CA4 curvature | Stable AD | 0.0033 | 0.00 | 1.26 | 1343.49 | 0.25 |
| L CA4 curvature | CN to MCI/AD | -0.0104 | 0.00 | -2.63 | 1286.50 | 0.01 |
| L CA4 curvature | MCI to AD | -0.0009 | 0.00 | -0.35 | 1308.92 | 0.76 |
| L CA4 curvature | Followup Years | 0.0003 | 0.00 | 2.43 | 638.76 | 0.02 |
| L CA4 curvature | Age | 0.0039 | 0.00 | 38.53 | 1734.05 | < 0.01 |
| L CA4 curvature | Education | -0.0030 | 0.00 | -10.12 | 1311.22 | < 0.01 |
| L CA4 curvature | Scanner Site | 0.0000 | 6.95E-06 | -4.47 | 1193.45 | < 0.01 |
| L CA4 curvature | SexM | -0.0372 | 0.00 | -22.05 | 1315.60 | < 0.01 |
| L CA4 curvature | Stable MCI:Followup Years | 0.0003 | 0.00 | 1.70 | 605.80 | 0.11 |
| L CA4 curvature | Stable AD:Followup Years | 0.0007 | 0.00 | 1.70 | 758.88 | 0.11 |
| L CA4 curvature | CN to MCI/AD:Followup Years | 0.0010 | 0.00 | 3.57 | 461.34 | < 0.01 |
| L CA4 curvature | MCI to AD:Followup Years | 0.0010 | 0.00 | 4.60 | 722.62 | < 0.01 |
| L CA4 gyrification | (Intercept) | -0.6746 | 0.01 | -74.76 | 1709.51 | < 0.01 |
| L CA4 gyrification | Stable MCI | 0.0031 | 0.00 | 1.39 | 1325.40 | 0.20 |
| L CA4 gyrification | Stable AD | 0.0017 | 0.00 | 0.67 | 1340.73 | 0.56 |
| L CA4 gyrification | CN to MCI/AD | -0.0112 | 0.00 | -2.93 | 1286.43 | 0.01 |
| L CA4 gyrification | MCI to AD | -0.0029 | 0.00 | -1.13 | 1307.51 | 0.31 |
| L CA4 gyrification | Followup Years | 0.0003 | 0.00 | 2.04 | 664.59 | 0.06 |
| L CA4 gyrification | Age | 0.0039 | 9.85E-05 | 39.81 | 1776.61 | < 0.01 |
| L CA4 gyrification | Education | -0.0029 | 0.00 | -10.08 | 1310.95 | < 0.01 |
| L CA4 gyrification | Scanner Site | 0.0000 | 6.78E-06 | -4.42 | 1210.55 | < 0.01 |

| | | | | | | |
|---|---|---|---|---|---|---|
| L CA4 gyrification | SexM | -0.0348 | 0.00 | -21.23 | 1316.11 | < 0.01 |
| L CA4 gyrification | Stable MCI:Followup Years | 0.0002 | 0.00 | 1.23 | 627.33 | 0.26 |
| L CA4 gyrification | Stable AD:Followup Years | 0.0011 | 0.00 | 2.71 | 832.52 | 0.01 |
| L CA4 gyrification | CN to MCI/AD:Followup Years | 0.0011 | 0.00 | 4.01 | 477.97 | < 0.01 |
| L CA4 gyrification | MCI to AD:Followup Years | 0.0011 | 0.00 | 5.12 | 737.98 | < 0.01 |
| L CA4 thickness | (Intercept) | -0.7034 | 0.01 | -77.66 | 1755.23 | 0.00 |
| L CA4 thickness | Stable MCI | 0.0051 | 0.00 | 2.27 | 1322.98 | 0.03 |
| L CA4 thickness | Stable AD | 0.0050 | 0.00 | 1.90 | 1336.08 | 0.08 |
| L CA4 thickness | CN to MCI/AD | -0.0106 | 0.00 | -2.72 | 1290.84 | 0.01 |
| L CA4 thickness | MCI to AD | 0.0003 | 0.00 | 0.10 | 1308.01 | 0.93 |
| L CA4 thickness | Followup Years | 0.0003 | 0.00 | 2.51 | 709.76 | 0.02 |
| L CA4 thickness | Age | 0.0040 | 9.80E-05 | 41.12 | 1822.63 | < 0.01 |
| L CA4 thickness | Education | -0.0029 | 0.00 | -9.72 | 1309.48 | < 0.01 |
| L CA4 thickness | Scanner Site | 0.0000 | 6.92E-06 | -4.60 | 1200.81 | < 0.01 |
| L CA4 thickness | SexM | -0.0365 | 0.00 | -21.75 | 1314.67 | < 0.01 |
| L CA4 thickness | Stable MCI:Followup Years | 0.0003 | 0.00 | 1.78 | 663.23 | 0.10 |
| L CA4 thickness | Stable AD:Followup Years | 0.0010 | 0.00 | 2.79 | 892.06 | 0.01 |
| L CA4 thickness | CN to MCI/AD:Followup Years | 0.0010 | 0.00 | 3.97 | 507.41 | < 0.01 |
| L CA4 thickness | MCI to AD:Followup Years | 0.0012 | 0.00 | 5.82 | 775.90 | < 0.01 |
| L DG volume | (Intercept) | 0.5750 | 0.05 | 12.50 | 1489.54 | < 0.01 |
| L DG volume | Stable MCI | -0.1119 | 0.01 | -11.23 | 1311.41 | < 0.01 |
| L DG volume | Stable AD | -0.2717 | 0.01 | -23.26 | 1350.64 | < 0.01 |
| L DG volume | CN to MCI/AD | -0.0474 | 0.02 | -2.73 | 1246.48 | 0.01 |
| L DG volume | MCI to AD | -0.2144 | 0.01 | -18.41 | 1284.49 | < 0.01 |
| L DG volume | Followup Years | -0.0006 | 0.00 | -0.48 | 321.14 | 0.68 |

| | | | | | | |
|---|---|---|---|---|---|---|
| L DG volume | Age | -0.0053 | 0.00 | -10.24 | 1557.34 | < 0.01 |
| L DG volume | Education | -0.0026 | 0.00 | -1.89 | 1292.14 | 0.08 |
| L DG volume | Scanner Site | 3.44E-05 | 3.27E-05 | 1.05 | 1341.66 | 0.34 |
| L DG volume | SexM | 0.0158 | 0.01 | 1.93 | 1465.06 | 0.07 |
| L DG volume | ETIV | -0.0314 | 0.00 | -11.23 | 3866.03 | < 0.01 |
| L DG volume | Stable MCI:Followup Years | -0.0052 | 0.00 | -2.75 | 313.97 | 0.01 |
| L DG volume | Stable AD:Followup Years | -0.0117 | 0.00 | -3.53 | 1031.69 | < 0.01 |
| L DG volume | CN to MCI/AD:Followup Years | -0.0108 | 0.00 | -4.09 | 230.56 | < 0.01 |
| L DG volume | MCI to AD:Followup Years | -0.0156 | 0.00 | -7.63 | 329.78 | < 0.01 |
| L DG curvature | (Intercept) | -0.7187 | 0.01 | -76.32 | 1697.78 | < 0.01 |
| L DG curvature | Stable MCI | 0.0061 | 0.00 | 2.72 | 1324.07 | 0.01 |
| L DG curvature | Stable AD | 0.0067 | 0.00 | 2.56 | 1339.92 | 0.02 |
| L DG curvature | CN to MCI/AD | -0.0103 | 0.00 | -2.62 | 1283.06 | 0.01 |
| L DG curvature | MCI to AD | 0.0016 | 0.00 | 0.60 | 1305.23 | 0.60 |
| L DG curvature | Followup Years | 0.0004 | 0.00 | 2.51 | 541.18 | 0.02 |
| L DG curvature | Age | 0.0041 | 0.00 | 39.45 | 1758.61 | < 0.01 |
| L DG curvature | Education | -0.0029 | 0.00 | -9.50 | 1311.27 | < 0.01 |
| L DG curvature | Scanner Site | 0.0000 | 7.08E-06 | -4.34 | 1255.69 | < 0.01 |
| L DG curvature | SexM | -0.0370 | 0.00 | -21.73 | 1318.26 | < 0.01 |
| L DG curvature | Stable MCI:Followup Years | 0.0003 | 0.00 | 1.51 | 509.73 | 0.16 |
| L DG curvature | Stable AD:Followup Years | 0.0009 | 0.00 | 2.26 | 715.54 | 0.03 |
| L DG curvature | CN to MCI/AD:Followup Years | 0.0011 | 0.00 | 3.90 | 385.19 | < 0.01 |
| L DG curvature | MCI to AD:Followup Years | 0.0013 | 0.00 | 5.73 | 603.62 | < 0.01 |
| L DG gyrification | (Intercept) | -0.6671 | 0.01 | -73.92 | 1716.31 | < 0.01 |
| L DG gyrification | Stable MCI | 0.0021 | 0.00 | 0.96 | 1325.34 | 0.39 |

| | | | | | | |
|---|---|---|---|---|---|---|
| L DG gyrification | Stable AD | -0.0023 | 0.00 | -0.90 | 1340.14 | 0.42 |
| L DG gyrification | CN to MCI/AD | -0.0119 | 0.00 | -3.08 | 1289.08 | < 0.01 |
| L DG gyrification | MCI to AD | -0.0056 | 0.00 | -2.16 | 1308.66 | 0.04 |
| L DG gyrification | Followup Years | 0.0004 | 0.00 | 2.76 | 675.57 | 0.01 |
| L DG gyrification | Age | 0.0038 | 9.83E-05 | 38.53 | 1781.79 | < 0.01 |
| L DG gyrification | Education | -0.0029 | 0.00 | -10.08 | 1310.93 | < 0.01 |
| L DG gyrification | Scanner Site | 0.0000 | 6.80E-06 | -4.33 | 1203.79 | < 0.01 |
| L DG gyrification | SexM | -0.0352 | 0.00 | -21.36 | 1315.80 | < 0.01 |
| L DG gyrification | Stable MCI:Followup Years | 0.0001 | 0.00 | 0.72 | 637.08 | 0.53 |
| L DG gyrification | Stable AD:Followup Years | 0.0008 | 0.00 | 1.96 | 865.51 | 0.07 |
| L DG gyrification | CN to MCI/AD:Followup Years | 0.0007 | 0.00 | 2.77 | 487.17 | 0.01 |
| L DG gyrification | MCI to AD:Followup Years | 0.0007 | 0.00 | 3.33 | 744.41 | < 0.01 |
| L Sub volume | (Intercept) | 2.2677 | 0.11 | 21.24 | 1816.65 | < 0.01 |
| L Sub volume | Stable MCI | -0.1662 | 0.03 | -6.29 | 1302.94 | < 0.01 |
| L Sub volume | Stable AD | -0.3986 | 0.03 | -12.94 | 1309.26 | < 0.01 |
| L Sub volume | CN to MCI/AD | -0.0420 | 0.05 | -0.90 | 1285.56 | 0.42 |
| L Sub volume | MCI to AD | -0.3131 | 0.03 | -10.10 | 1293.78 | < 0.01 |
| L Sub volume | Followup Years | 0.0044 | 0.00 | 3.31 | 336.19 | < 0.01 |
| L Sub volume | Age | -0.0089 | 0.00 | -7.84 | 1724.91 | < 0.01 |
| L Sub volume | Education | 0.0046 | 0.00 | 1.25 | 1298.46 | 0.25 |
| L Sub volume | Scanner Site | 3.65E-05 | 8.68E-05 | 0.42 | 1306.30 | 0.72 |
| L Sub volume | SexM | 0.1356 | 0.02 | 6.43 | 1395.20 | < 0.01 |
| L Sub volume | ETIV | 0.0128 | 0.00 | 2.90 | 4087.08 | 0.01 |
| L Sub volume | Stable MCI:Followup Years | -0.0012 | 0.00 | -0.71 | 298.19 | 0.53 |
| L Sub volume | Stable AD:Followup Years | -0.0103 | 0.00 | -2.90 | 531.76 | 0.01 |

| | | | | | | |
|---|---|---|---|---|---|---|
| L Sub volume | CN to MCI/AD:Followup Years | -0.0050 | 0.00 | -2.12 | 217.69 | 0.05 |
| L Sub volume | MCI to AD:Followup Years | -0.0017 | 0.00 | -0.88 | 351.25 | 0.43 |
| L Sub curvature | (Intercept) | -0.7019 | 0.01 | -66.89 | 1454.11 | < 0.01 |
| L Sub curvature | Stable MCI | 0.0053 | 0.00 | 2.30 | 1362.87 | 0.03 |
| L Sub curvature | Stable AD | 0.0062 | 0.00 | 2.28 | 1465.03 | 0.03 |
| L Sub curvature | CN to MCI/AD | -0.0105 | 0.00 | -2.69 | 1174.62 | 0.01 |
| L Sub curvature | MCI to AD | 0.0003 | 0.00 | 0.12 | 1286.86 | 0.91 |
| L Sub curvature | Followup Years | 0.0005 | 0.00 | 1.36 | 387.91 | 0.21 |
| L Sub curvature | Age | 0.0039 | 0.00 | 32.43 | 1477.49 | < 0.01 |
| L Sub curvature | Education | -0.0029 | 0.00 | -9.47 | 1314.92 | < 0.01 |
| L Sub curvature | Scanner Site | 0.0000 | 7.40E-06 | -3.80 | 1430.24 | < 0.01 |
| L Sub curvature | SexM | -0.0374 | 0.00 | -21.59 | 1327.82 | < 0.01 |
| L Sub curvature | Stable MCI:Followup Years | 0.0003 | 0.00 | 0.47 | 407.16 | 0.68 |
| L Sub curvature | Stable AD:Followup Years | 0.0007 | 0.00 | 0.65 | 865.14 | 0.57 |
| L Sub curvature | CN to MCI/AD:Followup Years | 0.0010 | 0.00 | 1.31 | 291.54 | 0.23 |
| L Sub curvature | MCI to AD:Followup Years | 0.0019 | 0.00 | 3.03 | 444.86 | 0.01 |
| L Sub gyrification | (Intercept) | -0.7003 | 0.01 | -77.74 | 1758.93 | 0.00 |
| L Sub gyrification | Stable MCI | 0.0048 | 0.00 | 2.14 | 1323.62 | 0.04 |
| L Sub gyrification | Stable AD | 0.0041 | 0.00 | 1.59 | 1336.64 | 0.14 |
| L Sub gyrification | CN to MCI/AD | -0.0107 | 0.00 | -2.76 | 1291.26 | 0.01 |
| L Sub gyrification | MCI to AD | -0.0004 | 0.00 | -0.15 | 1308.52 | 0.90 |
| L Sub gyrification | Followup Years | 0.0003 | 0.00 | 2.62 | 711.61 | 0.01 |
| L Sub gyrification | Age | 0.0040 | 9.75E-05 | 40.59 | 1826.85 | < 0.01 |
| L Sub gyrification | Education | -0.0029 | 0.00 | -9.83 | 1310.75 | < 0.01 |
| L Sub gyrification | Scanner Site | 0.0000 | 6.89E-06 | -4.63 | 1204.95 | < 0.01 |

| | | | | | | |
|---|---|---|---|---|---|---|
| L Sub gyrification | SexM | -0.0364 | 0.00 | -21.82 | 1316.10 | < 0.01 |
| L Sub gyrification | Stable MCI:Followup Years | 0.0003 | 0.00 | 1.69 | 664.46 | 0.11 |
| L Sub gyrification | Stable AD:Followup Years | 0.0008 | 0.00 | 2.34 | 887.38 | 0.03 |
| L Sub gyrification | CN to MCI/AD:Followup Years | 0.0010 | 0.00 | 3.95 | 508.29 | < 0.01 |
| L Sub gyrification | MCI to AD:Followup Years | 0.0010 | 0.00 | 5.14 | 779.20 | < 0.01 |
| L Sub thickness | (Intercept) | -0.7010 | 0.01 | -64.58 | 1264.62 | < 0.01 |
| L Sub thickness | Stable MCI | 0.0053 | 0.00 | 2.04 | 964.85 | 0.06 |
| L Sub thickness | Stable AD | 0.0061 | 0.00 | 2.00 | 974.69 | 0.06 |
| L Sub thickness | CN to MCI/AD | -0.0085 | 0.00 | -1.78 | 940.61 | 0.10 |
| L Sub thickness | MCI to AD | 0.0015 | 0.00 | 0.50 | 953.64 | 0.67 |
| L Sub thickness | Followup Years | 0.0004 | 0.00 | 2.88 | 557.72 | 0.01 |
| L Sub thickness | Age | 0.0040 | 0.00 | 34.06 | 1330.41 | < 0.01 |
| L Sub thickness | Education | -0.0031 | 0.00 | -8.76 | 957.43 | < 0.01 |
| L Sub thickness | Scanner Site | 7.88E-06 | 3.10E-05 | 0.25 | 952.85 | 0.82 |
| L Sub thickness | SexM | -0.0355 | 0.00 | -18.01 | 965.88 | < 0.01 |
| L Sub thickness | Stable MCI:Followup Years | 0.0002 | 0.00 | 0.88 | 525.49 | 0.43 |
| L Sub thickness | Stable AD:Followup Years | 0.0013 | 0.00 | 2.82 | 1303.36 | 0.01 |
| L Sub thickness | CN to MCI/AD:Followup Years | 0.0008 | 0.00 | 2.46 | 358.21 | 0.02 |
| L Sub thickness | MCI to AD:Followup Years | 0.0012 | 0.00 | 5.15 | 557.50 | < 0.01 |
| L SRLM volume | (Intercept) | 3.9508 | 0.15 | 27.14 | 1520.07 | < 0.01 |
| L SRLM volume | Stable MCI | -0.3446 | 0.03 | -10.88 | 1302.24 | < 0.01 |
| L SRLM volume | Stable AD | -0.8281 | 0.04 | -22.32 | 1335.29 | < 0.01 |
| L SRLM volume | CN to MCI/AD | -0.1694 | 0.06 | -3.07 | 1244.92 | < 0.01 |
| L SRLM volume | MCI to AD | -0.6667 | 0.04 | -18.00 | 1278.02 | < 0.01 |
| L SRLM volume | Followup Years | -0.0032 | 0.00 | -0.88 | 373.81 | 0.43 |

| | | | | | | |
|---|---|---|---|---|---|---|
| L SRLM volume | Age | -0.0234 | 0.00 | -14.19 | 1603.67 | < 0.01 |
| L SRLM volume | Education | -0.0002 | 0.00 | -0.04 | 1284.17 | 0.97 |
| L SRLM volume | Scanner Site | 0.0001 | 0.00 | 1.01 | 1329.29 | 0.36 |
| L SRLM volume | SexM | 0.1544 | 0.03 | 5.95 | 1452.43 | < 0.01 |
| L SRLM volume | ETIV | 0.0251 | 0.01 | 2.93 | 4082.57 | 0.01 |
| L SRLM volume | Stable MCI:Followup Years | -0.0139 | 0.01 | -2.69 | 359.34 | 0.01 |
| L SRLM volume | Stable AD:Followup Years | -0.0268 | 0.01 | -2.83 | 1023.37 | 0.01 |
| L SRLM volume | CN to MCI/AD:Followup Years | -0.0229 | 0.01 | -3.17 | 263.98 | < 0.01 |
| L SRLM volume | MCI to AD:Followup Years | -0.0438 | 0.01 | -7.71 | 384.37 | < 0.01 |
| L Cyst volume | (Intercept) | -0.4945 | 0.02 | -19.94 | 1775.78 | < 0.01 |
| L Cyst volume | Stable MCI | -0.0194 | 0.01 | -3.21 | 1319.44 | < 0.01 |
| L Cyst volume | Stable AD | -0.0368 | 0.01 | -5.22 | 1330.67 | < 0.01 |
| L Cyst volume | CN to MCI/AD | -0.0083 | 0.01 | -0.78 | 1295.52 | 0.48 |
| L Cyst volume | MCI to AD | -0.0199 | 0.01 | -2.80 | 1308.21 | 0.01 |
| L Cyst volume | Followup Years | 0.0005 | 0.00 | 1.47 | 406.53 | 0.17 |
| L Cyst volume | Age | 0.0022 | 0.00 | 8.29 | 1817.71 | < 0.01 |
| L Cyst volume | Education | -0.0029 | 0.00 | -3.54 | 1339.01 | < 0.01 |
| L Cyst volume | Scanner Site | 0.0000 | 1.90E-05 | -1.45 | 1304.63 | 0.18 |
| L Cyst volume | SexM | -0.0156 | 0.00 | -3.33 | 1459.97 | < 0.01 |
| L Cyst volume | ETIV | -0.0304 | 0.00 | -26.18 | 4310.37 | < 0.01 |
| L Cyst volume | Stable MCI:Followup Years | -0.0004 | 0.00 | -0.86 | 376.45 | 0.44 |
| L Cyst volume | Stable AD:Followup Years | -0.0032 | 0.00 | -3.25 | 686.85 | < 0.01 |
| L Cyst volume | CN to MCI/AD:Followup Years | -0.0012 | 0.00 | -1.70 | 283.56 | 0.11 |
| L Cyst volume | MCI to AD:Followup Years | -0.0033 | 0.00 | -5.90 | 432.00 | < 0.01 |
| R CA1 volume | (Intercept) | 6.2576 | 0.21 | 30.15 | 1550.78 | < 0.01 |

| | | | | | | |
|---|---|---|---|---|---|---|
| R CA1 volume | Stable MCI | -0.4195 | 0.05 | -9.15 | 1307.11 | < 0.01 |
| R CA1 volume | Stable AD | -0.9012 | 0.05 | -16.80 | 1337.54 | < 0.01 |
| R CA1 volume | CN to MCI/AD | -0.1208 | 0.08 | -1.51 | 1249.40 | 0.16 |
| R CA1 volume | MCI to AD | -0.7389 | 0.05 | -13.78 | 1281.97 | < 0.01 |
| R CA1 volume | Followup Years | -0.0108 | 0.00 | -2.32 | 449.57 | 0.03 |
| R CA1 volume | Age | -0.0280 | 0.00 | -11.96 | 1640.80 | < 0.01 |
| R CA1 volume | Education | -0.0031 | 0.01 | -0.49 | 1289.48 | 0.68 |
| R CA1 volume | Scanner Site | 0.0002 | 0.00 | 1.09 | 1326.74 | 0.32 |
| R CA1 volume | SexM | 0.1992 | 0.04 | 5.35 | 1459.21 | < 0.01 |
| R CA1 volume | ETIV | 0.1786 | 0.01 | 14.51 | 4285.33 | < 0.01 |
| R CA1 volume | Stable MCI:Followup Years | -0.0044 | 0.01 | -0.66 | 432.49 | 0.56 |
| R CA1 volume | Stable AD:Followup Years | -0.0583 | 0.01 | -4.57 | 987.41 | < 0.01 |
| R CA1 volume | CN to MCI/AD:Followup Years | -0.0298 | 0.01 | -3.24 | 319.77 | < 0.01 |
| R CA1 volume | MCI to AD:Followup Years | -0.0360 | 0.01 | -4.89 | 475.08 | < 0.01 |
| R CA1 curvature | (Intercept) | -0.7156 | 0.01 | -78.84 | 1764.38 | < 0.01 |
| R CA1 curvature | Stable MCI | 0.0057 | 0.00 | 2.52 | 1322.29 | 0.02 |
| R CA1 curvature | Stable AD | 0.0060 | 0.00 | 2.31 | 1335.17 | 0.03 |
| R CA1 curvature | CN to MCI/AD | -0.0105 | 0.00 | -2.66 | 1290.44 | 0.01 |
| R CA1 curvature | MCI to AD | 0.0012 | 0.00 | 0.44 | 1307.30 | 0.70 |
| R CA1 curvature | Followup Years | 0.0003 | 0.00 | 2.57 | 707.70 | 0.02 |
| R CA1 curvature | Age | 0.0041 | 9.82E-05 | 41.42 | 1833.78 | < 0.01 |
| R CA1 curvature | Education | -0.0029 | 0.00 | -9.70 | 1309.87 | < 0.01 |
| R CA1 curvature | Scanner Site | 0.0000 | 6.96E-06 | -4.60 | 1206.91 | < 0.01 |
| R CA1 curvature | SexM | -0.0369 | 0.00 | -21.92 | 1315.29 | < 0.01 |
| R CA1 curvature | Stable MCI:Followup Years | 0.0003 | 0.00 | 1.79 | 660.79 | 0.09 |

| | | | | | | |
|---|---|---|---|---|---|---|
| R CA1 curvature | Stable AD:Followup Years | 0.0010 | 0.00 | 2.70 | 892.61 | 0.01 |
| R CA1 curvature | CN to MCI/AD:Followup Years | 0.0010 | 0.00 | 4.20 | 505.58 | < 0.01 |
| R CA1 curvature | MCI to AD:Followup Years | 0.0012 | 0.00 | 5.95 | 773.78 | < 0.01 |
| R CA1 gyrification | (Intercept) | -0.6943 | 0.01 | -77.61 | 1755.24 | < 0.01 |
| R CA1 gyrification | Stable MCI | 0.0045 | 0.00 | 2.02 | 1323.45 | 0.06 |
| R CA1 gyrification | Stable AD | 0.0036 | 0.00 | 1.40 | 1336.53 | 0.20 |
| R CA1 gyrification | CN to MCI/AD | -0.0105 | 0.00 | -2.71 | 1291.16 | 0.01 |
| R CA1 gyrification | MCI to AD | -0.0008 | 0.00 | -0.30 | 1308.28 | 0.79 |
| R CA1 gyrification | Followup Years | 0.0003 | 0.00 | 2.51 | 706.73 | 0.02 |
| R CA1 gyrification | Age | 0.0039 | 9.68E-05 | 40.77 | 1820.68 | < 0.01 |
| R CA1 gyrification | Education | -0.0029 | 0.00 | -9.90 | 1309.48 | < 0.01 |
| R CA1 gyrification | Scanner Site | 0.0000 | 6.84E-06 | -4.59 | 1202.08 | < 0.01 |
| R CA1 gyrification | SexM | -0.0361 | 0.00 | -21.77 | 1314.64 | < 0.01 |
| R CA1 gyrification | Stable MCI:Followup Years | 0.0003 | 0.00 | 1.65 | 661.15 | 0.13 |
| R CA1 gyrification | Stable AD:Followup Years | 0.0008 | 0.00 | 2.17 | 888.84 | 0.04 |
| R CA1 gyrification | CN to MCI/AD:Followup Years | 0.0009 | 0.00 | 3.79 | 506.20 | < 0.01 |
| R CA1 gyrification | MCI to AD:Followup Years | 0.0010 | 0.00 | 5.21 | 774.66 | < 0.01 |
| R CA1 thickness | (Intercept) | -0.7091 | 0.01 | -78.43 | 1763.02 | < 0.01 |
| R CA1 thickness | Stable MCI | 0.0056 | 0.00 | 2.49 | 1322.26 | 0.02 |
| R CA1 thickness | Stable AD | 0.0059 | 0.00 | 2.28 | 1335.20 | 0.03 |
| R CA1 thickness | CN to MCI/AD | -0.0105 | 0.00 | -2.70 | 1290.25 | 0.01 |
| R CA1 thickness | MCI to AD | 0.0010 | 0.00 | 0.39 | 1307.20 | 0.74 |
| R CA1 thickness | Followup Years | 0.0003 | 0.00 | 2.49 | 707.06 | 0.02 |
| R CA1 thickness | Age | 0.0041 | 9.78E-05 | 41.56 | 1832.44 | < 0.01 |
| R CA1 thickness | Education | -0.0029 | 0.00 | -9.74 | 1309.68 | < 0.01 |

| | | | | | | |
|---|---|---|---|---|---|---|
| R CA1 thickness | Scanner Site | 0.0000 | 6.92E-06 | -4.62 | 1206.37 | < 0.01 |
| R CA1 thickness | SexM | -0.0366 | 0.00 | -21.83 | 1315.08 | < 0.01 |
| R CA1 thickness | Stable MCI:Followup Years | 0.0003 | 0.00 | 1.80 | 660.32 | 0.09 |
| R CA1 thickness | Stable AD:Followup Years | 0.0010 | 0.00 | 2.78 | 890.93 | 0.01 |
| R CA1 thickness | CN to MCI/AD:Followup Years | 0.0010 | 0.00 | 4.26 | 505.04 | < 0.01 |
| R CA1 thickness | MCI to AD:Followup Years | 0.0013 | 0.00 | 6.19 | 773.34 | < 0.01 |
| R CA2 volume | (Intercept) | -0.4107 | 0.07 | -5.73 | 1423.59 | < 0.01 |
| R CA2 volume | Stable MCI | -0.0032 | 0.02 | -0.20 | 1360.84 | 0.86 |
| R CA2 volume | Stable AD | 0.0606 | 0.02 | 3.21 | 1488.19 | < 0.01 |
| R CA2 volume | CN to MCI/AD | -0.0505 | 0.03 | -1.88 | 1122.69 | 0.08 |
| R CA2 volume | MCI to AD | 0.0205 | 0.02 | 1.12 | 1263.33 | 0.31 |
| R CA2 volume | Followup Years | -0.0014 | 0.00 | -0.58 | 334.17 | 0.61 |
| R CA2 volume | Age | 0.0074 | 0.00 | 9.11 | 1422.34 | < 0.01 |
| R CA2 volume | Education | -0.0020 | 0.00 | -0.95 | 1280.30 | 0.39 |
| R CA2 volume | Scanner Site | 0.0000 | 5.05E-05 | -0.58 | 1316.24 | 0.61 |
| R CA2 volume | SexM | 0.0314 | 0.01 | 2.44 | 1430.54 | 0.02 |
| R CA2 volume | ETIV | -0.0112 | 0.01 | -1.94 | 2343.08 | 0.07 |
| R CA2 volume | Stable MCI:Followup Years | 0.0046 | 0.00 | 1.29 | 349.10 | 0.24 |
| R CA2 volume | Stable AD:Followup Years | 0.0196 | 0.01 | 2.71 | 450.01 | 0.01 |
| R CA2 volume | CN to MCI/AD:Followup Years | 0.0169 | 0.00 | 3.61 | 268.12 | < 0.01 |
| R CA2 volume | MCI to AD:Followup Years | 0.0265 | 0.00 | 6.65 | 433.00 | < 0.01 |
| R CA2 curvature | (Intercept) | -0.7153 | 0.01 | -78.76 | 1764.69 | 0.00 |
| R CA2 curvature | Stable MCI | 0.0057 | 0.00 | 2.55 | 1322.33 | 0.02 |
| R CA2 curvature | Stable AD | 0.0062 | 0.00 | 2.39 | 1335.22 | 0.03 |
| R CA2 curvature | CN to MCI/AD | -0.0105 | 0.00 | -2.66 | 1290.49 | 0.01 |

| | | | | | | |
|---|---|---|---|---|---|---|
| R CA2 curvature | MCI to AD | 0.0013 | 0.00 | 0.49 | 1307.35 | 0.68 |
| R CA2 curvature | Followup Years | 0.0003 | 0.00 | 2.56 | 707.33 | 0.02 |
| R CA2 curvature | Age | 0.0041 | 9.82E-05 | 41.42 | 1834.18 | < 0.01 |
| R CA2 curvature | Education | -0.0029 | 0.00 | -9.70 | 1309.98 | < 0.01 |
| R CA2 curvature | Scanner Site | 0.0000 | 6.96E-06 | -4.60 | 1207.12 | < 0.01 |
| R CA2 curvature | SexM | -0.0369 | 0.00 | -21.91 | 1315.40 | < 0.01 |
| R CA2 curvature | Stable MCI:Followup Years | 0.0003 | 0.00 | 1.81 | 660.45 | 0.09 |
| R CA2 curvature | Stable AD:Followup Years | 0.0010 | 0.00 | 2.79 | 892.57 | 0.01 |
| R CA2 curvature | CN to MCI/AD:Followup Years | 0.0010 | 0.00 | 4.26 | 505.30 | < 0.01 |
| R CA2 curvature | MCI to AD:Followup Years | 0.0012 | 0.00 | 6.06 | 773.33 | < 0.01 |
| R CA2 gyrification | (Intercept) | -0.7121 | 0.01 | -78.59 | 1752.76 | < 0.01 |
| R CA2 gyrification | Stable MCI | 0.0056 | 0.00 | 2.50 | 1322.08 | 0.02 |
| R CA2 gyrification | Stable AD | 0.0062 | 0.00 | 2.40 | 1335.55 | 0.02 |
| R CA2 gyrification | CN to MCI/AD | -0.0108 | 0.00 | -2.76 | 1289.32 | 0.01 |
| R CA2 gyrification | MCI to AD | 0.0010 | 0.00 | 0.40 | 1306.75 | 0.73 |
| R CA2 gyrification | Followup Years | 0.0003 | 0.00 | 2.41 | 709.68 | 0.02 |
| R CA2 gyrification | Age | 0.0041 | 9.82E-05 | 41.64 | 1826.16 | < 0.01 |
| R CA2 gyrification | Education | -0.0029 | 0.00 | -9.73 | 1309.46 | < 0.01 |
| R CA2 gyrification | Scanner Site | 0.0000 | 6.91E-06 | -4.64 | 1204.53 | < 0.01 |
| R CA2 gyrification | SexM | -0.0366 | 0.00 | -21.90 | 1314.64 | < 0.01 |
| R CA2 gyrification | Stable MCI:Followup Years | 0.0003 | 0.00 | 1.81 | 664.40 | 0.09 |
| R CA2 gyrification | Stable AD:Followup Years | 0.0011 | 0.00 | 3.07 | 907.29 | 0.00 |
| R CA2 gyrification | CN to MCI/AD:Followup Years | 0.0011 | 0.00 | 4.48 | 508.48 | < 0.01 |
| R CA2 gyrification | MCI to AD:Followup Years | 0.0014 | 0.00 | 6.78 | 774.70 | < 0.01 |
| R CA2 thickness | (Intercept) | -0.7116 | 0.01 | -78.55 | 1760.88 | < 0.01 |

| | | | | | | |
|---|---|---|---|---|---|---|
| R CA2 thickness | Stable MCI | 0.0057 | 0.00 | 2.54 | 1322.23 | 0.02 |
| R CA2 thickness | Stable AD | 0.0063 | 0.00 | 2.41 | 1335.26 | 0.02 |
| R CA2 thickness | CN to MCI/AD | -0.0105 | 0.00 | -2.68 | 1290.05 | 0.01 |
| R CA2 thickness | MCI to AD | 0.0013 | 0.00 | 0.49 | 1307.11 | 0.67 |
| R CA2 thickness | Followup Years | 0.0003 | 0.00 | 2.49 | 709.51 | 0.02 |
| R CA2 thickness | Age | 0.0041 | 9.80E-05 | 41.63 | 1830.81 | < 0.01 |
| R CA2 thickness | Education | -0.0029 | 0.00 | -9.72 | 1309.55 | < 0.01 |
| R CA2 thickness | Scanner Site | 0.0000 | 6.93E-06 | -4.62 | 1205.33 | < 0.01 |
| R CA2 thickness | SexM | -0.0366 | 0.00 | -21.82 | 1314.87 | < 0.01 |
| R CA2 thickness | Stable MCI:Followup Years | 0.0003 | 0.00 | 1.84 | 662.97 | 0.09 |
| R CA2 thickness | Stable AD:Followup Years | 0.0010 | 0.00 | 2.88 | 894.18 | 0.01 |
| R CA2 thickness | CN to MCI/AD:Followup Years | 0.0010 | 0.00 | 4.30 | 507.29 | < 0.01 |
| R CA2 thickness | MCI to AD:Followup Years | 0.0013 | 0.00 | 6.27 | 776.09 | < 0.01 |
| R CA3 volume | (Intercept) | 1.1321 | 0.10 | 11.47 | 1442.33 | < 0.01 |
| R CA3 volume | Stable MCI | -0.1380 | 0.02 | -6.53 | 1323.62 | < 0.01 |
| R CA3 volume | Stable AD | -0.2948 | 0.02 | -11.81 | 1418.15 | < 0.01 |
| R CA3 volume | CN to MCI/AD | -0.1159 | 0.04 | -3.21 | 1157.12 | < 0.01 |
| R CA3 volume | MCI to AD | -0.3012 | 0.02 | -12.30 | 1253.68 | < 0.01 |
| R CA3 volume | Followup Years | 0.0013 | 0.00 | 0.46 | 281.20 | 0.69 |
| R CA3 volume | Age | -0.0035 | 0.00 | -3.08 | 1478.56 | < 0.01 |
| R CA3 volume | Education | 0.0032 | 0.00 | 1.12 | 1279.72 | 0.31 |
| R CA3 volume | Scanner Site | 3.00E-05 | 7.00E-05 | 0.43 | 1399.45 | 0.71 |
| R CA3 volume | SexM | 0.1179 | 0.02 | 6.66 | 1445.85 | < 0.01 |
| R CA3 volume | ETIV | -0.0471 | 0.01 | -6.43 | 2768.19 | < 0.01 |
| R CA3 volume | Stable MCI:Followup Years | -0.0059 | 0.00 | -1.37 | 290.30 | 0.21 |

| | | | | | | |
|---|---|---|---|---|---|---|
| R CA3 volume | Stable AD:Followup Years | 0.0172 | 0.01 | 1.95 | 509.76 | 0.07 |
| R CA3 volume | CN to MCI/AD:Followup Years | -0.0015 | 0.01 | -0.27 | 207.25 | 0.81 |
| R CA3 volume | MCI to AD:Followup Years | 0.0032 | 0.00 | 0.67 | 337.65 | 0.56 |
| R CA3 curvature | (Intercept) | -0.7161 | 0.01 | -78.85 | 1764.70 | < 0.01 |
| R CA3 curvature | Stable MCI | 0.0057 | 0.00 | 2.56 | 1322.20 | 0.02 |
| R CA3 curvature | Stable AD | 0.0063 | 0.00 | 2.42 | 1335.09 | 0.02 |
| R CA3 curvature | CN to MCI/AD | -0.0105 | 0.00 | -2.66 | 1290.36 | 0.01 |
| R CA3 curvature | MCI to AD | 0.0013 | 0.00 | 0.50 | 1307.22 | 0.67 |
| R CA3 curvature | Followup Years | 0.0003 | 0.00 | 2.54 | 707.57 | 0.02 |
| R CA3 curvature | Age | 0.0041 | 9.82E-05 | 41.52 | 1834.67 | < 0.01 |
| R CA3 curvature | Education | -0.0029 | 0.00 | -9.70 | 1309.83 | < 0.01 |
| R CA3 curvature | Scanner Site | 0.0000 | 6.96E-06 | -4.60 | 1207.40 | < 0.01 |
| R CA3 curvature | SexM | -0.0369 | 0.00 | -21.92 | 1315.27 | < 0.01 |
| R CA3 curvature | Stable MCI:Followup Years | 0.0003 | 0.00 | 1.82 | 660.60 | 0.09 |
| R CA3 curvature | Stable AD:Followup Years | 0.0010 | 0.00 | 2.78 | 893.35 | 0.01 |
| R CA3 curvature | CN to MCI/AD:Followup Years | 0.0010 | 0.00 | 4.24 | 505.39 | < 0.01 |
| R CA3 curvature | MCI to AD:Followup Years | 0.0012 | 0.00 | 6.15 | 773.41 | < 0.01 |
| R CA3 gyrification | (Intercept) | -0.7043 | 0.01 | -77.83 | 1747.34 | < 0.01 |
| R CA3 gyrification | Stable MCI | 0.0049 | 0.00 | 2.19 | 1322.29 | 0.04 |
| R CA3 gyrification | Stable AD | 0.0047 | 0.00 | 1.83 | 1336.00 | 0.09 |
| R CA3 gyrification | CN to MCI/AD | -0.0109 | 0.00 | -2.80 | 1288.64 | 0.01 |
| R CA3 gyrification | MCI to AD | -0.0004 | 0.00 | -0.14 | 1306.56 | 0.90 |
| R CA3 gyrification | Followup Years | 0.0003 | 0.00 | 2.40 | 709.77 | 0.02 |
| R CA3 gyrification | Age | 0.0040 | 9.82E-05 | 41.21 | 1821.76 | < 0.01 |
| R CA3 gyrification | Education | -0.0029 | 0.00 | -9.71 | 1309.25 | < 0.01 |

| | | | | | | |
|---|---|---|---|---|---|---|
| R CA3 gyrification | Scanner Site | 0.0000 | 6.88E-06 | -4.63 | 1206.59 | < 0.01 |
| R CA3 gyrification | SexM | -0.0363 | 0.00 | -21.76 | 1314.48 | < 0.01 |
| R CA3 gyrification | Stable MCI:Followup Years | 0.0003 | 0.00 | 1.64 | 664.95 | 0.13 |
| R CA3 gyrification | Stable AD:Followup Years | 0.0011 | 0.00 | 3.02 | 903.24 | 0.01 |
| R CA3 gyrification | CN to MCI/AD:Followup Years | 0.0010 | 0.00 | 4.07 | 509.06 | < 0.01 |
| R CA3 gyrification | MCI to AD:Followup Years | 0.0013 | 0.00 | 6.13 | 776.51 | < 0.01 |
| R CA3 thickness | (Intercept) | -0.7079 | 0.01 | -78.16 | 1760.32 | < 0.01 |
| R CA3 thickness | Stable MCI | 0.0055 | 0.00 | 2.45 | 1322.43 | 0.02 |
| R CA3 thickness | Stable AD | 0.0057 | 0.00 | 2.18 | 1335.46 | 0.04 |
| R CA3 thickness | CN to MCI/AD | -0.0106 | 0.00 | -2.70 | 1290.26 | 0.01 |
| R CA3 thickness | MCI to AD | 0.0008 | 0.00 | 0.32 | 1307.31 | 0.78 |
| R CA3 thickness | Followup Years | 0.0003 | 0.00 | 2.60 | 709.03 | 0.02 |
| R CA3 thickness | Age | 0.0041 | 9.80E-05 | 41.37 | 1829.35 | < 0.01 |
| R CA3 thickness | Education | -0.0029 | 0.00 | -9.69 | 1309.69 | < 0.01 |
| R CA3 thickness | Scanner Site | 0.0000 | 6.93E-06 | -4.59 | 1205.12 | < 0.01 |
| R CA3 thickness | SexM | -0.0366 | 0.00 | -21.80 | 1315.00 | < 0.01 |
| R CA3 thickness | Stable MCI:Followup Years | 0.0003 | 0.00 | 1.77 | 662.67 | 0.10 |
| R CA3 thickness | Stable AD:Followup Years | 0.0010 | 0.00 | 2.70 | 893.18 | 0.01 |
| R CA3 thickness | CN to MCI/AD:Followup Years | 0.0010 | 0.00 | 4.21 | 507.29 | < 0.01 |
| R CA3 thickness | MCI to AD:Followup Years | 0.0012 | 0.00 | 5.88 | 775.93 | < 0.01 |
| R CA4 volume | (Intercept) | 0.5720 | 0.08 | 7.45 | 1477.21 | < 0.01 |
| R CA4 volume | Stable MCI | -0.1296 | 0.02 | -7.80 | 1347.90 | < 0.01 |
| R CA4 volume | Stable AD | -0.2472 | 0.02 | -12.62 | 1432.10 | < 0.01 |
| R CA4 volume | CN to MCI/AD | -0.0345 | 0.03 | -1.21 | 1194.08 | 0.27 |
| R CA4 volume | MCI to AD | -0.2224 | 0.02 | -11.53 | 1283.14 | < 0.01 |

| | | | | | | |
|---|---|---|---|---|---|---|
| R CA4 volume | Followup Years | -0.0033 | 0.00 | -1.48 | 313.14 | 0.17 |
| R CA4 volume | Age | -0.0011 | 0.00 | -1.29 | 1513.29 | 0.24 |
| R CA4 volume | Education | -0.0018 | 0.00 | -0.82 | 1311.00 | 0.46 |
| R CA4 volume | Scanner Site | 3.10E-06 | 5.44E-05 | 0.06 | 1411.91 | 0.96 |
| R CA4 volume | SexM | 0.0050 | 0.01 | 0.37 | 1483.03 | 0.75 |
| R CA4 volume | ETIV | 0.0047 | 0.01 | 0.83 | 2956.19 | 0.45 |
| R CA4 volume | Stable MCI:Followup Years | -0.0005 | 0.00 | -0.14 | 322.32 | 0.90 |
| R CA4 volume | Stable AD:Followup Years | 0.0067 | 0.01 | 1.01 | 556.34 | 0.36 |
| R CA4 volume | CN to MCI/AD:Followup Years | -0.0056 | 0.00 | -1.26 | 235.57 | 0.25 |
| R CA4 volume | MCI to AD:Followup Years | -0.0031 | 0.00 | -0.85 | 375.38 | 0.45 |
| R CA4 curvature | (Intercept) | -0.7188 | 0.01 | -76.97 | 1714.30 | 0.00 |
| R CA4 curvature | Stable MCI | 0.0064 | 0.00 | 2.80 | 1326.08 | 0.01 |
| R CA4 curvature | Stable AD | 0.0085 | 0.00 | 3.21 | 1341.08 | 0.00 |
| R CA4 curvature | CN to MCI/AD | -0.0104 | 0.00 | -2.61 | 1286.22 | 0.02 |
| R CA4 curvature | MCI to AD | 0.0024 | 0.00 | 0.90 | 1307.57 | 0.42 |
| R CA4 curvature | Followup Years | 0.0003 | 0.00 | 2.42 | 644.70 | 0.02 |
| R CA4 curvature | Age | 0.0041 | 0.00 | 40.04 | 1773.97 | < 0.01 |
| R CA4 curvature | Education | -0.0028 | 0.00 | -9.44 | 1311.56 | < 0.01 |
| R CA4 curvature | Scanner Site | 0.0000 | 7.04E-06 | -4.55 | 1212.11 | < 0.01 |
| R CA4 curvature | SexM | -0.0364 | 0.00 | -21.36 | 1316.84 | < 0.01 |
| R CA4 curvature | Stable MCI:Followup Years | 0.0004 | 0.00 | 1.88 | 607.69 | 0.08 |
| R CA4 curvature | Stable AD:Followup Years | 0.0009 | 0.00 | 2.27 | 783.24 | 0.03 |
| R CA4 curvature | CN to MCI/AD:Followup Years | 0.0010 | 0.00 | 3.94 | 460.78 | < 0.01 |
| R CA4 curvature | MCI to AD:Followup Years | 0.0015 | 0.00 | 6.64 | 722.58 | < 0.01 |
| R CA4 gyrification | (Intercept) | -0.6743 | 0.01 | -74.71 | 1699.41 | 0.00 |

| | | | | | | |
|---|---|---|---|---|---|---|
| R CA4 gyrification | Stable MCI | 0.0030 | 0.00 | 1.39 | 1324.30 | 0.20 |
| R CA4 gyrification | Stable AD | 0.0021 | 0.00 | 0.84 | 1340.55 | 0.45 |
| R CA4 gyrification | CN to MCI/AD | -0.0105 | 0.00 | -2.76 | 1284.24 | 0.01 |
| R CA4 gyrification | MCI to AD | -0.0026 | 0.00 | -1.01 | 1305.87 | 0.36 |
| R CA4 gyrification | Followup Years | 0.0003 | 0.00 | 1.80 | 653.75 | 0.09 |
| R CA4 gyrification | Age | 0.0039 | 9.89E-05 | 39.87 | 1775.14 | < 0.01 |
| R CA4 gyrification | Education | -0.0029 | 0.00 | -10.06 | 1309.60 | < 0.01 |
| R CA4 gyrification | Scanner Site | 0.0000 | 6.75E-06 | -4.58 | 1224.44 | < 0.01 |
| R CA4 gyrification | SexM | -0.0353 | 0.00 | -21.61 | 1315.10 | < 0.01 |
| R CA4 gyrification | Stable MCI:Followup Years | 0.0003 | 0.00 | 1.49 | 618.64 | 0.17 |
| R CA4 gyrification | Stable AD:Followup Years | 0.0011 | 0.00 | 2.68 | 862.88 | 0.01 |
| R CA4 gyrification | CN to MCI/AD:Followup Years | 0.0010 | 0.00 | 3.54 | 472.15 | 0.00 |
| R CA4 gyrification | MCI to AD:Followup Years | 0.0013 | 0.00 | 5.65 | 721.81 | < 0.01 |
| R CA4 thickness | (Intercept) | -0.7037 | 0.01 | -77.79 | 1758.74 | < 0.01 |
| R CA4 thickness | Stable MCI | 0.0052 | 0.00 | 2.33 | 1322.37 | 0.03 |
| R CA4 thickness | Stable AD | 0.0050 | 0.00 | 1.93 | 1335.46 | 0.07 |
| R CA4 thickness | CN to MCI/AD | -0.0107 | 0.00 | -2.73 | 1290.19 | 0.01 |
| R CA4 thickness | MCI to AD | 0.0003 | 0.00 | 0.13 | 1307.26 | 0.91 |
| R CA4 thickness | Followup Years | 0.0003 | 0.00 | 2.56 | 709.48 | 0.02 |
| R CA4 thickness | Age | 0.0040 | 9.79E-05 | 41.27 | 1827.74 | < 0.01 |
| R CA4 thickness | Education | -0.0029 | 0.00 | -9.70 | 1309.74 | < 0.01 |
| R CA4 thickness | Scanner Site | 0.0000 | 6.92E-06 | -4.58 | 1204.31 | < 0.01 |
| R CA4 thickness | SexM | -0.0366 | 0.00 | -21.81 | 1314.99 | < 0.01 |
| R CA4 thickness | Stable MCI:Followup Years | 0.0003 | 0.00 | 1.72 | 663.39 | 0.11 |
| R CA4 thickness | Stable AD:Followup Years | 0.0010 | 0.00 | 2.64 | 895.90 | 0.01 |

| | | | | | | |
|---|---|---|---|---|---|---|
| R CA4 thickness | CN to MCI/AD:Followup Years | 0.0010 | 0.00 | 4.10 | 507.96 | < 0.01 |
| R CA4 thickness | MCI to AD:Followup Years | 0.0012 | 0.00 | 5.70 | 776.17 | < 0.01 |
| R DG volume | (Intercept) | 0.5664 | 0.05 | 12.24 | 1549.07 | < 0.01 |
| R DG volume | Stable MCI | -0.1021 | 0.01 | -10.10 | 1303.99 | < 0.01 |
| R DG volume | Stable AD | -0.2554 | 0.01 | -21.61 | 1331.46 | < 0.01 |
| R DG volume | CN to MCI/AD | -0.0391 | 0.02 | -2.21 | 1257.00 | 0.04 |
| R DG volume | MCI to AD | -0.2075 | 0.01 | -17.55 | 1283.91 | < 0.01 |
| R DG volume | Followup Years | -0.0010 | 0.00 | -0.84 | 469.57 | 0.45 |
| R DG volume | Age | -0.0056 | 0.00 | -10.68 | 1649.17 | < 0.01 |
| R DG volume | Education | -0.0014 | 0.00 | -1.00 | 1290.04 | 0.37 |
| R DG volume | Scanner Site | 3.79E-05 | 3.32E-05 | 1.14 | 1326.46 | 0.30 |
| R DG volume | SexM | 0.0252 | 0.01 | 3.06 | 1449.23 | < 0.01 |
| R DG volume | ETIV | -0.0366 | 0.00 | -14.36 | 4127.09 | < 0.01 |
| R DG volume | Stable MCI:Followup Years | -0.0047 | 0.00 | -2.91 | 443.46 | 0.01 |
| R DG volume | Stable AD:Followup Years | -0.0124 | 0.00 | -4.33 | 1330.96 | < 0.01 |
| R DG volume | CN to MCI/AD:Followup Years | -0.0097 | 0.00 | -4.24 | 329.37 | < 0.01 |
| R DG volume | MCI to AD:Followup Years | -0.0171 | 0.00 | -9.65 | 464.89 | < 0.01 |
| R DG curvature | (Intercept) | -0.7128 | 0.01 | -78.43 | 1759.01 | < 0.01 |
| R DG curvature | Stable MCI | 0.0054 | 0.00 | 2.40 | 1322.94 | 0.02 |
| R DG curvature | Stable AD | 0.0054 | 0.00 | 2.07 | 1335.93 | 0.05 |
| R DG curvature | CN to MCI/AD | -0.0108 | 0.00 | -2.74 | 1290.63 | 0.01 |
| R DG curvature | MCI to AD | 0.0007 | 0.00 | 0.28 | 1307.75 | 0.81 |
| R DG curvature | Followup Years | 0.0003 | 0.00 | 2.55 | 703.67 | 0.02 |
| R DG curvature | Age | 0.0040 | 9.83E-05 | 41.08 | 1824.70 | < 0.01 |
| R DG curvature | Education | -0.0029 | 0.00 | -9.70 | 1310.17 | < 0.01 |

| | | | | | | |
|---|---|---|---|---|---|---|
| R DG curvature | Scanner Site | 0.0000 | 6.96E-06 | -4.55 | 1205.30 | < 0.01 |
| R DG curvature | SexM | -0.0371 | 0.00 | -21.99 | 1315.48 | < 0.01 |
| R DG curvature | Stable MCI:Followup Years | 0.0003 | 0.00 | 1.69 | 657.89 | 0.11 |
| R DG curvature | Stable AD:Followup Years | 0.0010 | 0.00 | 2.73 | 880.89 | 0.01 |
| R DG curvature | CN to MCI/AD:Followup Years | 0.0010 | 0.00 | 4.24 | 503.54 | < 0.01 |
| R DG curvature | MCI to AD:Followup Years | 0.0012 | 0.00 | 5.85 | 772.06 | < 0.01 |
| R DG gyrification | (Intercept) | -0.6667 | 0.01 | -73.67 | 1713.07 | < 0.01 |
| R DG gyrification | Stable MCI | 0.0021 | 0.00 | 0.93 | 1325.84 | 0.40 |
| R DG gyrification | Stable AD | -0.0017 | 0.00 | -0.67 | 1340.83 | 0.56 |
| R DG gyrification | CN to MCI/AD | -0.0114 | 0.00 | -2.95 | 1289.53 | 0.01 |
| R DG gyrification | MCI to AD | -0.0058 | 0.00 | -2.22 | 1309.04 | 0.04 |
| R DG gyrification | Followup Years | 0.0003 | 0.00 | 2.35 | 706.96 | 0.03 |
| R DG gyrification | Age | 0.0038 | 9.86E-05 | 38.30 | 1781.76 | < 0.01 |
| R DG gyrification | Education | -0.0029 | 0.00 | -9.98 | 1307.35 | < 0.01 |
| R DG gyrification | Scanner Site | 0.0000 | 6.82E-06 | -4.32 | 1190.92 | < 0.01 |
| R DG gyrification | SexM | -0.0355 | 0.00 | -21.45 | 1311.66 | < 0.01 |
| R DG gyrification | Stable MCI:Followup Years | 0.0003 | 0.00 | 1.51 | 667.48 | 0.16 |
| R DG gyrification | Stable AD:Followup Years | 0.0008 | 0.00 | 2.08 | 905.44 | 0.05 |
| R DG gyrification | CN to MCI/AD:Followup Years | 0.0007 | 0.00 | 2.78 | 510.39 | 0.01 |
| R DG gyrification | MCI to AD:Followup Years | 0.0008 | 0.00 | 3.57 | 776.88 | < 0.01 |
| R Sub volume | (Intercept) | 2.4209 | 0.11 | 21.31 | 1518.81 | < 0.01 |
| R Sub volume | Stable MCI | -0.1502 | 0.03 | -5.89 | 1321.38 | < 0.01 |
| R Sub volume | Stable AD | -0.3535 | 0.03 | -11.84 | 1358.47 | < 0.01 |
| R Sub volume | CN to MCI/AD | -0.0463 | 0.04 | -1.05 | 1230.24 | 0.34 |
| R Sub volume | MCI to AD | -0.2525 | 0.03 | -8.49 | 1281.15 | < 0.01 |

| | | | | | | |
|---|---|---|---|---|---|---|
| R Sub volume | Followup Years | 0.0001 | 0.00 | 0.06 | 455.76 | 0.96 |
| R Sub volume | Age | -0.0107 | 0.00 | -8.33 | 1576.41 | < 0.01 |
| R Sub volume | Education | 0.0015 | 0.00 | 0.45 | 1289.03 | 0.70 |
| R Sub volume | Scanner Site | 0.0001 | 8.12E-05 | 1.35 | 1300.46 | 0.21 |
| R Sub volume | SexM | 0.1244 | 0.02 | 6.05 | 1470.89 | < 0.01 |
| R Sub volume | ETIV | 0.0486 | 0.01 | 6.30 | 3871.69 | < 0.01 |
| R Sub volume | Stable MCI:Followup Years | 0.0000 | 0.00 | 0.00 | 455.15 | 1.00 |
| R Sub volume | Stable AD:Followup Years | -0.0282 | 0.01 | -3.81 | 652.03 | 0.00 |
| R Sub volume | CN to MCI/AD:Followup Years | -0.0232 | 0.00 | -4.79 | 337.63 | < 0.01 |
| R Sub volume | MCI to AD:Followup Years | -0.0334 | 0.00 | -8.16 | 541.78 | < 0.01 |
| R Sub curvature | (Intercept) | -0.7162 | 0.01 | -78.85 | 1761.21 | 0.00 |
| R Sub curvature | Stable MCI | 0.0057 | 0.00 | 2.53 | 1322.41 | 0.02 |
| R Sub curvature | Stable AD | 0.0061 | 0.00 | 2.34 | 1335.38 | 0.03 |
| R Sub curvature | CN to MCI/AD | -0.0105 | 0.00 | -2.67 | 1290.15 | 0.01 |
| R Sub curvature | MCI to AD | 0.0012 | 0.00 | 0.45 | 1307.24 | 0.70 |
| R Sub curvature | Followup Years | 0.0003 | 0.00 | 2.54 | 706.37 | 0.02 |
| R Sub curvature | Age | 0.0041 | 9.83E-05 | 41.42 | 1829.23 | < 0.01 |
| R Sub curvature | Education | -0.0029 | 0.00 | -9.71 | 1309.59 | < 0.01 |
| R Sub curvature | Scanner Site | 0.0000 | 6.96E-06 | -4.60 | 1205.78 | < 0.01 |
| R Sub curvature | SexM | -0.0369 | 0.00 | -21.92 | 1314.96 | < 0.01 |
| R Sub curvature | Stable MCI:Followup Years | 0.0003 | 0.00 | 1.79 | 659.92 | 0.09 |
| R Sub curvature | Stable AD:Followup Years | 0.0010 | 0.00 | 2.73 | 884.98 | 0.01 |
| R Sub curvature | CN to MCI/AD:Followup Years | 0.0010 | 0.00 | 4.24 | 504.84 | < 0.01 |
| R Sub curvature | MCI to AD:Followup Years | 0.0012 | 0.00 | 5.95 | 774.04 | < 0.01 |
| R Sub gyrification | (Intercept) | -0.7021 | 0.01 | -77.89 | 1759.32 | < 0.01 |

| | | | | | | |
|---|---|---|---|---|---|---|
| R Sub gyrification | Stable MCI | 0.0050 | 0.00 | 2.22 | 1323.24 | 0.04 |
| R Sub gyrification | Stable AD | 0.0045 | 0.00 | 1.73 | 1336.29 | 0.11 |
| R Sub gyrification | CN to MCI/AD | -0.0105 | 0.00 | -2.70 | 1290.89 | 0.01 |
| R Sub gyrification | MCI to AD | 0.0000 | 0.00 | -0.01 | 1308.04 | 1.00 |
| R Sub gyrification | Followup Years | 0.0003 | 0.00 | 2.63 | 708.00 | 0.01 |
| R Sub gyrification | Age | 0.0040 | 9.75E-05 | 40.77 | 1826.26 | < 0.01 |
| R Sub gyrification | Education | -0.0029 | 0.00 | -9.78 | 1310.51 | < 0.01 |
| R Sub gyrification | Scanner Site | 0.0000 | 6.90E-06 | -4.57 | 1204.54 | < 0.01 |
| R Sub gyrification | SexM | -0.0364 | 0.00 | -21.80 | 1315.76 | < 0.01 |
| R Sub gyrification | Stable MCI:Followup Years | 0.0003 | 0.00 | 1.74 | 662.03 | 0.10 |
| R Sub gyrification | Stable AD:Followup Years | 0.0009 | 0.00 | 2.42 | 887.29 | 0.02 |
| R Sub gyrification | CN to MCI/AD:Followup Years | 0.0009 | 0.00 | 3.86 | 506.67 | < 0.01 |
| R Sub gyrification | MCI to AD:Followup Years | 0.0011 | 0.00 | 5.26 | 776.17 | < 0.01 |
| R Sub thickness | (Intercept) | -0.7097 | 0.01 | -78.50 | 1763.62 | < 0.01 |
| R Sub thickness | Stable MCI | 0.0057 | 0.00 | 2.53 | 1322.31 | 0.02 |
| R Sub thickness | Stable AD | 0.0061 | 0.00 | 2.35 | 1335.23 | 0.03 |
| R Sub thickness | CN to MCI/AD | -0.0105 | 0.00 | -2.68 | 1290.32 | 0.01 |
| R Sub thickness | MCI to AD | 0.0012 | 0.00 | 0.46 | 1307.26 | 0.69 |
| R Sub thickness | Followup Years | 0.0003 | 0.00 | 2.55 | 705.58 | 0.02 |
| R Sub thickness | Age | 0.0041 | 9.78E-05 | 41.62 | 1832.34 | < 0.01 |
| R Sub thickness | Education | -0.0029 | 0.00 | -9.72 | 1309.99 | < 0.01 |
| R Sub thickness | Scanner Site | 0.0000 | 6.93E-06 | -4.62 | 1206.56 | < 0.01 |
| R Sub thickness | SexM | -0.0365 | 0.00 | -21.79 | 1315.39 | < 0.01 |
| R Sub thickness | Stable MCI:Followup Years | 0.0003 | 0.00 | 1.83 | 658.97 | 0.09 |
| R Sub thickness | Stable AD:Followup Years | 0.0010 | 0.00 | 2.74 | 888.40 | 0.01 |

| | | | | | | |
|---|---|---|---|---|---|---|
| R Sub thickness | CN to MCI/AD:Followup Years | 0.0010 | 0.00 | 4.23 | 504.00 | < 0.01 |
| R Sub thickness | MCI to AD:Followup Years | 0.0012 | 0.00 | 6.14 | 772.01 | < 0.01 |
| R SRLM volume | (Intercept) | 4.1240 | 0.15 | 27.54 | 1592.99 | < 0.01 |
| R SRLM volume | Stable MCI | -0.3388 | 0.03 | -10.25 | 1302.56 | < 0.01 |
| R SRLM volume | Stable AD | -0.7887 | 0.04 | -20.42 | 1323.90 | < 0.01 |
| R SRLM volume | CN to MCI/AD | -0.1262 | 0.06 | -2.18 | 1266.45 | 0.04 |
| R SRLM volume | MCI to AD | -0.6670 | 0.04 | -17.23 | 1287.09 | < 0.01 |
| R SRLM volume | Followup Years | -0.0039 | 0.00 | -1.12 | 564.08 | 0.31 |
| R SRLM volume | Age | -0.0253 | 0.00 | -15.05 | 1718.82 | < 0.01 |
| R SRLM volume | Education | 0.0021 | 0.00 | 0.46 | 1289.19 | 0.69 |
| R SRLM volume | Scanner Site | 0.0001 | 0.00 | 1.17 | 1316.26 | 0.28 |
| R SRLM volume | SexM | 0.1812 | 0.03 | 6.78 | 1433.93 | < 0.01 |
| R SRLM volume | ETIV | 0.0007 | 0.01 | 0.09 | 4079.56 | 0.94 |
| R SRLM volume | Stable MCI:Followup Years | -0.0122 | 0.00 | -2.47 | 520.69 | 0.02 |
| R SRLM volume | Stable AD:Followup Years | -0.0372 | 0.01 | -4.37 | 1551.52 | < 0.01 |
| R SRLM volume | CN to MCI/AD:Followup Years | -0.0248 | 0.01 | -3.51 | 391.44 | < 0.01 |
| R SRLM volume | MCI to AD:Followup Years | -0.0466 | 0.01 | -8.61 | 539.77 | < 0.01 |
| R Cyst volume | (Intercept) | -0.4780 | 0.03 | -16.84 | 1779.33 | < 0.01 |
| R Cyst volume | Stable MCI | -0.0175 | 0.01 | -2.50 | 1319.57 | 0.02 |
| R Cyst volume | Stable AD | -0.0402 | 0.01 | -4.93 | 1331.12 | < 0.01 |
| R Cyst volume | CN to MCI/AD | -0.0025 | 0.01 | -0.20 | 1298.03 | 0.86 |
| R Cyst volume | MCI to AD | -0.0194 | 0.01 | -2.36 | 1309.49 | 0.03 |
| R Cyst volume | Followup Years | 0.0003 | 0.00 | 0.60 | 494.11 | 0.60 |
| R Cyst volume | Age | 0.0021 | 0.00 | 6.83 | 1845.81 | < 0.01 |
| R Cyst volume | Education | -0.0020 | 0.00 | -2.18 | 1335.82 | 0.04 |

| | | | | | | |
|---|---|---|---|---|---|---|
| R Cyst volume | Scanner Site | 0.0000 | 2.18E-05 | -0.96 | 1270.54 | 0.39 |
| R Cyst volume | SexM | -0.0195 | 0.01 | -3.62 | 1446.80 | < 0.01 |
| R Cyst volume | ETIV | -0.0293 | 0.00 | -22.97 | 4148.84 | < 0.01 |
| R Cyst volume | Stable MCI:Followup Years | 8.33E-05 | 0.00 | 0.14 | 455.85 | 0.90 |
| R Cyst volume | Stable AD:Followup Years | -0.0022 | 0.00 | -1.94 | 905.52 | 0.07 |
| R Cyst volume | CN to MCI/AD:Followup Years | -0.0018 | 0.00 | -2.18 | 350.16 | 0.04 |
| R Cyst volume | MCI to AD:Followup Years | -0.0043 | 0.00 | -6.58 | 511.85 | < 0.01 |

**Table S2**

*Random Effects from Linear Mixed-Effects Models*

| Model | Term | Estimate |
|---|---|---|
| L CA1 volume | sd (Intercept) | 0.5751 |
| L CA1 volume | cor (Intercept).TimeSinceBL | 0.0227 |
| L CA1 volume | sd TimeSinceBL | 0.0369 |
| L CA1 volume | sd Observation | 0.2742 |
| L CA1 curvature | sd (Intercept) | 0.0302 |
| L CA1 curvature | cor (Intercept).TimeSinceBL | -0.4148 |
| L CA1 curvature | sd TimeSinceBL | 0.0012 |
| L CA1 curvature | sd Observation | 0.0088 |
| L CA1 gyrification | sd (Intercept) | 0.0298 |
| L CA1 gyrification | cor (Intercept).TimeSinceBL | -0.4148 |
| L CA1 gyrification | sd TimeSinceBL | 0.0012 |
| L CA1 gyrification | sd Observation | 0.0088 |
| L CA1 thickness | sd (Intercept) | 0.0301 |
| L CA1 thickness | cor (Intercept).TimeSinceBL | -0.4155 |
| L CA1 thickness | sd TimeSinceBL | 0.0012 |

| | | |
|---|---|---|
| L CA1 thickness | sd Observation | 0.0088 |
| L CA2 volume | sd (Intercept) | 0.1950 |
| L CA2 volume | cor (Intercept).TimeSinceBL | 0.5932 |
| L CA2 volume | sd TimeSinceBL | 0.0226 |
| L CA2 volume | sd Observation | 0.2037 |
| L CA2 curvature | sd (Intercept) | 0.0302 |
| L CA2 curvature | cor (Intercept).TimeSinceBL | -0.4161 |
| L CA2 curvature | sd TimeSinceBL | 0.0012 |
| L CA2 curvature | sd Observation | 0.0088 |
| L CA2 gyrification | sd (Intercept) | 0.0300 |
| L CA2 gyrification | cor (Intercept).TimeSinceBL | -0.4107 |
| L CA2 gyrification | sd TimeSinceBL | 0.0012 |
| L CA2 gyrification | sd Observation | 0.0089 |
| L CA2 thickness | sd (Intercept) | 0.0301 |
| L CA2 thickness | cor (Intercept).TimeSinceBL | -0.4155 |
| L CA2 thickness | sd TimeSinceBL | 0.0012 |
| L CA2 thickness | sd Observation | 0.0088 |
| L CA3 volume | sd (Intercept) | 0.3467 |
| L CA3 volume | cor (Intercept).TimeSinceBL | 0.0486 |
| L CA3 volume | sd TimeSinceBL | 0.0347 |
| L CA3 volume | sd Observation | 0.2434 |
| L CA3 curvature | sd (Intercept) | 0.0302 |
| L CA3 curvature | cor (Intercept).TimeSinceBL | -0.4178 |
| L CA3 curvature | sd TimeSinceBL | 0.0012 |
| L CA3 curvature | sd Observation | 0.0088 |

| | | |
|---|---|---|
| L CA3 gyrification | sd (Intercept) | 0.0298 |
| L CA3 gyrification | cor (Intercept).TimeSinceBL | -0.4254 |
| L CA3 gyrification | sd TimeSinceBL | 0.0012 |
| L CA3 gyrification | sd Observation | 0.0090 |
| L CA3 thickness | sd (Intercept) | 0.0301 |
| L CA3 thickness | cor (Intercept).TimeSinceBL | -0.4198 |
| L CA3 thickness | sd TimeSinceBL | 0.0012 |
| L CA3 thickness | sd Observation | 0.0088 |
| L CA4 volume | sd (Intercept) | 0.2097 |
| L CA4 volume | cor (Intercept).TimeSinceBL | -0.1286 |
| L CA4 volume | sd TimeSinceBL | 0.0193 |
| L CA4 volume | sd Observation | 0.1526 |
| L CA4 curvature | sd (Intercept) | 0.0303 |
| L CA4 curvature | cor (Intercept).TimeSinceBL | -0.4541 |
| L CA4 curvature | sd TimeSinceBL | 0.0013 |
| L CA4 curvature | sd Observation | 0.0103 |
| L CA4 gyrification | sd (Intercept) | 0.0293 |
| L CA4 gyrification | cor (Intercept).TimeSinceBL | -0.4140 |
| L CA4 gyrification | sd TimeSinceBL | 0.0013 |
| L CA4 gyrification | sd Observation | 0.0097 |
| L CA4 thickness | sd (Intercept) | 0.0301 |
| L CA4 thickness | cor (Intercept).TimeSinceBL | -0.4265 |
| L CA4 thickness | sd TimeSinceBL | 0.0012 |
| L CA4 thickness | sd Observation | 0.0089 |
| L DG volume | sd (Intercept) | 0.1297 |

| | | |
|---|---|---|
| L DG volume | cor (Intercept).TimeSinceBL | 0.0059 |
| L DG volume | sd TimeSinceBL | 0.0166 |
| L DG volume | sd Observation | 0.0575 |
| L DG curvature | sd (Intercept) | 0.0299 |
| L DG curvature | cor (Intercept).TimeSinceBL | -0.2983 |
| L DG curvature | sd TimeSinceBL | 0.0013 |
| L DG curvature | sd Observation | 0.0102 |
| L DG gyrification | sd (Intercept) | 0.0296 |
| L DG gyrification | cor (Intercept).TimeSinceBL | -0.4236 |
| L DG gyrification | sd TimeSinceBL | 0.0013 |
| L DG gyrification | sd Observation | 0.0093 |
| L Sub volume | sd (Intercept) | 0.3595 |
| L Sub volume | cor (Intercept).TimeSinceBL | 0.0065 |
| L Sub volume | sd TimeSinceBL | 0.0116 |
| L Sub volume | sd Observation | 0.0802 |
| L Sub curvature | sd (Intercept) | 0.0264 |
| L Sub curvature | cor (Intercept).TimeSinceBL | 0.0230 |
| L Sub curvature | sd TimeSinceBL | 0.0041 |
| L Sub curvature | sd Observation | 0.0225 |
| L Sub gyrification | sd (Intercept) | 0.0299 |
| L Sub gyrification | cor (Intercept).TimeSinceBL | -0.4203 |
| L Sub gyrification | sd TimeSinceBL | 0.0012 |
| L Sub gyrification | sd Observation | 0.0088 |
| L Sub thickness | sd (Intercept) | 0.0300 |
| L Sub thickness | cor (Intercept).TimeSinceBL | -0.4049 |

| | | |
|---|---|---|
| L Sub thickness | sd TimeSinceBL | 0.0013 |
| L Sub thickness | sd Observation | 0.0080 |
| L SRLM volume | sd (Intercept) | 0.4151 |
| L SRLM volume | cor (Intercept).TimeSinceBL | 0.0459 |
| L SRLM volume | sd TimeSinceBL | 0.0435 |
| L SRLM volume | sd Observation | 0.1739 |
| L Cyst volume | sd (Intercept) | 0.0819 |
| L Cyst volume | cor (Intercept).TimeSinceBL | -0.3447 |
| L Cyst volume | sd TimeSinceBL | 0.0037 |
| L Cyst volume | sd Observation | 0.0214 |
| R CA1 volume | sd (Intercept) | 0.6010 |
| R CA1 volume | cor (Intercept).TimeSinceBL | -0.0411 |
| R CA1 volume | sd TimeSinceBL | 0.0520 |
| R CA1 volume | sd Observation | 0.2529 |
| R CA1 curvature | sd (Intercept) | 0.0302 |
| R CA1 curvature | cor (Intercept).TimeSinceBL | -0.4153 |
| R CA1 curvature | sd TimeSinceBL | 0.0012 |
| R CA1 curvature | sd Observation | 0.0088 |
| R CA1 gyrification | sd (Intercept) | 0.0298 |
| R CA1 gyrification | cor (Intercept).TimeSinceBL | -0.4208 |
| R CA1 gyrification | sd TimeSinceBL | 0.0012 |
| R CA1 gyrification | sd Observation | 0.0087 |
| R CA1 thickness | sd (Intercept) | 0.0301 |
| R CA1 thickness | cor (Intercept).TimeSinceBL | -0.4160 |
| R CA1 thickness | sd TimeSinceBL | 0.0012 |

| | | |
|---|---|---|
| R CA1 thickness | sd Observation | 0.0088 |
| R CA2 volume | sd (Intercept) | 0.1708 |
| R CA2 volume | cor (Intercept).TimeSinceBL | 0.0131 |
| R CA2 volume | sd TimeSinceBL | 0.0210 |
| R CA2 volume | sd Observation | 0.1844 |
| R CA2 curvature | sd (Intercept) | 0.0302 |
| R CA2 curvature | cor (Intercept).TimeSinceBL | -0.4150 |
| R CA2 curvature | sd TimeSinceBL | 0.0012 |
| R CA2 curvature | sd Observation | 0.0088 |
| R CA2 gyrification | sd (Intercept) | 0.0300 |
| R CA2 gyrification | cor (Intercept).TimeSinceBL | -0.4200 |
| R CA2 gyrification | sd TimeSinceBL | 0.0012 |
| R CA2 gyrification | sd Observation | 0.0089 |
| R CA2 thickness | sd (Intercept) | 0.0301 |
| R CA2 thickness | cor (Intercept).TimeSinceBL | -0.4184 |
| R CA2 thickness | sd TimeSinceBL | 0.0012 |
| R CA2 thickness | sd Observation | 0.0088 |
| R CA3 volume | sd (Intercept) | 0.2474 |
| R CA3 volume | cor (Intercept).TimeSinceBL | 0.3093 |
| R CA3 volume | sd TimeSinceBL | 0.0288 |
| R CA3 volume | sd Observation | 0.2015 |
| R CA3 curvature | sd (Intercept) | 0.0302 |
| R CA3 curvature | cor (Intercept).TimeSinceBL | -0.4142 |
| R CA3 curvature | sd TimeSinceBL | 0.0012 |
| R CA3 curvature | sd Observation | 0.0088 |

| | | |
|---|---|---|
| R CA3 gyrification | sd (Intercept) | 0.0299 |
| R CA3 gyrification | cor (Intercept).TimeSinceBL | -0.4141 |
| R CA3 gyrification | sd TimeSinceBL | 0.0012 |
| R CA3 gyrification | sd Observation | 0.0090 |
| R CA3 thickness | sd (Intercept) | 0.0301 |
| R CA3 thickness | cor (Intercept).TimeSinceBL | -0.4184 |
| R CA3 thickness | sd TimeSinceBL | 0.0012 |
| R CA3 thickness | sd Observation | 0.0088 |
| R CA4 volume | sd (Intercept) | 0.1978 |
| R CA4 volume | cor (Intercept).TimeSinceBL | 0.1309 |
| R CA4 volume | sd TimeSinceBL | 0.0222 |
| R CA4 volume | sd Observation | 0.1515 |
| R CA4 curvature | sd (Intercept) | 0.0304 |
| R CA4 curvature | cor (Intercept).TimeSinceBL | -0.4105 |
| R CA4 curvature | sd TimeSinceBL | 0.0012 |
| R CA4 curvature | sd Observation | 0.0101 |
| R CA4 gyrification | sd (Intercept) | 0.0290 |
| R CA4 gyrification | cor (Intercept).TimeSinceBL | -0.3749 |
| R CA4 gyrification | sd TimeSinceBL | 0.0013 |
| R CA4 gyrification | sd Observation | 0.0097 |
| R CA4 thickness | sd (Intercept) | 0.0301 |
| R CA4 thickness | cor (Intercept).TimeSinceBL | -0.4205 |
| R CA4 thickness | sd TimeSinceBL | 0.0012 |
| R CA4 thickness | sd Observation | 0.0088 |
| R DG volume | sd (Intercept) | 0.1338 |

| | | |
|---|---|---|
| R DG volume | cor (Intercept).TimeSinceBL | 0.0241 |
| R DG volume | sd TimeSinceBL | 0.0143 |
| R DG volume | sd Observation | 0.0495 |
| R DG curvature | sd (Intercept) | 0.0302 |
| R DG curvature | cor (Intercept).TimeSinceBL | -0.4196 |
| R DG curvature | sd TimeSinceBL | 0.0012 |
| R DG curvature | sd Observation | 0.0089 |
| R DG gyrification | sd (Intercept) | 0.0297 |
| R DG gyrification | cor (Intercept).TimeSinceBL | -0.4349 |
| R DG gyrification | sd TimeSinceBL | 0.0013 |
| R DG gyrification | sd Observation | 0.0093 |
| R Sub volume | sd (Intercept) | 0.3239 |
| R Sub volume | cor (Intercept).TimeSinceBL | -0.1517 |
| R Sub volume | sd TimeSinceBL | 0.0228 |
| R Sub volume | sd Observation | 0.1806 |
| R Sub curvature | sd (Intercept) | 0.0302 |
| R Sub curvature | cor (Intercept).TimeSinceBL | -0.4172 |
| R Sub curvature | sd TimeSinceBL | 0.0012 |
| R Sub curvature | sd Observation | 0.0089 |
| R Sub gyrification | sd (Intercept) | 0.0300 |
| R Sub gyrification | cor (Intercept).TimeSinceBL | -0.4215 |
| R Sub gyrification | sd TimeSinceBL | 0.0012 |
| R Sub gyrification | sd Observation | 0.0088 |
| R Sub thickness | sd (Intercept) | 0.0301 |
| R Sub thickness | cor (Intercept).TimeSinceBL | -0.4168 |

| | | |
|---|---|---|
| R Sub thickness | sd TimeSinceBL | 0.0012 |
| R Sub thickness | sd Observation | 0.0088 |
| R SRLM volume | sd (Intercept) | 0.4418 |
| R SRLM volume | cor (Intercept).TimeSinceBL | -0.0219 |
| R SRLM volume | sd TimeSinceBL | 0.0450 |
| R SRLM volume | sd Observation | 0.1417 |
| R Cyst volume | sd (Intercept) | 0.0950 |
| R Cyst volume | cor (Intercept).TimeSinceBL | -0.3865 |
| R Cyst volume | sd TimeSinceBL | 0.0047 |
| R Cyst volume | sd Observation | 0.0230 |

**Table S3**

*Interaction of Morphometry and Diagnosis on Cognitive Domain Filtered by Adjusted Significance*

| Model | Term | Estimate | Std.Error | Statistic | Adj p.value |
|---|---|---|---|---|---|
| L CA1 curvature PHC EXF | morphometry | -3.6882 | 0.51 | -7.25 | <0.01 |
| L CA1 curvature PHC EXF | morphometry:Stable AD | 3.8833 | 0.80 | 4.83 | <0.01 |
| L CA1 curvature PHC EXF | morphometry:CN to MCI/AD | -2.5039 | 1.08 | -2.31 | 0.04 |
| L CA1 curvature PHC LAN | morphometry | -3.7435 | 0.50 | -7.54 | <0.01 |
| L CA1 curvature PHC LAN | morphometry:CN to MCI/AD | -3.4259 | 1.06 | -3.24 | <0.01 |
| L CA1 curvature PHC MEM | morphometry:MCI to AD | -3.0408 | 0.60 | -5.03 | <0.01 |
| L CA1 curvature PHC MEM | morphometry:Stable MCI | -2.5701 | 0.58 | -4.44 | <0.01 |
| L CA1 curvature PHC MEM | morphometry:CN to MCI/AD | -3.7193 | 0.87 | -4.27 | <0.01 |
| L CA1 curvature PHC VSP | morphometry:Stable AD | 3.8744 | 1.22 | 3.18 | <0.01 |
| L CA1 gyrification PHC EXF | morphometry | -3.7687 | 0.52 | -7.31 | <0.01 |
| L CA1 gyrification PHC EXF | morphometry:Stable AD | 3.8894 | 0.81 | 4.77 | <0.01 |
| L CA1 gyrification PHC LAN | morphometry | -3.8219 | 0.50 | -7.58 | <0.01 |

| | | | | | |
|---|---|---|---|---|---|
| L CA1 gyrification PHC LAN | morphometry:CN to MCI/AD | -3.3647 | 1.07 | -3.13 | <0.01 |
| L CA1 gyrification PHC MEM | morphometry:MCI to AD | -2.8948 | 0.62 | -4.69 | <0.01 |
| L CA1 gyrification PHC MEM | morphometry:Stable MCI | -2.4087 | 0.59 | -4.08 | <0.01 |
| L CA1 gyrification PHC MEM | morphometry:CN to MCI/AD | -3.6169 | 0.89 | -4.08 | <0.01 |
| L CA1 gyrification PHC VSP | morphometry:Stable AD | 3.8421 | 1.24 | 3.11 | <0.01 |
| L CA1 thickness PHC EXF | morphometry | -3.7026 | 0.51 | -7.26 | <0.01 |
| L CA1 thickness PHC EXF | morphometry:Stable AD | 3.9049 | 0.81 | 4.85 | <0.01 |
| L CA1 thickness PHC EXF | morphometry:CN to MCI/AD | -2.5200 | 1.09 | -2.32 | 0.04 |
| L CA1 thickness PHC LAN | morphometry | -3.7578 | 0.50 | -7.54 | <0.01 |
| L CA1 thickness PHC LAN | morphometry:CN to MCI/AD | -3.4538 | 1.06 | -3.26 | <0.01 |
| L CA1 thickness PHC MEM | morphometry:MCI to AD | -3.0622 | 0.61 | -5.06 | <0.01 |
| L CA1 thickness PHC MEM | morphometry:Stable MCI | -2.5790 | 0.58 | -4.44 | <0.01 |
| L CA1 thickness PHC MEM | morphometry:CN to MCI/AD | -3.7490 | 0.87 | -4.29 | <0.01 |
| L CA1 thickness PHC VSP | morphometry:Stable AD | 3.8916 | 1.22 | 3.18 | <0.01 |
| L CA1 volume PHC EXF | morphometry:Stable MCI | 0.1737 | 0.05 | 3.62 | <0.01 |
| L CA1 volume PHC EXF | morphometry:CN to MCI/AD | 0.2084 | 0.07 | 3.08 | <0.01 |
| L CA1 volume PHC EXF | morphometry:Stable AD | -0.1408 | 0.06 | -2.49 | 0.02 |
| L CA1 volume PHC LAN | morphometry:Stable MCI | 0.2325 | 0.05 | 4.96 | <0.01 |
| L CA1 volume PHC LAN | morphometry | 0.1000 | 0.04 | 2.82 | 0.01 |
| L CA1 volume PHC LAN | morphometry:CN to MCI/AD | 0.1769 | 0.07 | 2.68 | 0.01 |
| L CA1 volume PHC MEM | morphometry:Stable MCI | 0.3188 | 0.04 | 8.64 | <0.01 |
| L CA1 volume PHC MEM | morphometry:MCI to AD | 0.2008 | 0.04 | 5.45 | <0.01 |
| L CA1 volume PHC MEM | morphometry:CN to MCI/AD | 0.2265 | 0.05 | 4.37 | <0.01 |
| L CA1 volume PHC MEM | morphometry | 0.1039 | 0.03 | 3.73 | <0.01 |
| L CA1 volume PHC MEM | morphometry:Stable AD | 0.1036 | 0.04 | 2.39 | 0.03 |

| | | | | | |
|---|---|---|---|---|---|
| L CA2 curvature PHC EXF | morphometry | -3.6891 | 0.51 | -7.25 | <0.01 |
| L CA2 curvature PHC EXF | morphometry:Stable AD | 3.8874 | 0.80 | 4.83 | <0.01 |
| L CA2 curvature PHC EXF | morphometry:CN to MCI/AD | -2.5045 | 1.08 | -2.32 | 0.04 |
| L CA2 curvature PHC LAN | morphometry | -3.7449 | 0.50 | -7.54 | <0.01 |
| L CA2 curvature PHC LAN | morphometry:CN to MCI/AD | -3.4359 | 1.06 | -3.25 | <0.01 |
| L CA2 curvature PHC MEM | morphometry:MCI to AD | -3.0158 | 0.60 | -4.99 | <0.01 |
| L CA2 curvature PHC MEM | morphometry:Stable MCI | -2.5546 | 0.58 | -4.41 | <0.01 |
| L CA2 curvature PHC MEM | morphometry:CN to MCI/AD | -3.7251 | 0.87 | -4.27 | <0.01 |
| L CA2 curvature PHC VSP | morphometry:Stable AD | 3.8781 | 1.22 | 3.18 | <0.01 |
| L CA2 gyrification PHC EXF | morphometry | -3.7182 | 0.51 | -7.31 | <0.01 |
| L CA2 gyrification PHC EXF | morphometry:Stable AD | 3.8340 | 0.80 | 4.77 | <0.01 |
| L CA2 gyrification PHC EXF | morphometry:CN to MCI/AD | -2.4652 | 1.08 | -2.28 | 0.04 |
| L CA2 gyrification PHC LAN | morphometry | -3.7727 | 0.50 | -7.60 | <0.01 |
| L CA2 gyrification PHC LAN | morphometry:CN to MCI/AD | -3.3840 | 1.06 | -3.20 | <0.01 |
| L CA2 gyrification PHC MEM | morphometry:MCI to AD | -3.1669 | 0.60 | -5.24 | <0.01 |
| L CA2 gyrification PHC MEM | morphometry:Stable MCI | -2.6194 | 0.58 | -4.52 | <0.01 |
| L CA2 gyrification PHC MEM | morphometry:CN to MCI/AD | -3.8191 | 0.87 | -4.38 | <0.01 |
| L CA2 gyrification PHC VSP | morphometry:Stable AD | 3.8094 | 1.22 | 3.12 | <0.01 |
| L CA2 thickness PHC EXF | morphometry | -3.7030 | 0.51 | -7.27 | <0.01 |
| L CA2 thickness PHC EXF | morphometry:Stable AD | 3.8923 | 0.80 | 4.84 | <0.01 |
| L CA2 thickness PHC EXF | morphometry:CN to MCI/AD | -2.5504 | 1.08 | -2.35 | 0.03 |
| L CA2 thickness PHC LAN | morphometry | -3.7630 | 0.50 | -7.57 | <0.01 |
| L CA2 thickness PHC LAN | morphometry:CN to MCI/AD | -3.4812 | 1.06 | -3.29 | <0.01 |
| L CA2 thickness PHC MEM | morphometry:MCI to AD | -3.0678 | 0.60 | -5.08 | <0.01 |
| L CA2 thickness PHC MEM | morphometry:Stable MCI | -2.6043 | 0.58 | -4.49 | <0.01 |

| | | | | | |
|---|---|---|---|---|---|
| L CA2 thickness PHC MEM | morphometry:CN to MCI/AD | -3.7823 | 0.87 | -4.33 | <0.01 |
| L CA2 thickness PHC VSP | morphometry:Stable AD | 3.8859 | 1.22 | 3.18 | <0.01 |
| L CA2 volume PHC EXF | morphometry | -0.4037 | 0.13 | -3.05 | 0.01 |
| L CA2 volume PHC LAN | morphometry | -0.3647 | 0.13 | -2.81 | 0.01 |
| L CA2 volume PHC LAN | morphometry:CN to MCI/AD | 0.4626 | 0.20 | 2.34 | 0.04 |
| L CA2 volume PHC MEM | morphometry:Stable MCI | -0.3831 | 0.13 | -2.92 | 0.01 |
| L CA3 curvature PHC EXF | morphometry | -3.6886 | 0.51 | -7.25 | <0.01 |
| L CA3 curvature PHC EXF | morphometry:Stable AD | 3.9003 | 0.80 | 4.85 | <0.01 |
| L CA3 curvature PHC EXF | morphometry:CN to MCI/AD | -2.5087 | 1.08 | -2.32 | 0.04 |
| L CA3 curvature PHC LAN | morphometry | -3.7422 | 0.50 | -7.53 | <0.01 |
| L CA3 curvature PHC LAN | morphometry:CN to MCI/AD | -3.4424 | 1.06 | -3.25 | <0.01 |
| L CA3 curvature PHC MEM | morphometry:MCI to AD | -3.0052 | 0.61 | -4.97 | <0.01 |
| L CA3 curvature PHC MEM | morphometry:Stable MCI | -2.5506 | 0.58 | -4.40 | <0.01 |
| L CA3 curvature PHC MEM | morphometry:CN to MCI/AD | -3.7235 | 0.87 | -4.27 | <0.01 |
| L CA3 curvature PHC VSP | morphometry:Stable AD | 3.8861 | 1.22 | 3.19 | <0.01 |
| L CA3 gyrification PHC EXF | morphometry | -3.7953 | 0.51 | -7.40 | <0.01 |
| L CA3 gyrification PHC EXF | morphometry:Stable AD | 3.8735 | 0.81 | 4.78 | <0.01 |
| L CA3 gyrification PHC EXF | morphometry:CN to MCI/AD | -2.4599 | 1.08 | -2.27 | 0.04 |
| L CA3 gyrification PHC LAN | morphometry | -3.8662 | 0.50 | -7.72 | <0.01 |
| L CA3 gyrification PHC LAN | morphometry:CN to MCI/AD | -3.4017 | 1.06 | -3.21 | <0.01 |
| L CA3 gyrification PHC MEM | morphometry:MCI to AD | -3.0639 | 0.61 | -5.02 | <0.01 |
| L CA3 gyrification PHC MEM | morphometry:CN to MCI/AD | -3.6846 | 0.87 | -4.21 | <0.01 |
| L CA3 gyrification PHC MEM | morphometry:Stable MCI | -2.3936 | 0.58 | -4.09 | <0.01 |
| L CA3 gyrification PHC MEM | morphometry | -0.9292 | 0.41 | -2.25 | 0.04 |
| L CA3 gyrification PHC VSP | morphometry:Stable AD | 3.7694 | 1.23 | 3.07 | <0.01 |

| | | | | | |
|---|---|---|---|---|---|
| L CA3 thickness PHC EXF | morphometry | -3.7021 | 0.51 | -7.26 | <0.01 |
| L CA3 thickness PHC EXF | morphometry:Stable AD | 3.9184 | 0.81 | 4.86 | <0.01 |
| L CA3 thickness PHC EXF | morphometry:CN to MCI/AD | -2.5082 | 1.09 | -2.31 | 0.04 |
| L CA3 thickness PHC LAN | morphometry | -3.7618 | 0.50 | -7.55 | <0.01 |
| L CA3 thickness PHC LAN | morphometry:CN to MCI/AD | -3.4525 | 1.06 | -3.26 | <0.01 |
| L CA3 thickness PHC MEM | morphometry:MCI to AD | -3.0046 | 0.61 | -4.96 | <0.01 |
| L CA3 thickness PHC MEM | morphometry:Stable MCI | -2.5564 | 0.58 | -4.40 | <0.01 |
| L CA3 thickness PHC MEM | morphometry:CN to MCI/AD | -3.7323 | 0.87 | -4.27 | <0.01 |
| L CA3 thickness PHC VSP | morphometry:Stable AD | 3.8914 | 1.22 | 3.18 | <0.01 |
| L CA3 volume PHC EXF | morphometry:MCI to AD | -0.2441 | 0.09 | -2.85 | 0.01 |
| L CA3 volume PHC LAN | morphometry:Stable MCI | 0.3528 | 0.09 | 3.73 | <0.01 |
| L CA3 volume PHC MEM | morphometry:Stable MCI | 0.5041 | 0.08 | 6.55 | <0.01 |
| L CA4 curvature PHC EXF | morphometry | -3.6205 | 0.51 | -7.16 | <0.01 |
| L CA4 curvature PHC EXF | morphometry:Stable AD | 3.8789 | 0.81 | 4.82 | <0.01 |
| L CA4 curvature PHC LAN | morphometry | -3.6605 | 0.49 | -7.40 | <0.01 |
| L CA4 curvature PHC LAN | morphometry:CN to MCI/AD | -3.0247 | 1.04 | -2.91 | 0.01 |
| L CA4 curvature PHC MEM | morphometry:MCI to AD | -2.9211 | 0.61 | -4.82 | <0.01 |
| L CA4 curvature PHC MEM | morphometry:Stable MCI | -2.3755 | 0.58 | -4.11 | <0.01 |
| L CA4 curvature PHC MEM | morphometry:CN to MCI/AD | -3.4577 | 0.86 | -4.03 | <0.01 |
| L CA4 curvature PHC VSP | morphometry:Stable AD | 3.9542 | 1.22 | 3.25 | <0.01 |
| L CA4 gyrification PHC EXF | morphometry | -3.9462 | 0.52 | -7.60 | <0.01 |
| L CA4 gyrification PHC EXF | morphometry:Stable AD | 3.8107 | 0.82 | 4.64 | <0.01 |
| L CA4 gyrification PHC LAN | morphometry | -4.0518 | 0.51 | -7.98 | <0.01 |
| L CA4 gyrification PHC LAN | morphometry:CN to MCI/AD | -3.5379 | 1.07 | -3.30 | <0.01 |
| L CA4 gyrification PHC MEM | morphometry:MCI to AD | -2.6776 | 0.62 | -4.30 | <0.01 |

| | | | | | |
|---|---|---|---|---|---|
| L CA4 gyrification PHC MEM | morphometry:CN to MCI/AD | -3.5233 | 0.89 | -3.97 | <0.01 |
| L CA4 gyrification PHC MEM | morphometry:Stable MCI | -1.9811 | 0.59 | -3.33 | <0.01 |
| L CA4 gyrification PHC MEM | morphometry | -1.0651 | 0.42 | -2.54 | 0.02 |
| L CA4 gyrification PHC VSP | morphometry:Stable AD | 3.5509 | 1.25 | 2.84 | 0.01 |
| L CA4 thickness PHC EXF | morphometry | -3.6989 | 0.51 | -7.24 | <0.01 |
| L CA4 thickness PHC EXF | morphometry:Stable AD | 3.9176 | 0.81 | 4.85 | <0.01 |
| L CA4 thickness PHC EXF | morphometry:CN to MCI/AD | -2.4793 | 1.09 | -2.28 | 0.04 |
| L CA4 thickness PHC LAN | morphometry | -3.7687 | 0.50 | -7.56 | <0.01 |
| L CA4 thickness PHC LAN | morphometry:CN to MCI/AD | -3.4273 | 1.06 | -3.23 | <0.01 |
| L CA4 thickness PHC MEM | morphometry:MCI to AD | -3.0154 | 0.61 | -4.97 | <0.01 |
| L CA4 thickness PHC MEM | morphometry:Stable MCI | -2.5031 | 0.58 | -4.30 | <0.01 |
| L CA4 thickness PHC MEM | morphometry:CN to MCI/AD | -3.6863 | 0.88 | -4.21 | <0.01 |
| L CA4 thickness PHC VSP | morphometry:Stable AD | 3.8789 | 1.22 | 3.17 | <0.01 |
| L CA4 volume PHC LAN | morphometry:Stable MCI | 0.6826 | 0.14 | 4.89 | <0.01 |
| L CA4 volume PHC LAN | morphometry:MCI to AD | 0.5454 | 0.14 | 4.01 | <0.01 |
| L CA4 volume PHC LAN | morphometry | -0.3261 | 0.11 | -3.05 | 0.01 |
| L CA4 volume PHC LAN | morphometry:Stable AD | 0.3766 | 0.15 | 2.53 | 0.02 |
| L CA4 volume PHC MEM | morphometry:Stable MCI | 0.8061 | 0.11 | 7.15 | <0.01 |
| L CA4 volume PHC MEM | morphometry:MCI to AD | 0.5876 | 0.11 | 5.35 | <0.01 |
| L CA4 volume PHC MEM | morphometry:Stable AD | 0.3723 | 0.12 | 3.09 | <0.01 |
| L CA4 volume PHC MEM | morphometry:CN to MCI/AD | 0.3645 | 0.16 | 2.31 | 0.04 |
| L Cyst volume PHC EXF | morphometry:MCI to AD | -1.0873 | 0.37 | -2.95 | 0.01 |
| L Cyst volume PHC EXF | morphometry | -0.5144 | 0.19 | -2.71 | 0.01 |
| L Cyst volume PHC LAN | morphometry | -0.6252 | 0.19 | -3.33 | <0.01 |
| L DG curvature PHC EXF | morphometry | -3.6887 | 0.51 | -7.28 | <0.01 |

| | | | | | |
|---|---|---|---|---|---|
| L DG curvature PHC EXF | morphometry:Stable AD | 3.8637 | 0.80 | 4.81 | <0.01 |
| L DG curvature PHC EXF | morphometry:CN to MCI/AD | -2.5379 | 1.08 | -2.35 | 0.03 |
| L DG curvature PHC LAN | morphometry | -3.7439 | 0.49 | -7.57 | <0.01 |
| L DG curvature PHC LAN | morphometry:CN to MCI/AD | -3.4984 | 1.05 | -3.32 | <0.01 |
| L DG curvature PHC MEM | morphometry:MCI to AD | -2.8578 | 0.59 | -4.84 | <0.01 |
| L DG curvature PHC MEM | morphometry:Stable MCI | -2.6541 | 0.58 | -4.60 | <0.01 |
| L DG curvature PHC MEM | morphometry:CN to MCI/AD | -3.7695 | 0.87 | -4.34 | <0.01 |
| L DG curvature PHC VSP | morphometry:Stable AD | 3.8348 | 1.22 | 3.14 | <0.01 |
| L DG gyrification PHC EXF | morphometry | -3.8453 | 0.52 | -7.38 | <0.01 |
| L DG gyrification PHC EXF | morphometry:Stable AD | 3.9726 | 0.83 | 4.78 | <0.01 |
| L DG gyrification PHC LAN | morphometry | -3.9461 | 0.51 | -7.74 | <0.01 |
| L DG gyrification PHC LAN | morphometry:CN to MCI/AD | -3.0291 | 1.08 | -2.79 | 0.01 |
| L DG gyrification PHC MEM | morphometry:MCI to AD | -2.5333 | 0.63 | -4.01 | <0.01 |
| L DG gyrification PHC MEM | morphometry:CN to MCI/AD | -3.1022 | 0.90 | -3.46 | <0.01 |
| L DG gyrification PHC MEM | morphometry:Stable MCI | -1.9259 | 0.60 | -3.21 | <0.01 |
| L DG gyrification PHC VSP | morphometry:Stable AD | 3.8765 | 1.26 | 3.08 | <0.01 |
| L DG volume PHC EXF | morphometry:Stable MCI | 1.0922 | 0.24 | 4.52 | <0.01 |
| L DG volume PHC LAN | morphometry:Stable MCI | 1.4322 | 0.24 | 6.09 | <0.01 |
| L DG volume PHC LAN | morphometry:MCI to AD | 0.8138 | 0.24 | 3.43 | <0.01 |
| L DG volume PHC LAN | morphometry:Stable AD | 0.8382 | 0.28 | 3.00 | 0.01 |
| L DG volume PHC MEM | morphometry:Stable MCI | 1.3093 | 0.18 | 7.18 | <0.01 |
| L DG volume PHC MEM | morphometry | 0.8746 | 0.15 | 5.95 | <0.01 |
| L DG volume PHC MEM | morphometry:MCI to AD | 0.5430 | 0.18 | 2.95 | 0.01 |
| L DG volume PHC MEM | morphometry:CN to MCI/AD | 0.7413 | 0.26 | 2.91 | 0.01 |
| L SRLM volume PHC EXF | morphometry:Stable MCI | 0.2368 | 0.07 | 3.21 | <0.01 |

| | | | | | |
|---|---|---|---|---|---|
| L SRLM volume PHC LAN | morphometry:Stable MCI | 0.3970 | 0.07 | 5.53 | <0.01 |
| L SRLM volume PHC LAN | morphometry:MCI to AD | 0.1763 | 0.07 | 2.45 | 0.03 |
| L SRLM volume PHC MEM | morphometry:Stable MCI | 0.4966 | 0.06 | 8.92 | <0.01 |
| L SRLM volume PHC MEM | morphometry:MCI to AD | 0.3075 | 0.06 | 5.51 | <0.01 |
| L SRLM volume PHC MEM | morphometry | 0.1762 | 0.04 | 3.93 | <0.01 |
| L SRLM volume PHC MEM | morphometry:CN to MCI/AD | 0.3071 | 0.08 | 3.91 | <0.01 |
| L Sub curvature PHC EXF | morphometry | -3.6885 | 0.51 | -7.24 | <0.01 |
| L Sub curvature PHC EXF | morphometry:Stable AD | 3.8902 | 0.81 | 4.83 | <0.01 |
| L Sub curvature PHC EXF | morphometry:MCI to AD | 1.5563 | 0.64 | 2.45 | 0.03 |
| L Sub curvature PHC EXF | morphometry:CN to MCI/AD | -2.5217 | 1.08 | -2.33 | 0.04 |
| L Sub curvature PHC LAN | morphometry | -3.7420 | 0.50 | -7.52 | <0.01 |
| L Sub curvature PHC LAN | morphometry:CN to MCI/AD | -3.4565 | 1.06 | -3.26 | <0.01 |
| L Sub curvature PHC MEM | morphometry:Stable MCI | -2.5666 | 0.58 | -4.41 | <0.01 |
| L Sub curvature PHC MEM | morphometry:CN to MCI/AD | -3.7098 | 0.88 | -4.24 | <0.01 |
| L Sub curvature PHC MEM | morphometry:MCI to AD | -1.2755 | 0.51 | -2.49 | 0.02 |
| L Sub curvature PHC VSP | morphometry:Stable AD | 3.8827 | 1.22 | 3.18 | <0.01 |
| L Sub gyrification PHC EXF | morphometry | -3.7039 | 0.51 | -7.21 | <0.01 |
| L Sub gyrification PHC EXF | morphometry:Stable AD | 3.9480 | 0.81 | 4.86 | <0.01 |
| L Sub gyrification PHC EXF | morphometry:CN to MCI/AD | -2.4773 | 1.09 | -2.27 | 0.04 |
| L Sub gyrification PHC LAN | morphometry | -3.7680 | 0.50 | -7.51 | <0.01 |
| L Sub gyrification PHC LAN | morphometry:CN to MCI/AD | -3.4322 | 1.07 | -3.22 | <0.01 |
| L Sub gyrification PHC MEM | morphometry:MCI to AD | -2.9341 | 0.61 | -4.80 | <0.01 |
| L Sub gyrification PHC MEM | morphometry:Stable MCI | -2.4450 | 0.59 | -4.16 | <0.01 |
| L Sub gyrification PHC MEM | morphometry:CN to MCI/AD | -3.6163 | 0.88 | -4.11 | <0.01 |
| L Sub gyrification PHC VSP | morphometry:Stable AD | 3.9416 | 1.23 | 3.20 | <0.01 |

| | | | | | |
|---|---|---|---|---|---|
| L Sub thickness PHC EXF | morphometry | -3.7004 | 0.51 | -7.26 | <0.01 |
| L Sub thickness PHC EXF | morphometry:Stable AD | 3.8899 | 0.81 | 4.82 | <0.01 |
| L Sub thickness PHC EXF | morphometry:CN to MCI/AD | -2.5224 | 1.08 | -2.33 | 0.04 |
| L Sub thickness PHC LAN | morphometry | -3.7672 | 0.50 | -7.57 | <0.01 |
| L Sub thickness PHC LAN | morphometry:CN to MCI/AD | -3.4551 | 1.06 | -3.26 | <0.01 |
| L Sub thickness PHC MEM | morphometry:MCI to AD | -3.0472 | 0.61 | -5.03 | <0.01 |
| L Sub thickness PHC MEM | morphometry:Stable MCI | -2.6368 | 0.58 | -4.54 | <0.01 |
| L Sub thickness PHC MEM | morphometry:CN to MCI/AD | -3.7500 | 0.87 | -4.29 | <0.01 |
| L Sub thickness PHC VSP | morphometry:Stable AD | 3.9197 | 1.22 | 3.20 | <0.01 |
| L Sub volume PHC EXF | morphometry:Stable MCI | 0.2490 | 0.09 | 2.77 | 0.01 |
| L Sub volume PHC EXF | morphometry:MCI to AD | -0.2488 | 0.09 | -2.76 | 0.01 |
| L Sub volume PHC LAN | morphometry:Stable MCI | 0.5116 | 0.09 | 5.80 | <0.01 |
| L Sub volume PHC MEM | morphometry:Stable MCI | 0.6083 | 0.07 | 8.59 | <0.01 |
| L Sub volume PHC MEM | morphometry:CN to MCI/AD | 0.2872 | 0.10 | 2.92 | 0.01 |
| L Sub volume PHC MEM | morphometry:Stable AD | 0.2200 | 0.08 | 2.75 | 0.01 |
| L Sub volume PHC MEM | morphometry:MCI to AD | 0.1761 | 0.07 | 2.48 | 0.03 |
| R CA1 curvature PHC EXF | morphometry | -3.6840 | 0.51 | -7.25 | <0.01 |
| R CA1 curvature PHC EXF | morphometry:Stable AD | 3.8877 | 0.80 | 4.84 | <0.01 |
| R CA1 curvature PHC EXF | morphometry:CN to MCI/AD | -2.5152 | 1.08 | -2.33 | 0.04 |
| R CA1 curvature PHC LAN | morphometry | -3.7399 | 0.50 | -7.54 | <0.01 |
| R CA1 curvature PHC LAN | morphometry:CN to MCI/AD | -3.4451 | 1.06 | -3.26 | <0.01 |
| R CA1 curvature PHC MEM | morphometry:MCI to AD | -3.0225 | 0.60 | -5.01 | <0.01 |
| R CA1 curvature PHC MEM | morphometry:Stable MCI | -2.5703 | 0.58 | -4.44 | <0.01 |
| R CA1 curvature PHC MEM | morphometry:CN to MCI/AD | -3.7385 | 0.87 | -4.29 | <0.01 |
| R CA1 curvature PHC VSP | morphometry:Stable AD | 3.8821 | 1.22 | 3.19 | <0.01 |

| | | | | | |
|---|---|---|---|---|---|
| R CA1 gyrification PHC EXF | morphometry | -3.7788 | 0.52 | -7.31 | <0.01 |
| R CA1 gyrification PHC EXF | morphometry:Stable AD | 4.0023 | 0.82 | 4.90 | <0.01 |
| R CA1 gyrification PHC LAN | morphometry | -3.8314 | 0.50 | -7.59 | <0.01 |
| R CA1 gyrification PHC LAN | morphometry:CN to MCI/AD | -3.3884 | 1.08 | -3.15 | <0.01 |
| R CA1 gyrification PHC MEM | morphometry:MCI to AD | -2.9489 | 0.62 | -4.78 | <0.01 |
| R CA1 gyrification PHC MEM | morphometry:Stable MCI | -2.4326 | 0.59 | -4.12 | <0.01 |
| R CA1 gyrification PHC MEM | morphometry:CN to MCI/AD | -3.5715 | 0.89 | -4.02 | <0.01 |
| R CA1 gyrification PHC VSP | morphometry:Stable AD | 3.9767 | 1.24 | 3.21 | <0.01 |
| R CA1 thickness PHC EXF | morphometry | -3.6992 | 0.51 | -7.25 | <0.01 |
| R CA1 thickness PHC EXF | morphometry:Stable AD | 3.8935 | 0.81 | 4.83 | <0.01 |
| R CA1 thickness PHC EXF | morphometry:CN to MCI/AD | -2.5215 | 1.08 | -2.33 | 0.04 |
| R CA1 thickness PHC LAN | morphometry | -3.7599 | 0.50 | -7.55 | <0.01 |
| R CA1 thickness PHC LAN | morphometry:CN to MCI/AD | -3.4635 | 1.06 | -3.27 | <0.01 |
| R CA1 thickness PHC MEM | morphometry:MCI to AD | -3.0394 | 0.61 | -5.02 | <0.01 |
| R CA1 thickness PHC MEM | morphometry:Stable MCI | -2.5795 | 0.58 | -4.44 | <0.01 |
| R CA1 thickness PHC MEM | morphometry:CN to MCI/AD | -3.7590 | 0.87 | -4.30 | <0.01 |
| R CA1 thickness PHC VSP | morphometry:Stable AD | 3.8643 | 1.22 | 3.16 | <0.01 |
| R CA1 volume PHC EXF | morphometry:CN to MCI/AD | 0.1632 | 0.06 | 2.63 | 0.02 |
| R CA1 volume PHC EXF | morphometry:Stable MCI | 0.1163 | 0.05 | 2.49 | 0.02 |
| R CA1 volume PHC EXF | morphometry:MCI to AD | -0.1103 | 0.05 | -2.31 | 0.04 |
| R CA1 volume PHC EXF | morphometry | 0.0802 | 0.04 | 2.24 | 0.04 |
| R CA1 volume PHC LAN | morphometry:Stable MCI | 0.1692 | 0.05 | 3.69 | <0.01 |
| R CA1 volume PHC LAN | morphometry | 0.0829 | 0.04 | 2.36 | 0.03 |
| R CA1 volume PHC LAN | morphometry:CN to MCI/AD | 0.1433 | 0.06 | 2.36 | 0.03 |
| R CA1 volume PHC MEM | morphometry:Stable MCI | 0.3013 | 0.04 | 8.30 | <0.01 |

| | | | | | |
|---|---|---|---|---|---|
| R CA1 volume PHC MEM | morphometry:CN to MCI/AD | 0.2872 | 0.05 | 5.98 | <0.01 |
| R CA1 volume PHC MEM | morphometry:MCI to AD | 0.1926 | 0.04 | 5.21 | <0.01 |
| R CA1 volume PHC MEM | morphometry:Stable AD | 0.1776 | 0.04 | 4.26 | <0.01 |
| R CA1 volume PHC VSP | morphometry:CN to MCI/AD | 0.2906 | 0.11 | 2.72 | 0.01 |
| R CA2 curvature PHC EXF | morphometry | -3.6845 | 0.51 | -7.25 | <0.01 |
| R CA2 curvature PHC EXF | morphometry:Stable AD | 3.8788 | 0.80 | 4.83 | <0.01 |
| R CA2 curvature PHC EXF | morphometry:CN to MCI/AD | -2.5227 | 1.08 | -2.34 | 0.04 |
| R CA2 curvature PHC LAN | morphometry | -3.7398 | 0.50 | -7.54 | <0.01 |
| R CA2 curvature PHC LAN | morphometry:CN to MCI/AD | -3.4517 | 1.06 | -3.27 | <0.01 |
| R CA2 curvature PHC MEM | morphometry:MCI to AD | -3.0331 | 0.60 | -5.03 | <0.01 |
| R CA2 curvature PHC MEM | morphometry:Stable MCI | -2.5845 | 0.58 | -4.47 | <0.01 |
| R CA2 curvature PHC MEM | morphometry:CN to MCI/AD | -3.7527 | 0.87 | -4.31 | <0.01 |
| R CA2 curvature PHC VSP | morphometry:Stable AD | 3.8685 | 1.22 | 3.18 | <0.01 |
| R CA2 gyrification PHC EXF | morphometry | -3.7371 | 0.51 | -7.32 | <0.01 |
| R CA2 gyrification PHC EXF | morphometry:Stable AD | 3.8778 | 0.80 | 4.82 | <0.01 |
| R CA2 gyrification PHC EXF | morphometry:CN to MCI/AD | -2.5595 | 1.08 | -2.36 | 0.03 |
| R CA2 gyrification PHC LAN | morphometry | -3.8007 | 0.50 | -7.63 | <0.01 |
| R CA2 gyrification PHC LAN | morphometry:CN to MCI/AD | -3.5098 | 1.06 | -3.32 | <0.01 |
| R CA2 gyrification PHC MEM | morphometry:MCI to AD | -3.0653 | 0.60 | -5.08 | <0.01 |
| R CA2 gyrification PHC MEM | morphometry:Stable MCI | -2.5675 | 0.58 | -4.42 | <0.01 |
| R CA2 gyrification PHC MEM | morphometry:CN to MCI/AD | -3.8514 | 0.87 | -4.41 | <0.01 |
| R CA2 gyrification PHC VSP | morphometry:Stable AD | 3.8079 | 1.22 | 3.12 | <0.01 |
| R CA2 thickness PHC EXF | morphometry | -3.7006 | 0.51 | -7.27 | <0.01 |
| R CA2 thickness PHC EXF | morphometry:Stable AD | 3.8586 | 0.80 | 4.80 | <0.01 |
| R CA2 thickness PHC EXF | morphometry:CN to MCI/AD | -2.5613 | 1.08 | -2.36 | 0.03 |

| | | | | | |
|---|---|---|---|---|---|
| R CA2 thickness PHC LAN | morphometry | -3.7615 | 0.50 | -7.57 | <0.01 |
| R CA2 thickness PHC LAN | morphometry:CN to MCI/AD | -3.4920 | 1.06 | -3.30 | <0.01 |
| R CA2 thickness PHC MEM | morphometry:MCI to AD | -3.0510 | 0.60 | -5.05 | <0.01 |
| R CA2 thickness PHC MEM | morphometry:Stable MCI | -2.5939 | 0.58 | -4.48 | <0.01 |
| R CA2 thickness PHC MEM | morphometry:CN to MCI/AD | -3.8046 | 0.87 | -4.36 | <0.01 |
| R CA2 thickness PHC VSP | morphometry:Stable AD | 3.8309 | 1.22 | 3.14 | <0.01 |
| R CA2 volume PHC EXF | morphometry | -0.3891 | 0.15 | -2.68 | 0.01 |
| R CA2 volume PHC LAN | morphometry | -0.4347 | 0.14 | -3.04 | 0.01 |
| R CA2 volume PHC MEM | morphometry | -0.3214 | 0.12 | -2.74 | 0.01 |
| R CA3 curvature PHC EXF | morphometry | -3.6883 | 0.51 | -7.26 | <0.01 |
| R CA3 curvature PHC EXF | morphometry:Stable AD | 3.8720 | 0.80 | 4.83 | <0.01 |
| R CA3 curvature PHC EXF | morphometry:CN to MCI/AD | -2.5306 | 1.08 | -2.34 | 0.04 |
| R CA3 curvature PHC LAN | morphometry | -3.7437 | 0.50 | -7.55 | <0.01 |
| R CA3 curvature PHC LAN | morphometry:CN to MCI/AD | -3.4527 | 1.06 | -3.27 | <0.01 |
| R CA3 curvature PHC MEM | morphometry:MCI to AD | -3.0455 | 0.60 | -5.05 | <0.01 |
| R CA3 curvature PHC MEM | morphometry:Stable MCI | -2.5855 | 0.58 | -4.47 | <0.01 |
| R CA3 curvature PHC MEM | morphometry:CN to MCI/AD | -3.7550 | 0.87 | -4.32 | <0.01 |
| R CA3 curvature PHC VSP | morphometry:Stable AD | 3.8612 | 1.22 | 3.17 | <0.01 |
| R CA3 gyrification PHC EXF | morphometry | -3.7758 | 0.51 | -7.35 | <0.01 |
| R CA3 gyrification PHC EXF | morphometry:Stable AD | 3.8691 | 0.81 | 4.79 | <0.01 |
| R CA3 gyrification PHC EXF | morphometry:CN to MCI/AD | -2.5511 | 1.09 | -2.34 | 0.04 |
| R CA3 gyrification PHC LAN | morphometry | -3.8752 | 0.50 | -7.73 | <0.01 |
| R CA3 gyrification PHC LAN | morphometry:CN to MCI/AD | -3.4867 | 1.06 | -3.28 | <0.01 |
| R CA3 gyrification PHC MEM | morphometry:MCI to AD | -2.9740 | 0.61 | -4.88 | <0.01 |
| R CA3 gyrification PHC MEM | morphometry:CN to MCI/AD | -3.7422 | 0.88 | -4.26 | <0.01 |

| | | | | | |
|---|---|---|---|---|---|
| R CA3 gyrification PHC MEM | morphometry:Stable MCI | -2.4415 | 0.59 | -4.17 | <0.01 |
| R CA3 gyrification PHC VSP | morphometry:Stable AD | 3.7486 | 1.23 | 3.06 | 0.01 |
| R CA3 thickness PHC EXF | morphometry | -3.6938 | 0.51 | -7.25 | <0.01 |
| R CA3 thickness PHC EXF | morphometry:Stable AD | 3.8983 | 0.81 | 4.84 | <0.01 |
| R CA3 thickness PHC EXF | morphometry:CN to MCI/AD | -2.5254 | 1.08 | -2.33 | 0.04 |
| R CA3 thickness PHC LAN | morphometry | -3.7537 | 0.50 | -7.55 | <0.01 |
| R CA3 thickness PHC LAN | morphometry:CN to MCI/AD | -3.4791 | 1.06 | -3.28 | <0.01 |
| R CA3 thickness PHC MEM | morphometry:MCI to AD | -3.0095 | 0.61 | -4.97 | <0.01 |
| R CA3 thickness PHC MEM | morphometry:Stable MCI | -2.5580 | 0.58 | -4.40 | <0.01 |
| R CA3 thickness PHC MEM | morphometry:CN to MCI/AD | -3.7520 | 0.87 | -4.29 | <0.01 |
| R CA3 thickness PHC VSP | morphometry:Stable AD | 3.8815 | 1.22 | 3.18 | <0.01 |
| R CA3 volume PHC EXF | morphometry:MCI to AD | -0.3162 | 0.11 | -2.94 | 0.01 |
| R CA3 volume PHC EXF | morphometry:CN to MCI/AD | -0.4323 | 0.16 | -2.79 | 0.01 |
| R CA3 volume PHC LAN | morphometry:Stable MCI | 0.5938 | 0.11 | 5.20 | <0.01 |
| R CA3 volume PHC LAN | morphometry:CN to MCI/AD | -0.3811 | 0.15 | -2.50 | 0.02 |
| R CA3 volume PHC MEM | morphometry:Stable MCI | 0.7157 | 0.09 | 7.74 | <0.01 |
| R CA3 volume PHC VSP | morphometry:MCI to AD | -0.4842 | 0.18 | -2.65 | 0.02 |
| R CA3 volume PHC VSP | morphometry:CN to MCI/AD | -0.6841 | 0.26 | -2.62 | 0.02 |
| R CA4 curvature PHC EXF | morphometry | -3.6601 | 0.51 | -7.23 | <0.01 |
| R CA4 curvature PHC EXF | morphometry:Stable AD | 3.6327 | 0.79 | 4.57 | <0.01 |
| R CA4 curvature PHC EXF | morphometry:CN to MCI/AD | -2.5702 | 1.07 | -2.40 | 0.03 |
| R CA4 curvature PHC LAN | morphometry | -3.6932 | 0.49 | -7.48 | <0.01 |
| R CA4 curvature PHC LAN | morphometry:CN to MCI/AD | -3.5183 | 1.05 | -3.37 | <0.01 |
| R CA4 curvature PHC MEM | morphometry:MCI to AD | -3.0823 | 0.60 | -5.17 | <0.01 |
| R CA4 curvature PHC MEM | morphometry:Stable MCI | -2.6719 | 0.57 | -4.66 | <0.01 |

| | | | | | |
|---|---|---|---|---|---|
| R CA4 curvature PHC MEM | morphometry:CN to MCI/AD | -3.8172 | 0.86 | -4.43 | <0.01 |
| R CA4 curvature PHC VSP | morphometry:Stable AD | 3.6049 | 1.21 | 2.98 | 0.01 |
| R CA4 gyrification PHC EXF | morphometry | -4.0629 | 0.52 | -7.79 | <0.01 |
| R CA4 gyrification PHC EXF | morphometry:Stable AD | 3.8758 | 0.82 | 4.73 | <0.01 |
| R CA4 gyrification PHC LAN | morphometry | -4.1766 | 0.51 | -8.21 | <0.01 |
| R CA4 gyrification PHC LAN | morphometry:CN to MCI/AD | -3.4406 | 1.09 | -3.16 | <0.01 |
| R CA4 gyrification PHC MEM | morphometry:MCI to AD | -2.7588 | 0.62 | -4.44 | <0.01 |
| R CA4 gyrification PHC MEM | morphometry:Stable MCI | -2.1478 | 0.60 | -3.59 | <0.01 |
| R CA4 gyrification PHC MEM | morphometry:CN to MCI/AD | -3.1556 | 0.90 | -3.50 | <0.01 |
| R CA4 gyrification PHC MEM | morphometry | -1.0907 | 0.42 | -2.59 | 0.02 |
| R CA4 gyrification PHC VSP | morphometry:Stable AD | 3.4652 | 1.24 | 2.79 | 0.01 |
| R CA4 thickness PHC EXF | morphometry | -3.7042 | 0.51 | -7.26 | <0.01 |
| R CA4 thickness PHC EXF | morphometry:Stable AD | 3.9330 | 0.81 | 4.87 | <0.01 |
| R CA4 thickness PHC EXF | morphometry:CN to MCI/AD | -2.5262 | 1.09 | -2.32 | 0.04 |
| R CA4 thickness PHC LAN | morphometry | -3.7663 | 0.50 | -7.56 | <0.01 |
| R CA4 thickness PHC LAN | morphometry:CN to MCI/AD | -3.4596 | 1.06 | -3.25 | <0.01 |
| R CA4 thickness PHC MEM | morphometry:MCI to AD | -2.9811 | 0.61 | -4.91 | <0.01 |
| R CA4 thickness PHC MEM | morphometry:Stable MCI | -2.5075 | 0.58 | -4.31 | <0.01 |
| R CA4 thickness PHC MEM | morphometry:CN to MCI/AD | -3.7394 | 0.88 | -4.26 | <0.01 |
| R CA4 thickness PHC VSP | morphometry:Stable AD | 3.9039 | 1.22 | 3.19 | <0.01 |
| R CA4 volume PHC EXF | morphometry:Stable MCI | 0.6031 | 0.14 | 4.29 | <0.01 |
| R CA4 volume PHC EXF | morphometry | -0.2978 | 0.10 | -2.99 | 0.01 |
| R CA4 volume PHC LAN | morphometry:Stable MCI | 0.7421 | 0.14 | 5.33 | <0.01 |
| R CA4 volume PHC LAN | morphometry | -0.3972 | 0.10 | -4.03 | <0.01 |
| R CA4 volume PHC MEM | morphometry:Stable MCI | 0.9975 | 0.11 | 8.89 | <0.01 |

| | | | | | |
|---|---|---|---|---|---|
| R CA4 volume PHC MEM | morphometry:CN to MCI/AD | 0.7382 | 0.16 | 4.70 | <0.01 |
| R CA4 volume PHC MEM | morphometry:MCI to AD | 0.4116 | 0.10 | 3.95 | <0.01 |
| R Cyst volume PHC EXF | morphometry | -0.7162 | 0.20 | -3.59 | <0.01 |
| R Cyst volume PHC EXF | morphometry:Stable AD | 1.0112 | 0.42 | 2.41 | 0.03 |
| R Cyst volume PHC LAN | morphometry | -0.5974 | 0.20 | -3.03 | 0.01 |
| R Cyst volume PHC LAN | morphometry:CN to MCI/AD | -1.0988 | 0.40 | -2.75 | 0.01 |
| R Cyst volume PHC MEM | morphometry:CN to MCI/AD | -0.7633 | 0.32 | -2.35 | 0.03 |
| R Cyst volume PHC VSP | morphometry:Stable AD | 1.8805 | 0.60 | 3.13 | <0.01 |
| R DG curvature PHC EXF | morphometry | -3.6868 | 0.51 | -7.25 | <0.01 |
| R DG curvature PHC EXF | morphometry:Stable AD | 3.9068 | 0.80 | 4.87 | <0.01 |
| R DG curvature PHC EXF | morphometry:CN to MCI/AD | -2.5082 | 1.08 | -2.31 | 0.04 |
| R DG curvature PHC LAN | morphometry | -3.7366 | 0.50 | -7.52 | <0.01 |
| R DG curvature PHC LAN | morphometry:CN to MCI/AD | -3.4116 | 1.06 | -3.22 | <0.01 |
| R DG curvature PHC MEM | morphometry:MCI to AD | -2.9914 | 0.60 | -4.95 | <0.01 |
| R DG curvature PHC MEM | morphometry:Stable MCI | -2.5129 | 0.58 | -4.33 | <0.01 |
| R DG curvature PHC MEM | morphometry:CN to MCI/AD | -3.7040 | 0.87 | -4.24 | <0.01 |
| R DG curvature PHC VSP | morphometry:Stable AD | 3.8929 | 1.22 | 3.20 | <0.01 |
| R DG gyrification PHC EXF | morphometry | -3.8899 | 0.52 | -7.43 | <0.01 |
| R DG gyrification PHC EXF | morphometry:Stable AD | 4.1519 | 0.83 | 5.02 | <0.01 |
| R DG gyrification PHC LAN | morphometry | -3.9889 | 0.51 | -7.79 | <0.01 |
| R DG gyrification PHC LAN | morphometry:CN to MCI/AD | -3.3747 | 1.09 | -3.09 | <0.01 |
| R DG gyrification PHC MEM | morphometry:MCI to AD | -2.6685 | 0.63 | -4.23 | <0.01 |
| R DG gyrification PHC MEM | morphometry:CN to MCI/AD | -3.3748 | 0.90 | -3.73 | <0.01 |
| R DG gyrification PHC MEM | morphometry:Stable MCI | -2.0259 | 0.60 | -3.36 | <0.01 |
| R DG gyrification PHC VSP | morphometry:Stable AD | 4.0258 | 1.25 | 3.21 | <0.01 |

| | | | | | |
|---|---|---|---|---|---|
| R DG volume PHC EXF | morphometry:Stable MCI | 0.8774 | 0.23 | 3.74 | <0.01 |
| R DG volume PHC LAN | morphometry:Stable MCI | 1.1937 | 0.23 | 5.18 | <0.01 |
| R DG volume PHC LAN | morphometry:MCI to AD | 0.7736 | 0.24 | 3.22 | <0.01 |
| R DG volume PHC MEM | morphometry:Stable MCI | 1.1928 | 0.18 | 6.64 | <0.01 |
| R DG volume PHC MEM | morphometry:MCI to AD | 0.9711 | 0.19 | 5.18 | <0.01 |
| R DG volume PHC MEM | morphometry | 0.6482 | 0.14 | 4.47 | <0.01 |
| R DG volume PHC MEM | morphometry:CN to MCI/AD | 0.7048 | 0.24 | 2.93 | 0.01 |
| R SRLM volume PHC EXF | morphometry | 0.1879 | 0.05 | 3.44 | <0.01 |
| R SRLM volume PHC EXF | morphometry:MCI to AD | -0.1874 | 0.07 | -2.64 | 0.02 |
| R SRLM volume PHC EXF | morphometry:Stable MCI | 0.1552 | 0.07 | 2.28 | 0.04 |
| R SRLM volume PHC LAN | morphometry:Stable MCI | 0.2824 | 0.07 | 4.22 | <0.01 |
| R SRLM volume PHC LAN | morphometry | 0.1361 | 0.05 | 2.54 | 0.02 |
| R SRLM volume PHC MEM | morphometry:Stable MCI | 0.4332 | 0.05 | 8.32 | <0.01 |
| R SRLM volume PHC MEM | morphometry:MCI to AD | 0.3398 | 0.05 | 6.28 | <0.01 |
| R SRLM volume PHC MEM | morphometry:CN to MCI/AD | 0.3840 | 0.07 | 5.33 | <0.01 |
| R SRLM volume PHC MEM | morphometry:Stable AD | 0.2225 | 0.06 | 3.57 | <0.01 |
| R SRLM volume PHC MEM | morphometry | 0.1250 | 0.04 | 2.99 | 0.01 |
| R Sub curvature PHC EXF | morphometry | -3.6847 | 0.51 | -7.25 | <0.01 |
| R Sub curvature PHC EXF | morphometry:Stable AD | 3.8958 | 0.80 | 4.85 | <0.01 |
| R Sub curvature PHC EXF | morphometry:CN to MCI/AD | -2.5286 | 1.08 | -2.34 | 0.04 |
| R Sub curvature PHC LAN | morphometry | -3.7441 | 0.50 | -7.55 | <0.01 |
| R Sub curvature PHC LAN | morphometry:CN to MCI/AD | -3.4482 | 1.06 | -3.27 | <0.01 |
| R Sub curvature PHC MEM | morphometry:MCI to AD | -3.0555 | 0.60 | -5.06 | <0.01 |
| R Sub curvature PHC MEM | morphometry:Stable MCI | -2.5721 | 0.58 | -4.44 | <0.01 |
| R Sub curvature PHC MEM | morphometry:CN to MCI/AD | -3.7511 | 0.87 | -4.31 | <0.01 |

| | | | | | |
|---|---|---|---|---|---|
| R Sub curvature PHC VSP | morphometry:Stable AD | 3.8872 | 1.22 | 3.19 | <0.01 |
| R Sub gyrification PHC EXF | morphometry | -3.7186 | 0.51 | -7.25 | <0.01 |
| R Sub gyrification PHC EXF | morphometry:Stable AD | 3.9807 | 0.81 | 4.90 | <0.01 |
| R Sub gyrification PHC EXF | morphometry:CN to MCI/AD | -2.4716 | 1.09 | -2.27 | 0.04 |
| R Sub gyrification PHC LAN | morphometry | -3.7852 | 0.50 | -7.55 | <0.01 |
| R Sub gyrification PHC LAN | morphometry:CN to MCI/AD | -3.3885 | 1.07 | -3.18 | <0.01 |
| R Sub gyrification PHC MEM | morphometry:MCI to AD | -2.9441 | 0.61 | -4.81 | <0.01 |
| R Sub gyrification PHC MEM | morphometry:Stable MCI | -2.4591 | 0.59 | -4.20 | <0.01 |
| R Sub gyrification PHC MEM | morphometry:CN to MCI/AD | -3.5413 | 0.88 | -4.03 | <0.01 |
| R Sub gyrification PHC VSP | morphometry:Stable AD | 3.9907 | 1.23 | 3.24 | <0.01 |
| R Sub thickness PHC EXF | morphometry | -3.7038 | 0.51 | -7.27 | <0.01 |
| R Sub thickness PHC EXF | morphometry:Stable AD | 3.8781 | 0.81 | 4.81 | <0.01 |
| R Sub thickness PHC EXF | morphometry:CN to MCI/AD | -2.5324 | 1.08 | -2.34 | 0.04 |
| R Sub thickness PHC LAN | morphometry | -3.7701 | 0.50 | -7.58 | <0.01 |
| R Sub thickness PHC LAN | morphometry:CN to MCI/AD | -3.4534 | 1.06 | -3.26 | <0.01 |
| R Sub thickness PHC MEM | morphometry:MCI to AD | -3.0361 | 0.61 | -5.02 | <0.01 |
| R Sub thickness PHC MEM | morphometry:Stable MCI | -2.6444 | 0.58 | -4.55 | <0.01 |
| R Sub thickness PHC MEM | morphometry:CN to MCI/AD | -3.7588 | 0.87 | -4.31 | <0.01 |
| R Sub thickness PHC VSP | morphometry:Stable AD | 3.8896 | 1.22 | 3.18 | <0.01 |
| R Sub volume PHC EXF | morphometry:Stable MCI | 0.3045 | 0.09 | 3.32 | <0.01 |
| R Sub volume PHC LAN | morphometry:Stable MCI | 0.4896 | 0.09 | 5.44 | <0.01 |
| R Sub volume PHC LAN | morphometry:CN to MCI/AD | 0.3783 | 0.12 | 3.17 | <0.01 |
| R Sub volume PHC LAN | morphometry:MCI to AD | 0.2709 | 0.09 | 3.15 | <0.01 |
| R Sub volume PHC LAN | morphometry:Stable AD | 0.2443 | 0.10 | 2.46 | 0.03 |
| R Sub volume PHC MEM | morphometry:Stable MCI | 0.6177 | 0.07 | 8.58 | <0.01 |

| | | | | | |
|---|---|---|---|---|---|
| R Sub volume PHC MEM | morphometry:CN to MCI/AD | 0.7735 | 0.10 | 8.11 | <0.01 |
| R Sub volume PHC MEM | morphometry:MCI to AD | 0.4928 | 0.07 | 7.17 | <0.01 |
| R Sub volume PHC MEM | morphometry:Stable AD | 0.4461 | 0.08 | 5.61 | <0.01 |
